# Gender-Related Variables for Health Research

**DOI:** 10.1101/2020.09.17.20196824

**Authors:** Mathias W. Nielsen, Marcia L. Stefanick, Diana Peragine, Torsten B. Neilands, John P. A. Ioannidis, Louise Pilote, Judith J. Prochaska, Mark R. Cullen, Gillian Einstein, Ineke Klinge, Hannah LeBlanc, Hee Y. Paik, Londa Schiebinger

## Abstract

This study develops a gender assessment tool—the Stanford Gender-Related Variables for Health Research—for use in clinical and population research, including large-scale health surveys involving diverse Western populations. While analyzing sex as a biological variable is widely mandated, gender as a sociocultural variable is not, largely because the field lacks quantitative tools for analyzing the influence of gender on health outcomes. We conducted a comprehensive review of English-language measures of gender from 1975 to 2015 to identify variables across three domains: gender norms, gender-related traits, and gender relations. This yielded 11 variables tested with 44 items in three US cross-sectional survey populations: two internet-based (N= 2,051; N= 2,135) and a patient-research registry (N= 489), conducted between May 2017 and January 2018. Exploratory and confirmatory factor analyses distilled 11 constructs to 7 gender-related variables: caregiver strain, work strain, independence, risk-taking, emotional intelligence, social support, and discrimination. Regression analyses, adjusted for age, ethnicity, income, education, sex assigned at birth, and self-reported gender identity, identified associations between these gender-related variables and self-rated general health, physical and mental health, and health-risk behaviors. Our new instrument can be used to develop health interventions based on a fuller understanding of gender associations with health.

## Main Text

### Introduction

Nearly 20 years ago the U.S. Institute of Medicine (IOM, now the Academies of Science) recognized that sex (biology) matters in determining health outcomes but also that gender (sociocultural factors related to social gender norms and expectations, gender-related traits and identities, and gender relations) interacts with sex to influence health and disease processes across the lifespan (1, 2). In the past decade, both the Canadian Institutes of Health Research (2010) (3) and the European Commission (2014) (4) have endorsed integrating sex and gender (usually as male/female binaries) into health research, and the US National Institutes of Health (NIH) has mandated the inclusion of Sex as a Biological Variable (SABV—2016) (5). Yet still today, sex and gender are often inappropriately conflated in the biomedical literature (6), and the NIH has not mandated Gender as a Sociocultural Variable (GASV) largely because the field lacks quantitative tools for analyzing the influence of gender on health outcomes.

Gender behaviors interact with biological sex factors in health outcomes. COVID-19 fatality rates, for example, are proportionally higher in men (about 60%) in most countries, despite a similar proportion of men and women contracting the virus. Sex differences in viral receptor, viral reproduction, antibody production and efficacy, among other factors related to basic biological sex differences, account for some differences, but gender plays a major role as well. For example, smoking rates (higher in men), preventive measures such as hand washing (lower in men), and healthcare seeking (delayed in men) likely contribute to these outcomes (7).

Despite several efforts to examine gender and health (8, 9), the field lacks adequate tools to assess gender. To address this problem, we set out to develop a new instrument – the Stanford Gender-Related Variables for Health Research (GVHR).

Our interest was piqued by a 2007 study that reported that men with higher “femininity” scores had lower risk of coronary heart disease. No such relationship was observed among women (10). Similarly, a 2015 study found that, independent of biological sex, young adults with a gender score more strongly associated with “feminine characteristics” were more likely to experience a recurrence of acute coronary syndrome (ACS)—regardless of whether they identified as a man or a woman (a non-binary option was not offered) (11, 12).

These innovative studies sought to account for both sex and gender. We were concerned, however, that the conclusions were based on outdated gender constructs, such as the Bem Sex Role Inventory (BSRI, 1974) (13), developed using predominantly white, higher socioeconomic U.S. undergraduate student participants and based on outdated notions of masculinity and femininity (9, 10) or their cognates. We also saw that the GENESIS-PRAXY gender score used in the 2015 study was developed and tested in a sample of 909 heart-disease patients and had not been cross-validated in broader patient, or non-patient, populations. We were further concerned that despite its focus on gender, the GENESIS-PRAXY study used logistic regression with biological sex as the outcome to determine which variables to include in its final score. We also found that the internal consistency of the measure had not been explicitly tested.

More importantly, we were concerned that gender was reduced to a bipolar index of “masculinity” and “femininity,” i.e. concepts which were historically construed as complementary oppositions (16), with a man being one thing and a woman being the opposite (for example, rational/emotional; public/private; mind/body). These concepts are too broad and imprecise to be useful in health research. If the goal is to provide physicians and policy makers with gender-related health interventions, measures of gender-related behaviors, such as caregiving or risk-taking, should be labelled as such and not reduced to a “masculinity” versus “femininity” score.

To address these limitations, we develop a new gender-variables instrument. In a systematic review of the English-language literature from 1975 to 2015, we identified 74 eligible scales used in gender-related measures (see Materials and Methods). Several limitations made it impossible to simply plug them into a new questionnaire. Very few existing scales (N=8) focus on associations between gender and health (17–21). The eligible gender-related scales are generally restricted to either men or women and assess either masculinity (22) or femininity (23, 24), or both as unipolar or bipolar constructs (13, 25–31) (e.g., hyper-masculinity or hyper-femininity) (19, 21, 32–42). Further, most scales rely on “agree-disagree” ratings, making them susceptible to acquiescence bias (43).

From the 74 scales, we distilled 11 composite gender constructs and developed 44 items to measure gender, which we subjected to exploratory and confirmatory factor analysis (EFA and CFA) in three independent U.S. survey samples. This reduced the original 11 gender-related variables to 7 factors: caregiver strain, work strain, independence, risk-taking, emotional intelligence, social support, and discrimination and the 44 survey items to 25 (see Materials and Methods). We then examined the relevance of the derived subscales in allowing for more precise analysis of variations in health-related quality of life, obesity, and risky health behaviors. This initial step toward developing more comprehensive and precise survey-based measures of gender, in relation to health, captures key aspects of three dimensions of gender that can be deployed quantitatively in diverse clinical research or large health surveys: 1) Gender Norms; 2) Gender-Related Traits and 3) Gender Relations (see Tables S1 and S2), each of which may correlate with sex assigned at birth or self-reported gender identity, without a predetermined coding of that behavior as masculine or feminine.

**Box 1. Three interrelated dimensions of gender**

Gender refers to culturally constructed norms and relations that shape the identities, attitudes, behaviors, bodily appearances, and habits of women, men, and gender-diverse individuals. Gender is multidimensional and complex, changing as social norms and values change. Gender also intersects with other sociocultural categories. The gender variables developed here appear alongside self-reported variables collected in our survey, including: sex assigned at birth, self-reported gender identity, sexual orientation, ethnicity, age, individual and household income, and education, among others (see SI text).

- **Gender Norms** consist of legislated and both spoken and unspoken cultural rules produced through social institutions (such as governments, families, schools, workplaces, laboratories, universities, or boardrooms), cultural products (such as technologies, science, literature, and social media), and broader local and global cultures (77, 78). Here, we measure individuals’ adherence to gender norms through self-reported behaviors (not attitudes or stereotypes).
- **Gender-Related Traits** refers to how individuals or groups perceive and present themselves in relation to gender norms. Our key interest is how individuals think and act vis-à-vis cultural meanings ascribed to gender (79, 80). Thus, we measure gender-related traits through self-reported assessments of personality attributes.
- **Gender Relations** refer to how individuals interact with other people and institutions in specific sociocultural contexts. This dimension encompasses how gender shapes social interactions in families, schools, workplaces, and other public settings (78, 81). We measure gender relations through self-reported assessments of social relations between people of different gender identities within social institutions and societies at large.

These three meta-categories are not mutually exclusive. Gender is multidimensional: any given individual may experience different configurations of gender norms, traits, and relations that cannot be subsumed into a “masculine” or “feminine” score or considered “fixed”.

## Results

### EFA, CFA, and Reliability Assessment

Table 1 reports the standardized regression weights, Raykov’s ρ, and item scoring for the trimmed CFA model in samples 1, 2 and 3, with seven variables. The 7 factors listed above represent our final gender-related variables. Our analyses yielded low to moderate inter-factor correlations (Table S3), which is not unusual in multidimensional gender measures (19, 44, 45). We calculated mean-item subscale scores for each factor and used these as predictors in the regressions presented below. For subscales including continuous variables, mean-item scores were calculated based on standardized variables (z-scores). All variables are scored from lower to higher levels of the given constructs.

**Table 1.**
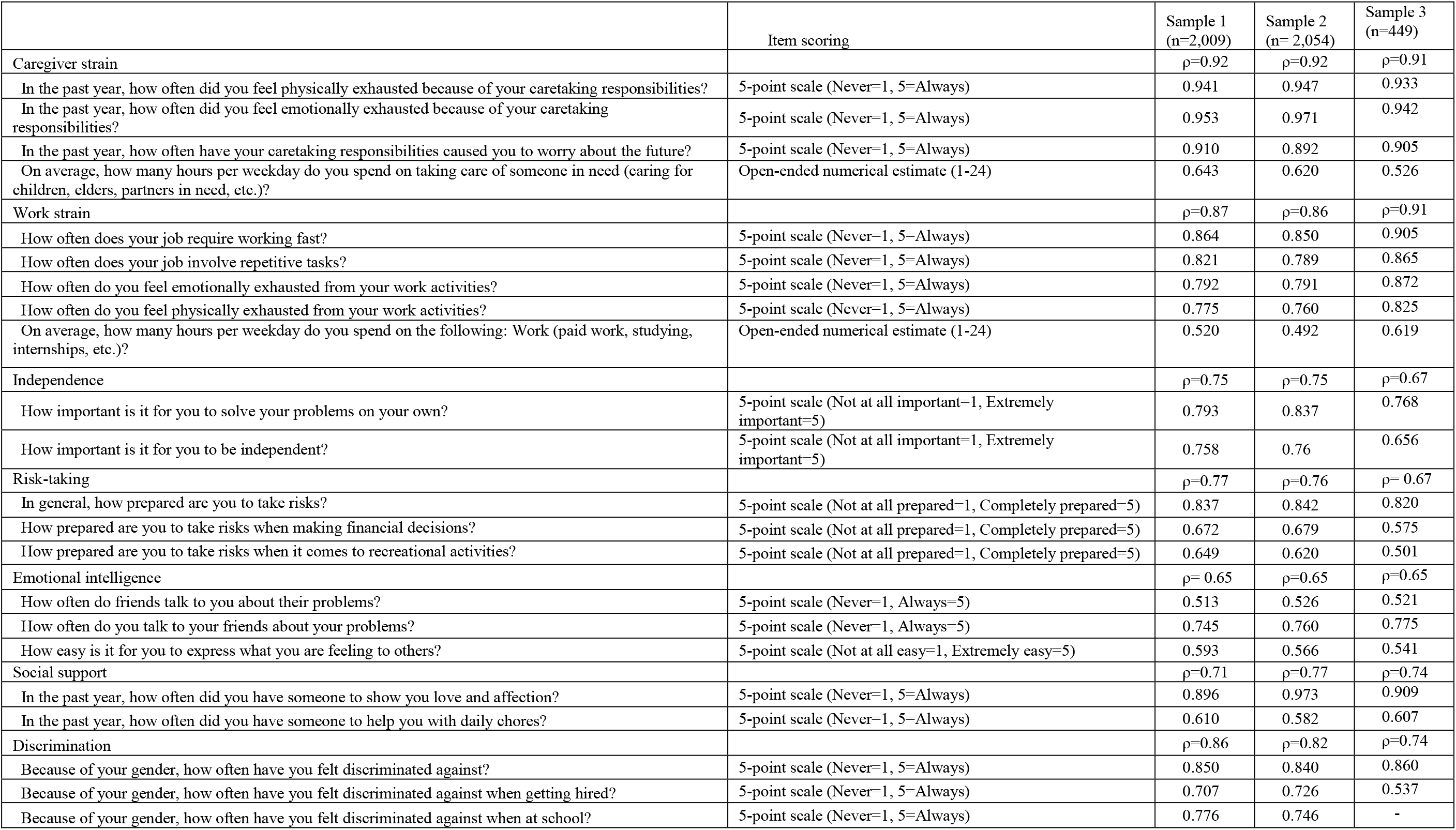
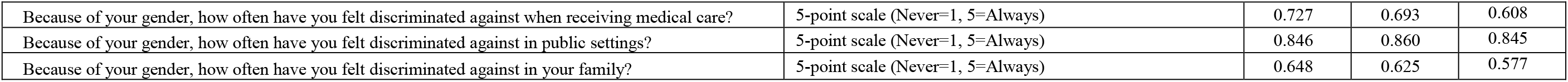
Factor loadings for the trimmed CFA models in Samples 1, 2 and 3 and Raykov’s ρ for each factor.

|  | Item scoring | Sample 1<br>(n=2,009) | Sample 2<br>(n= 2,054) | Sample 3<br>(n=449) |
| --- | --- | --- | --- | --- |
| Caregiver strain | | $\rho=0.92$ | $\rho=0.92$ | $\rho=0.91$ |
| In the past year, how often did you feel physically exhausted because of your caretaking responsibilities? | 5-point scale (Never=1, 5=Always) | 0.941 | 0.947 | 0.933 |
| In the past year, how often did you feel emotionally exhausted because of your caretaking responsibilities? | 5-point scale (Never=1, 5=Always) | 0.953 | 0.971 | 0.942 |
| In the past year, how often have your caretaking responsibilities caused you to worry about the future? | 5-point scale (Never=1, 5=Always) | 0.910 | 0.892 | 0.905 |
| On average, how many hours per weekday do you spend on taking care of someone in need (caring for children, elders, partners in need, etc.)? | Open-ended numerical estimate (1-24) | 0.643 | 0.620 | 0.526 |
| Work strain | | $\rho=0.87$ | $\rho=0.86$ | $\rho=0.91$ |
| How often does your job require working fast? | 5-point scale (Never=1, 5=Always) | 0.864 | 0.850 | 0.905 |
| How often does your job involve repetitive tasks? | 5-point scale (Never=1, 5=Always) | 0.821 | 0.789 | 0.865 |
| How often do you feel emotionally exhausted from your work activities? | 5-point scale (Never=1, 5=Always) | 0.792 | 0.791 | 0.872 |
| How often do you feel physically exhausted from your work activities? | 5-point scale (Never=1, 5=Always) | 0.775 | 0.760 | 0.825 |
| On average, how many hours per weekday do you spend on the following: Work (paid work, studying, internships, etc.)? | Open-ended numerical estimate (1-24) | 0.520 | 0.492 | 0.619 |
| Independence | | $\rho=0.75$ | $\rho=0.75$ | $\rho=0.67$ |
| How important is it for you to solve your problems on your own? | 5-point scale (Not at all important=1, Extremely important=5) | 0.793 | 0.837 | 0.768 |
| How important is it for you to be independent? | 5-point scale (Not at all important=1, Extremely important=5) | 0.758 | 0.76 | 0.656 |
| Risk-taking | | $\rho=0.77$ | $\rho=0.76$ | $\rho=0.67$ |
| In general, how prepared are you to take risks? | 5-point scale (Not at all prepared=1, Completely prepared=5) | 0.837 | 0.842 | 0.820 |
| How prepared are you to take risks when making financial decisions? | 5-point scale (Not at all prepared=1, Completely prepared=5) | 0.672 | 0.679 | 0.575 |
| How prepared are you to take risks when it comes to recreational activities? | 5-point scale (Not at all prepared=1, Completely prepared=5) | 0.649 | 0.620 | 0.501 |
| Emotional intelligence | | $\rho=0.65$ | $\rho=0.65$ | $\rho=0.65$ |
| How often do friends talk to you about their problems? | 5-point scale (Never=1, Always=5) | 0.513 | 0.526 | 0.521 |
| How often do you talk to your friends about your problems? | 5-point scale (Never=1, Always=5) | 0.745 | 0.760 | 0.775 |
| How easy is it for you to express what you are feeling to others? | 5-point scale (Not at all easy=1, Extremely easy=5) | 0.593 | 0.566 | 0.541 |
| Social support | | $\rho=0.71$ | $\rho=0.77$ | $\rho=0.74$ |
| In the past year, how often did you have someone to show you love and affection? | 5-point scale (Never=1, 5=Always) | 0.896 | 0.973 | 0.909 |
| In the past year, how often did you have someone to help you with daily chores? | 5-point scale (Never=1, 5=Always) | 0.610 | 0.582 | 0.607 |
| Discrimination | | $\rho=0.86$ | $\rho=0.82$ | $\rho=0.74$ |
| Because of your gender, how often have you felt discriminated against? | 5-point scale (Never=1, 5=Always) | 0.850 | 0.840 | 0.860 |
| Because of your gender, how often have you felt discriminated against when getting hired? | 5-point scale (Never=1, 5=Always) | 0.707 | 0.726 | 0.537 |
| Because of your gender, how often have you felt discriminated against when at school? | 5-point scale (Never=1, 5=Always) | 0.776 | 0.746 | - |
| Because of your gender, how often have you felt discriminated against when receiving medical care? | 5-point scale (Never=1, 5=Always) | 0.727 | 0.693 | 0.608 |
| Because of your gender, how often have you felt discriminated against in public settings? | 5-point scale (Never=1, 5=Always) | 0.846 | 0.860 | 0.845 |
| Because of your gender, how often have you felt discriminated against in your family? | 5-point scale (Never=1, 5=Always) | 0.648 | 0.625 | 0.577 |

Figure 1 displays the z-scores (averaged by group) for the 7 gender-related variables for respondents seeing themselves as men, women, and gender fluid/non-binary in sample one. The figure demonstrates the advantage of capturing specific gender-related behaviors and attitudes through multiple variables.

**Fig 1.**
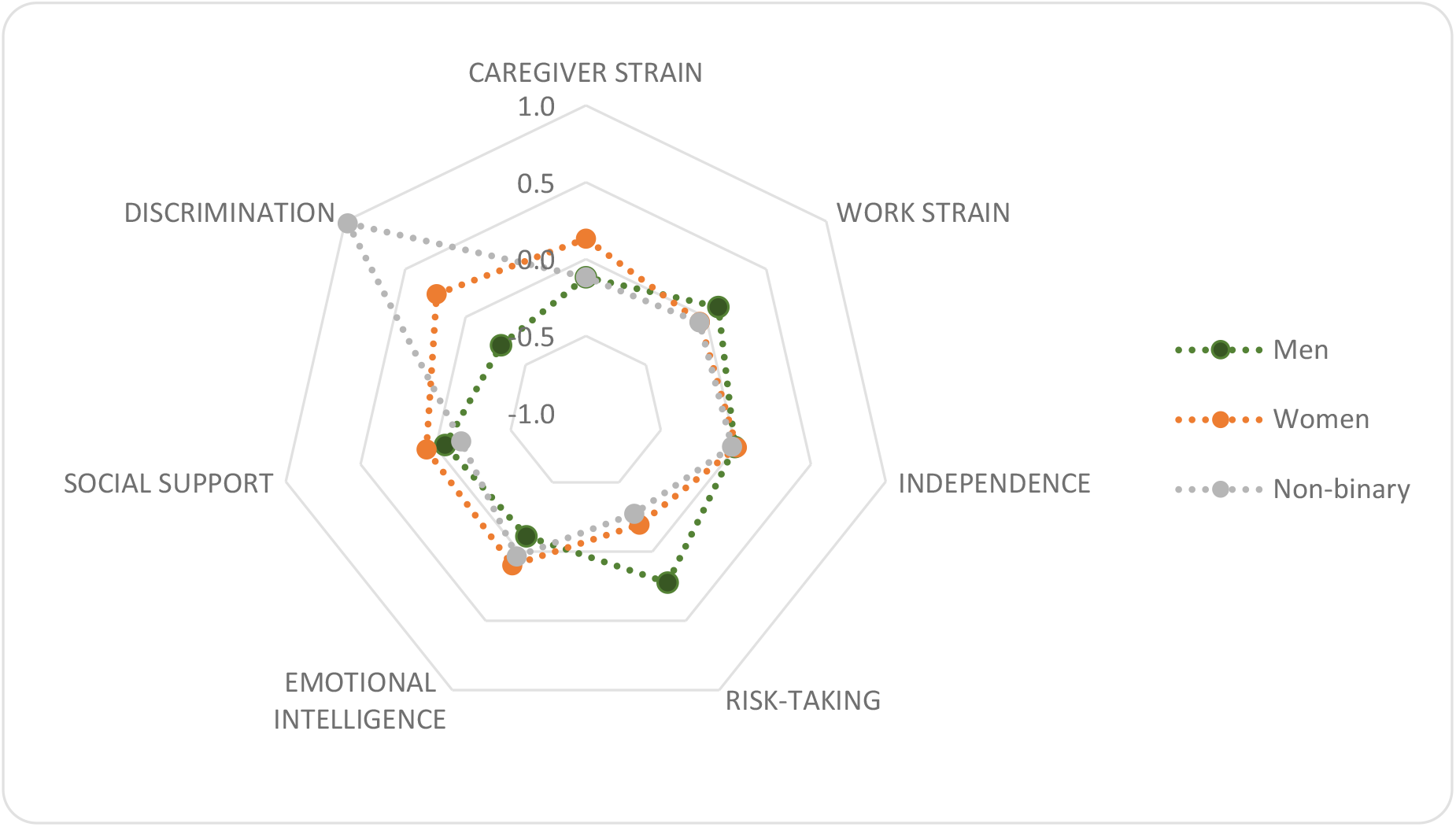
Gender-related variables capturing specific behaviors and attitudes. The figure displays the z-scores for the seven gender-related variables for respondents seeing themselves as men (green), women (orange) and gender fluid/Non-binary (grey) in sample 1 (N= 1893).

### Associations with self-rated health and health-risk behaviors

Existing research shows notable sex differences in health-related quality of life, obesity, and risky health behaviors, such as smoking and heavy drinking (46–48). Here, we examine the relevance of our gender-related variables in predicting self-related health and health-risk behaviors.

Associations between our 7 gender-related variables, sex assigned at birth, self-reported gender identity and physical health, mental health, and activity limitations due to poor physical or mental health were analyzed in samples 1 and 2 using negative binomial regressions, with birth year, personal income, education level, race, and ethnicity as covariates. Only associations that are consistent across samples 1 and 2 are reported here. Measures of reported birth sex and gender identity were highly correlated. Therefore, we ran all models twice: once including birth sex and once including gender identity. Here, we report the outcomes of the models including birth sex as a key predictor (see Tables S4-S6 for specifications on the regression models including gender identity).

As displayed in Table 2, caregiver strain and discrimination were associated with lower physical health, mental health, and activity levels in both samples, whereas social support was associated with higher mental health and activity levels in both samples. The results for the remaining associations were inconclusive in one or both samples.

**Table 2.** Adjusted incidence rate ratios and 95% confidence intervals of associations with health-related measures in negative binomial regressions

|  | Physical health<sup>a</sup> |  | Mental health<sup>b</sup> |  | Activity limitations<sup>c</sup> |  |
| --- | --- | --- | --- | --- | --- | --- |
|  | Sample 1 | Sample 2 | Sample 1 | Sample 2 | Sample 1 | Sample 2 |
|  | (n= 1,874) | (n= 2,005) | (n= 1,876 ) | (n= 2,004) | (n= 1,872) | (n= 2,004) |
| Caregiver strain | 1.20 (1.12:1.29) | 1.11 (1.04:1.18) | 1.21 (1.14:1.27) | 1.20 (1.14:1.27) | 1.13 (1.04:1.21) | 1.11 (1.03:1.19) |
| Work strain | 0.96 ( 0.87: 1.05) | 1.02 (0.94:1.11) | 1.03 (0.96:1.11) | 1.12 (1.04:1.21) | 0.88 (0.80:.96) | 0.99 (0.90:1.08) |
| Independence | 0.93 (0.85:1.01) | 0.84 (0.75:.95) | 0.97 (0.91:.1.04) | 0.83 (0.74:.94) | 1.02 (0.94:1.12) | 0.87 (0.77:1.00) |
| Risk-taking | 0.95 (0.87:1.04) | 1.00 (0.92:1.08) | 0.86 (0.80:.92) | 0.99 (0.91:1.08) | 0.93 (0.85:1.01) | 1.02 (0.94:1.12) |
| Emotional intelligence | 0.93 (0.84:1.03) | 0.93 (0.85:1.01) | 0.97 (0.89:1.05) | 0.86 (0.78:.93) | 0.98 (0.88:1.10) | 0.92 (0.84:1.02) |
| Social support | 0.95 (0.89:1.02) | 0.94 (0.89:1.00) | 0.86 (0.81:.90) | 0.81 (0.76:.86) | 0.92 (0.86:.99) | 0.87 (0.81:.94) |
| Discrimination | 1.46 (1.34:1.59) | 1.43 (1.30:1.57) | 1.44 (1.34:1.53) | 1.35 (1.24:1.48) | 1.76 (1.61:1.93) | 1.56 (1.40:1.73) |
| Sex (male=0) | 0.94 (0.81:1.10) | 0.93 (0.80:1.08) | 0.94 (0.83:1.06) | 1.21 (1.05:1.40) | 0.88 (0.74:1.04) | 1.11 (0.94:1.31) |

Note: Incidence rate ratios are adjusted for year of birth, ethnicity, race, education and personal income. See Tables S12-S14 for model specifications.
<sup>a</sup> Number of days with poor physical health during the last 30 days.
<sup>b</sup> Number of days with poor mental health during the last 30 days.
<sup>c</sup> Number of days with activity limitations due to poor physical and mental health during the last 30 days.

Associations between our gender-related variables, sex assigned at birth, self-reported gender identity and general health status, smoking, vaping, binge drinking, and BMI were analyzed using logistic regressions in samples 1 and 2 (Table 3, adjusted for year of birth, personal income, education level, ethnicity, and race), and we ran separate models with sex and gender identity (see Tables S7-S11 for specifications on the regression models including gender identity). In both samples, caregiver strain, discrimination, and male birth sex were associated with fair or poor self-rated health, while risk-taking and social support predicted good, very good, or excellent self-rated health. Further, caregiver strain and work strain were associated with smoking, while discrimination was associated with vaping and higher levels of risk-taking was associated with binge drinking. Caregiver strain, low levels of risk-taking, discrimination, and male birth sex were associated with overweight. The results for the remaining associations were inconclusive in one or both samples.

**Table 3.** Adjusted odds ratios and 95% CIs of associations with health-related measures in logistic regressions

|  | Health status<sup>a</sup> |  | Smoking<sup>b</sup> |  | Vaping<sup>c</sup> |  | Binge drinking<sup>d</sup> |  | BMI<sup>e</sup> |  |
| --- | --- | --- | --- | --- | --- | --- | --- | --- | --- | --- |
|  | Sample 1 | Sample 2 | Sample 1 | Sample 2 | Sample 1 | Sample 2 | Sample 1 | Sample 2 | Sample 1 | Sample 2 |
|  | (n= 1,884) | (n= 2,010) | (n= 1,883) | (n= 2,009) | (n= 1,882) | (n= 2,009) | (n= 1,880) | (n= 2,007) | (n=1,858) | (n=1,993) |
| Caregiver strain | 1.28 (1.11:1.47) | 1.18 (1.03:1.34) | 1.42 (1.24:1.62) | 1.18 (1.05:1.33) | 1.06 (0.90:1.26) | 1.10 (0.93:1.29) | 0.92 (0.81:1.06) | 0.91 (0.80:1.04) | 1.18 (1.05:1.32) | 1.27 (1.13:1.41) |
| Work strain | 0.88 (0.74:1.06) | 0.98 (0.83:1.17) | 1.37 (1.15:1.64) | 1.26 (1.08:1.46) | 1.23 (0.97:1.55) | 1.29 (1.05:1.59) | 1.28 (1.08:1.51) | 1.03 (0.88:1.21) | 1.04 (0.91:1.19) | 1.17 (1.02:1.33) |
| Independence | 0.91 (0.77:1.07) | 0.71 (0.56:0.89) | 1.22 (1.03:1.46) | 1.07 (0.85:1.35) | 1.17 (0.95:1.45) | 1.05 (0.76:1.44) | 1.08 (0.93:1.25) | 1.19 (0.94:1.49) | 0.99 (0.88:1.12) | 1.05 (0.87:1.26) |
| Risk-taking | 0.57 (0.48:0.68) | 0.69 (0.58:0.81) | 1.45 (1.21:1.72) | 1.05 (0.91:1.21) | 1.20 (0.98:1.48) | 1.00 (0.80:1.25) | 1.53 (1.31:1.78) | 1.25 (1.07:1.45) | 0.82 (0.72:0.93) | 0.85 (0.75:0.97) |
| Emotional intelligence | 0.91 (0.76:1.10) | 0.90 (0.75:1.07) | 1.46 (1.22:1.75) | 1.12 (0.96:1.30) | 1.39 (1.12:1.71) | 1.05 (0.84:1.31) | 1.18 (1.00:1.40) | 1.04 (0.88:1.21) | 0.99 (0.86:1.14) | 1.03 (0.90:1.18) |
| Social support | 0.83 (0.73:0.95) | 0.78 (0.69:0.88) | 0.98 (0.86:1.12) | 0.97 (0.87:1.09) | 0.98 (0.84:1.14) | 0.88 (0.76:1.04) | 1.01 (0.90:1.13) | 0.87 (0.78:0.97) | 1.00 (0.91:1.10) | 1.07 (0.97:1.18) |
| Discrimination | 1.43 (1.18:1.73) | 1.55 (1.29:1.87) | 1.09 (0.90:1.31) | 1.16 (0.97:1.38) | 1.40 (1.15:1.70) | 1.46 (1.17:1.81) | 1.18 (1.00:1.39) | 1.11 (0.92:1.32) | 1.23 (1.06:1.42) | 1.26 (1.08:1.48) |
| Sex (male= 0) | 0.60 (0.44:0.82) | 0.65 (0.48:0.87) | 0.66 (0.48:0.89) | 0.89 (0.69:1.16) | 0.45 (0.31:0.64) | 0.76 (0.52:1.11) | 0.55 (0.42:0.71) | 0.77 (0.59:1.01) | 0.76 (0.60:0.95) | 0.58 (0.46:0.73) |

Note: Odds ratios are adjusted for year of birth, ethnicity, race, education and personal income. Tables S15-S19 for model specifications.
<sup>a</sup> Good, very good, excellent=0, fair, and poor=1
<sup>b</sup> Not smoking=0, smoking=1
<sup>c</sup> Not vaping=0, vaping=1
<sup>d</sup> Less than monthly=0, monthly, weekly, and daily=1
<sup>e</sup> Under or normal weight (BMI<25)=0, overweight or obese (BMI≥25)=1

Combining the data from samples 1 and 2 (and adjusting for sample in the regression models), all associations with self-related health and health-risk behaviors reported above persisted at the 99.9% confidence level (Figures 2 and 3), as did the following associations: emotional intelligence with smoking and male birth sex with vaping and binge drinking.

**Fig 2.**
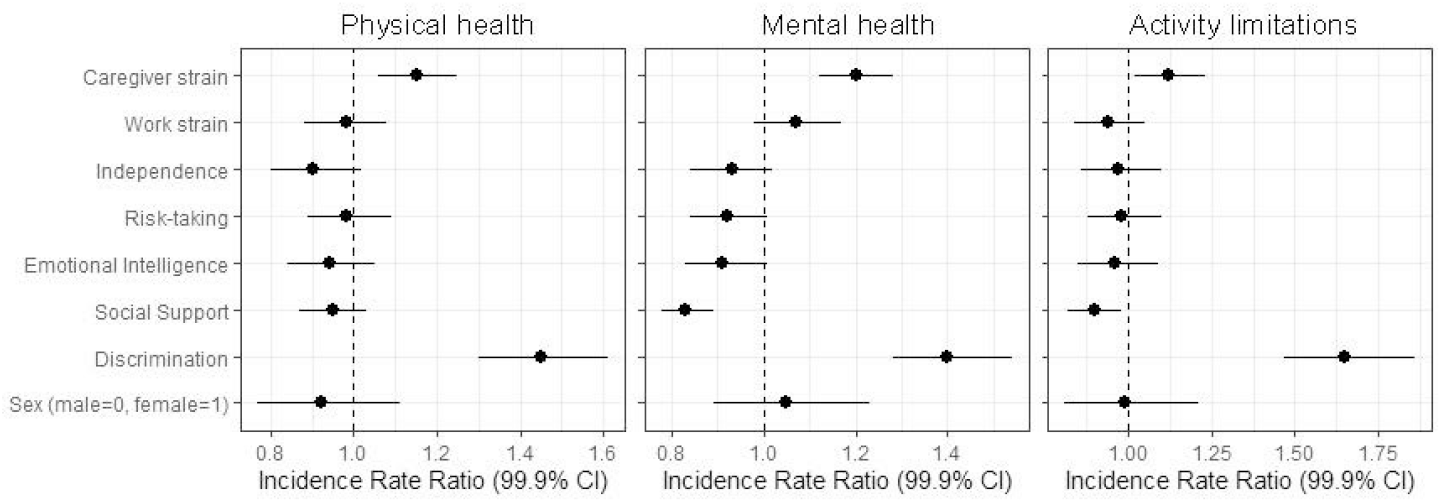
Adjusted incidence rate ratios of associations with recent physical health, mental health and activity limitations. This figure displays the outcomes of the negative binomial regressions predicting health outcomes in the combined sample (sample 1 + sample 2) (Physical health, N=3,879; mental health, N=3,880; activity limitations, N=3,876). Error bars represent 99.9% confidence intervals. See Tables S20-S22 for model specifications.

**Fig 3.**
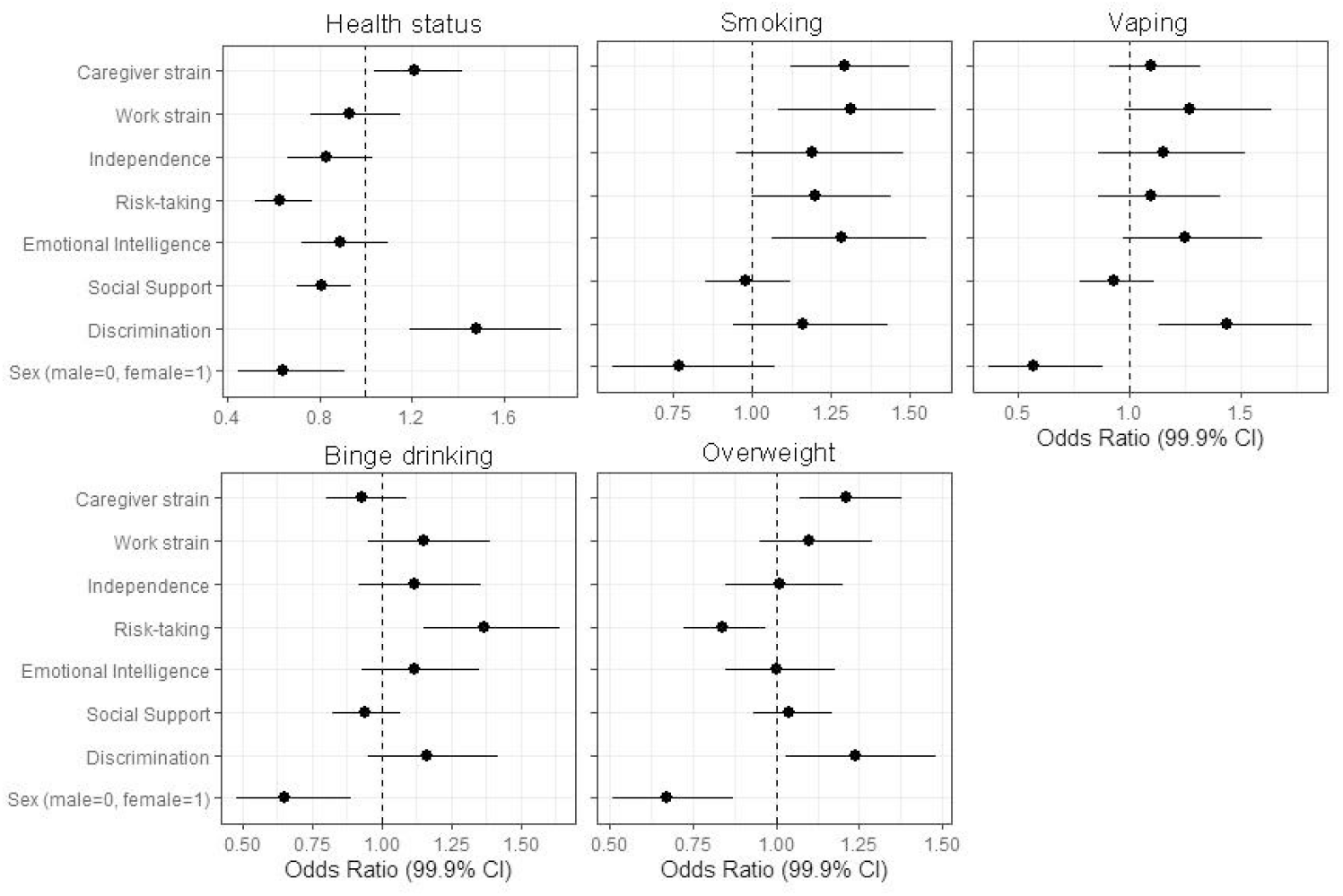
Adjusted odds ratios of associations with health status, smoking, vaping, binge drinking and BMI. This figure displays the outcomes of the binary logistic regressions predicting health outcomes in the combined sample (sample 1 + sample 2) (health status, N= 3,894; smoking, N=3,892 vaping, N=3,891; binge drinking, N=3,887; BMI, N= 3,851. Error bars represent 99.9% confidence intervals. See Tables S23-S27 for model specifications.

## Discussion

Following a comprehensive review of gender measures from 1975 to 2015, we have applied a rigorous process to identify key aspects of gender for the purpose of developing a new gender assessment tool for use in clinical and population research, including large-scale health surveys involving diverse Western populations. Through exploratory and confirmatory factor analyses, we reduced the original 44 survey items to 25 and the 11 original constructs to 7 gender-related variables: caregiver strain, work strain, independence, risk-taking, emotional intelligence, social support, and discrimination.

Each variable captures an important aspect of gender within the populations studied. Each variable measures an individual participant’s self-reported behavior or attitude for that characteristic and is designed to be scored individually, as a distinct human behavior or attribute. Behaviors are not coded “masculine” or “feminine,” and we recommend against consolidating the variable scores into these unipolar or bipolar indices. Studies that reduce gender-related variables to “femininity” or “masculinity” scores give little guidance for behavioral interventions.

For example, if caregiver strain is found to be associated with higher risk of recurrence or death in patients with ACS, it should be reported as such. Subsuming gender-related factors into masculine or feminine indices will reduce, rather than improve, the precision and applicability of survey-based measures of health.

Clinicians and public health researchers can employ the Stanford GVHR to gain a fuller understanding of gender differences in health outcomes than can be captured by simply asking people to self-identify their sex or gender. Specifically, we recommend that researchers start by measuring all 7 variables alongside sex assigned at birth, self-reported gender identity, and other relevant factors, such as sexual orientation, ethnicity, age, income, and education, to specify which characteristics, traits and behaviors may be predictive of the health issue in focus. In subsequent studies and patient-based outcome measures, the focus may be restricted to a subset of variables of documented relevance to a specific disease or health condition.

The Stanford GVHR is specific to the survey cohorts, time, place, and culture in which it was developed and tested. Because gender norms, traits, and relations vary across and within cultures and change over time (49), we recommend that variables be updated with each generation or even more frequently. Obvious limitations are that our variables were developed from English-language literature and validated in nonprobability samples, recruited online and through a research registry in U.S. populations, and the age composition of our samples (median ages range from 36 to 50 years) does not represent typical patient populations encountered in clinical practice. We strongly encourage more research to test the validity of the gender-related variables within and across age groups and cultures, as well as across a wide range of global settings.

Any association of a gender-related variable with a health outcome does not infer causality. Our assessment was cross-sectional. Confounding or even reverse causality (health phenotypes affecting gender-related behaviors and attitudes) should be considered. Another limitation is the small number of items per construct which might limit generalizability; at the same time, a smaller number of items likely increases the usefulness of the questionnaire to practitioners and researchers. An avenue for further research is the expansion the number of items for each construct, especially emotional intelligence, which had less than ideal reliability across all three samples. In addition, a few of the initial gender-related constructs that were not retained in the exploratory factor analysis, such as competition and quality of family relationships, may have been deselected due to insufficient items in the study’s initial pool of attitudes and behaviors and should be reconsidered in future research.

In the future, the proposed list of variables to measure other gender-related factors could be expanded to integrate factors such as decision-making power (including over household resources and health expenditures) and the distribution of labor among both same- and different-sex cohabiting or romantic partners. Indeed, many of the measures that have informed our work may be outdated (noting that some date back to the 1970s and 1980s). It may be desirable to complement our proposed list of variables with more “timely” variables that better represent how specific patients and persons conceive of gender in 2020. It would also be interesting to explore associations between our gender-related variables and other health-related aspects such as health literacy, health-seeking behavior, and provider-patient interactions.

In conclusion, this project is an initial step toward developing more comprehensive and precise survey-based measures of gender in relation to health. Our questionnaire is designed to shed light on how specific gender-related behaviors and attitudes contribute to health and disease processes, irrespective of—or in addition to—biological sex and self-reported gender identity. Use of these gender-related variables in experimental studies, such as clinical trials, may also help us understand if gender has an important role as treatment effect modifiers and would thus need to be further considered in treatment decision-making.

## Materials and Methods

Our research complies with all ethical regulations. Stanford University’s Institutional Review Board approved the study. We obtained informed consent from all participants.

### Literature search

We searched PsycINFO, PsycTESTS, and PubMed for all English-language studies using gender-related tests and scales, from 1975 through 2015, to identify existing questionnaires or scales and construct a comprehensive list of typical traits and/or characteristics used in gender-related measures in both psychology and medicine. We screened an initial sample of 2,981 articles from PubMed, PsycTESTS, and PsycINFO from which 405 articles were deemed relevant for further interrogation, within which 127 unique gender-related test and scales were identified.

We also screened existing literature reviews published in books and found four additional scales. Altogether 131 gender-related questionnaires were sorted into three overarching categories for analyzing gender norms, gender-related traits, and gender relations. Further, we checked citation frequencies for each scale to determine how often it has been used in the literature. All gender scales with at least 20 Google Scholar citations within the last 10 years were selected for further investigation, of which 74 scales met the criteria. All articles published from 2006-2015 were retained for further investigation (the search methods, selection criteria, and review procedures are specified in SI text, Figure S1, and Tables S28-S30).

### Questionnaire development

In developing the questionnaire, we recognized the need to minimize the time burden of completing the questionnaire; therefore, we limited the initial item pool to 3-6 items per construct (see Methods). To avoid acquiescence bias, we presented our items as construct-specific questions. The 74 scales guided our selection of core characteristics to be included among our gender-related measures.

#### 1. Gender Norms

Guided by three of the 74 eligible scales, respondents’ adherence to gender norms was measured by three composite constructs (caregiver strain, time use, and work strain) consisting of 16 items.

*Caregiver strain* captures perceived consequences of responsibility for unpaid, long-term caregiving to children, partners, friends, and elderly (excluding housework and caregiving occupations) (50, 51) and consists of three items, adapted from Graessel and colleagues (52), recorded on a five-point scale: emotional exhaustion, physical exhaustion, and worries about the future caused by caregiving for someone in need, such as a child, elder, partner, or disabled family member. Higher scores on these items indicate higher levels of caregiver strain.

*Time use* measures individual hours spent per day, recorded as open-ended numerical estimates of daily time-spent (53), in the following categories: paid work, household activities, eating and drinking, leisure and sport, caring for others, sleeping, and commuting. The items were adapted from the American Time Use Survey.

*Work strain* measures job strain and emotional job demands as six items, recorded on a five-point scale: work speed, work repetition, emotional job demands, physical job demands, perceived risk, and physical hazards at work. The first four items were adapted from Karasek and Theorell (54); the last two were developed by the authors. Higher scores on each item indicate higher levels of work strain.

#### 2. Gender-Related Traits

Guided by 31 of the original 74 eligible scales, gender-related traits were measured as five composite constructs: competitive, risk-taking, independence, communal, and expressive, consisting of 16 items.

*Competitive* consists of two items, recorded on a five-point scale, asking respondents how often they find themselves competing with others in situations that do not call for competition and how competitive they are in general, compared to others. The first item is modified from Ryckman et al. (55), the second is developed by the authors. Higher scores on each item indicate higher levels of competitiveness.

*Risk-taking* focuses on physical and behavioral, measured by three items adapted from Dohmen et al. (56), recorded on a five-point scale: general risk-taking behavior, risk-taking when making financial decisions, and risk-taking with respect to recreational activities. Higher scores on each item indicates higher levels of risk-taking.

*Independence* is a personality trait characterized by a focus on the person as an individual, not as part of a community or group, which includes agency, self-confidence, self-determination, and decision-making ability, but not self-control or self-esteem (57). Independence was based on three items, recorded on a five-point scale, adapted from Bakker et al. (58), Clark et al. (59), and Triandis et al. (60), asking respondents how important it is for them to be independent, how often they turn to others for help when in need, and how important it is for them to solve their problems independently. A higher score indicates a higher level of independence for all three items.

*Communal* is a trait characterized by a focus on the individual as part of a group or community, an orientation towards relationships and a concern for others’ needs and well-being (61). We used four items, scored on a five-point scale, asking respondents how often they worry about what other people think about them, how often they take other people’s needs into account when making important decisions, how often friends talk to them about their problems, and how easy it is for them to spot when someone in a group is feeling uncomfortable. Item one was developed by the authors, item two was adapted from Clark et al. (59), and items three and four were adapted from Baron-Cohen and Wheelwright (62). Higher scores indicate higher levels of communal orientation.

*Expressive* captures abilities to recognize and express emotions, such as sadness, anger, frustration, compassion, joy, or affection and includes aspects of emotional intelligence, i.e. individuals’ ability to recognize what they feel, manage those emotions, and use emotions in problem solving (63). We used four items, recorded on a five-point scale, asking respondents how often they talk to friends about their problems, how easy it is for them to understand their own feelings, how easy it is for them to express what they are feeling, and how easy it is for them to ask other people for help when in need. Item one was developed by the authors, item two was adapted from Salovey and colleagues (64), item three was adapted from Gross and John (65), and item four was adapted from Clark and colleagues (59). Higher scores indicate higher levels of expressivity.

#### 3. Gender Relations

Guided by 40 eligible scales identified in the literature search, gender relations was measured by three composite constructs: social support, discrimination, and quality of family relationships, consisting of 13 items and a single-item measure of personal income.

*Social support* captures perceived satisfaction with the type (physical, emotional, informational, and financial), the availability, and level of support a person might receive. Support may come from partners, relatives, friends, coworkers, health-care systems, the larger community, etc.

Social support consists of four items recorded on a five-point-scale measuring the availability and level of social support a person might receive. We asked respondents how often, within the past year, they had someone they could ask for advice, someone to show them love and affection, someone to help them with daily chores, and how often they felt lonely. Item one, two, and three were adapted from the ENRICHD Social Support Inventory (66) and item four was developed by the authors. Higher scores indicate higher levels of social support.

*Discrimination* refers to “systemic unfair treatment” and can occur at multiple levels. We measured micro or interpersonal discrimination. Specifically, we asked the respondents how often they had felt discriminated against because of their gender, in general, when getting hired, when at school, when receiving medical care, in other public settings, and in their family. The items were recorded on a five-point-scale, with higher scores indicating more frequent experiences of gender discrimination.

*Quality of family relationships* measures experiences of harmony and conflict in familial settings, based on two self-developed items recorded on a five-point-scale asking respondents about how they would describe the quality of their relationship with close relatives (in the past year) and how often they had argued with close relatives (in the past year). Higher scores indicate higher levels of perceived quality in family relationships.

### Initial testing of the questionnaire

Content validity of the first draft of the questionnaire was assessed by nine members of the author-group who had not taken part in the construction of the item-list, some with technical expertise in the construction of survey-questionnaires and some with expertise in gender-related aspects of health. Each coauthor was asked to rate each item with respect to its relevance in measuring a given variable construct. Ratings were made on a four-point scale (4= very relevant, 3= relevant but needs minor alteration, 2= unable to assess relevance without major revision, 1= not relevant). These coauthors were also invited to suggest additional items, if they considered the proposed item-list inadequate in capturing a given construct. To improve item quality, we conducted seven cognitive interviews with people recruited by posters in the local area who varied on demographic markers such as education-level, job, age, ethnicity, and gender. In the interviews, we used verbal probing techniques to identify questions that the interviewees found vague and unclear and to elicit how they arrived at answers to the questions. We did this by asking them to reflect on what the items meant to them, how they would rephrase the question in their own words, how they came up with their answers to the questions, and whether the questions were easy or hard to answer and why.

### Survey Participants

Participants were recruited from the U.S. through two online services and a health research registry: Prolific, Amazon Mechanical Turk, and the Stanford Research Registry, which consists of ∼ 4,000 former adult patients at the Stanford University Medical Center who have agreed to be contacted for participation in research studies. We used the web-based Qualtrics software to collect data from Prolific (sample 1) in August and September 2017, Mechanical Turk (sample 2) in December 2017 and January 2018, and the Stanford Research Registry in May and June 2017 (sample 3). Sample 1 consisted of 2,051 respondents; 1,992 completed the survey. Sample 2 consisted of 2,135 respondents; 2,043 completed the survey. Sample 3 consisted of 489 respondents; 452 completed the survey. Sample characteristics are presented in Table 4.

**Table 4.**
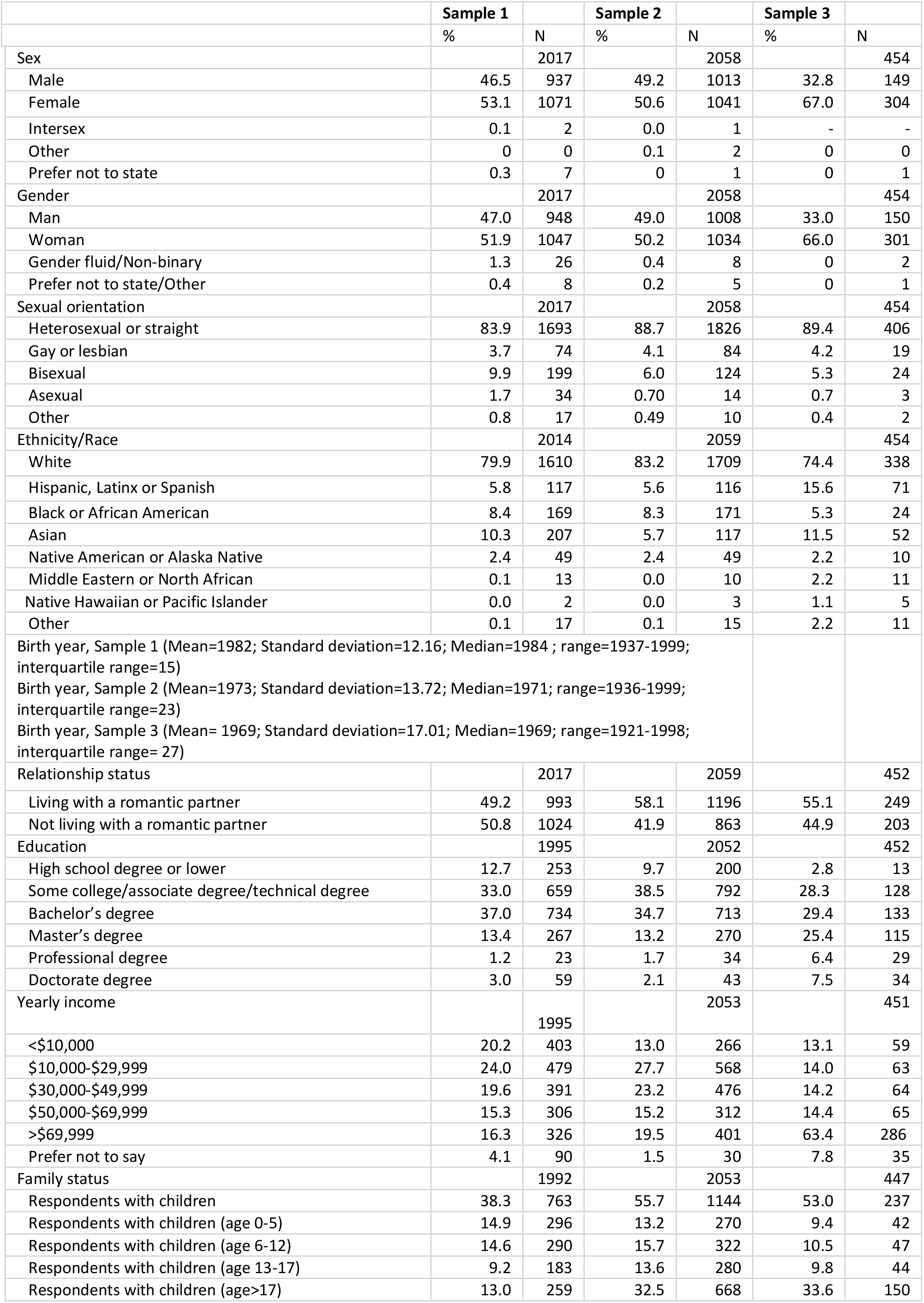
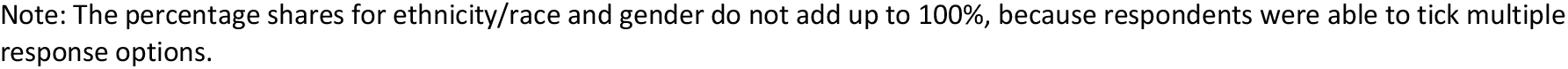
Sample Characteristics.

|  | Sample 1 |  | Sample 2 |  | Sample 3 |  |
| --- | --- | --- | --- | --- | --- | --- |
|  | % | N | % | N | % | N |
| Sex |  | 2017 |  | 2058 |  | 454 |
| Male | 46.5 | 937 | 49.2 | 1013 | 32.8 | 149 |
| Female | 53.1 | 1071 | 50.6 | 1041 | 67.0 | 304 |
| Intersex | 0.1 | 2 | 0.0 | 1 | - | - |
| Other | 0 | 0 | 0.1 | 2 | 0 | 0 |
| Prefer not to state | 0.3 | 7 | 0 | 1 | 0 | 1 |
| Gender |  | 2017 |  | 2058 |  | 454 |
| Man | 47.0 | 948 | 49.0 | 1008 | 33.0 | 150 |
| Woman | 51.9 | 1047 | 50.2 | 1034 | 66.0 | 301 |
| Gender fluid/Non-binary | 1.3 | 26 | 0.4 | 8 | 0 | 2 |
| Prefer not to state/Other | 0.4 | 8 | 0.2 | 5 | 0 | 1 |
| Sexual orientation |  | 2017 |  | 2058 |  | 454 |
| Heterosexual or straight | 83.9 | 1693 | 88.7 | 1826 | 89.4 | 406 |
| Gay or lesbian | 3.7 | 74 | 4.1 | 84 | 4.2 | 19 |
| Bisexual | 9.9 | 199 | 6.0 | 124 | 5.3 | 24 |
| Asexual | 1.7 | 34 | 0.70 | 14 | 0.7 | 3 |
| Other | 0.8 | 17 | 0.49 | 10 | 0.4 | 2 |
| Ethnicity/Race |  | 2014 |  | 2059 |  | 454 |
| White | 79.9 | 1610 | 83.2 | 1709 | 74.4 | 338 |
| Hispanic, Latinx or Spanish | 5.8 | 117 | 5.6 | 116 | 15.6 | 71 |
| Black or African American | 8.4 | 169 | 8.3 | 171 | 5.3 | 24 |
| Asian | 10.3 | 207 | 5.7 | 117 | 11.5 | 52 |
| Native American or Alaska Native | 2.4 | 49 | 2.4 | 49 | 2.2 | 10 |
| Middle Eastern or North African | 0.1 | 13 | 0.0 | 10 | 2.2 | 11 |
| Native Hawaiian or Pacific Islander | 0.0 | 2 | 0.0 | 3 | 1.1 | 5 |
| Other | 0.1 | 17 | 0.1 | 15 | 2.2 | 11 |
| Birth year, Sample 1 (Mean=1982; Standard deviation=12.16; Median=1984 ; range=1937-1999; interquartile range=15) |  |  |  |  |  |  |
| Birth year, Sample 2 (Mean=1973; Standard deviation=13.72; Median=1971; range=1936-1999; interquartile range=23) |  |  |  |  |  |  |
| Birth year, Sample 3 (Mean= 1969; Standard deviation=17.01; Median=1969; range=1921-1998; interquartile range= 27) |  |  |  |  |  |  |
| Relationship status |  | 2017 |  | 2059 |  | 452 |
| Living with a romantic partner | 49.2 | 993 | 58.1 | 1196 | 55.1 | 249 |
| Not living with a romantic partner | 50.8 | 1024 | 41.9 | 863 | 44.9 | 203 |
| Education |  | 1995 |  | 2052 |  | 452 |
| High school degree or lower | 12.7 | 253 | 9.7 | 200 | 2.8 | 13 |
| Some college/associate degree/technical degree | 33.0 | 659 | 38.5 | 792 | 28.3 | 128 |
| Bachelor's degree | 37.0 | 734 | 34.7 | 713 | 29.4 | 133 |
| Master's degree | 13.4 | 267 | 13.2 | 270 | 25.4 | 115 |
| Professional degree | 1.2 | 23 | 1.7 | 34 | 6.4 | 29 |
| Doctorate degree | 3.0 | 59 | 2.1 | 43 | 7.5 | 34 |
| Yearly income |  | 1995 |  | 2053 |  | 451 |
| <\$10,000 | 20.2 | 403 | 13.0 | 266 | 13.1 | 59 |
| \$10,000-\$29,999 | 24.0 | 479 | 27.7 | 568 | 14.0 | 63 |
| \$30,000-\$49,999 | 19.6 | 391 | 23.2 | 476 | 14.2 | 64 |
| \$50,000-\$69,999 | 15.3 | 306 | 15.2 | 312 | 14.4 | 65 |
| >\$69,999 | 16.3 | 326 | 19.5 | 401 | 63.4 | 286 |
| Prefer not to say | 4.1 | 90 | 1.5 | 30 | 7.8 | 35 |
| Family status |  | 1992 |  | 2053 |  | 447 |
| Respondents with children | 38.3 | 763 | 55.7 | 1144 | 53.0 | 237 |
| Respondents with children (age 0-5) | 14.9 | 296 | 13.2 | 270 | 9.4 | 42 |
| Respondents with children (age 6-12) | 14.6 | 290 | 15.7 | 322 | 10.5 | 47 |
| Respondents with children (age 13-17) | 9.2 | 183 | 13.6 | 280 | 9.8 | 44 |
| Respondents with children (age>17) | 13.0 | 259 | 32.5 | 668 | 33.6 | 150 |

Note: The percentage shares for ethnicity/race and gender do not add up to 100%, because respondents were able to tick multiple response options.

### Procedures

Self-rated health was assessed using the Health-related Quality of Life Core Module (CDC HRQoL-4). This module consists of four items about perceived general health, recent physical health, recent mental health, and recent activity limitations. The ordinal question about perceived general health did not meet the assumption of proportionality of odds required for ordered logistic regressions. Hence, we dichotomized the item into (1) fair or poor and (2) good, very good or excellent. We measured current smoking and current vaping by number of cigarettes smoked per day and number of times vaping per day. These variables were dichotomized in the analysis (not smoking= 0, smoking= 1; not vaping= 0, vaping= 1) due to a high frequency of zero values (>75%). Binge drinking was measured by the frequency of consuming five or more drinks on one occasion for males and four or more drinks on one occasion for females (within the last three months) (67). We followed standard procedure and recoded these items into a unisex dichotomous variable (binge drinking less than monthly= 0, binge drinking monthly, weekly, or daily/almost daily= 1). BMI was calculated based on self-reported height and weight and dichotomized for analysis to reflect under or normal weight (BMI<25= 0) and overweight or obese (BMI≥25= 1) (item phrasing and response options for the health questions are reported in Table S31). Specifications on the nine demographic covariates (including item phrasing and response options) used in the regressions are presented in Table S32).

### Exploratory factor analysis

We opted to start our analysis with EFA, rather than CFA, because we considered EFA to be the most appropriate first step for a survey measure of this novelty. While the systematic review allowed us to distill core gender-related attitudes and behaviors, we were uncertain how many latent factors would emerge in the subsequent testing. Moreover, we were uncertain how several of the items would distribute across factors (e.g. the items for time-use and quality in family relationships) and we wanted to leave open the possibility that various items would cross-load onto factors other than their parent factor, potentially leading to fewer latent factors than initially expected. As described in the results section, this happened to be the case. In addition, since we had the opportunity and resources to collect three survey samples, a complementary approach combining EFA and CFA allowed us to benefit from advantages of each method.

The EFA was based on iterated principal axis factoring as the extraction method and Promax (oblique) rotation to allow for correlated factors. Exploratory factor analysis and statistical analyzes were done in SPSS. In the EFA, we examined questions from all three gender categories in a common factor model with multiple factors. All respondents with missing data for relevant items were removed from sample 1 prior to the analysis. To allow for analysis of the largest possible sample, 10 items targeting caregivers and employees were recoded so that people not currently caring for someone in need or not currently employed (or employed in the past) were ascribed the value 1, which represents no strain due to caregiving or work (see Table S33).

We subjected the 44 questionnaire items to EFA in sample 1. Velicer’s minimum average partial test suggested a 7-factor structure (Table S34, screeplot, communalities, and unique variances are presented in Figure S2 and Table S35) (68). The conceptual clarity of this solution also best resembled the thematic gender dimensions identified in the literature review. This solution retained 35 of the 44 items subjected to EFA and explained 51% of the variance in item scores. For purposes of interpretability, we excluded all items with loadings below 0.40.

Factor one in this solution includes six items addressing perceived discrimination. Factor two consists of seven items capturing daily time spent on work and work strain-related characteristics. Factor three encompasses four items concerning perceived strain and time-use related to caregiving. Factor four includes five items capturing competitive and risk-taking behavior. Factor five encompasses three items concerning perceived social support. Factor six includes six items capturing empathy and expressive behavior. Finally, factor seven includes four questions about independence. The distribution of items on factors was consistent across alternative factor rotation methods (Tables S36-S39).

### Confirmatory factor analysis

The CFA was carried out in SPSS AMOS Graphics 26 and based on Maximum Likelihood estimations. We allowed the factors to be correlated. The Likelihood-Ratio test (also known as the *χ*^*2*^-test) is highly sensitive to even small departures of the data from exact fit, especially in large-N samples. Moreover, *χ*^*2*^ values increase with sample size and the number of variables in the model (69). Therefore, we followed Cheung and Rensvold (70) and Yuan and Bentler (71) and determined global model fit and invariance based on the approximate-fit statistics.

Specifically, we used the Tuckler Lewis Index (TLI), the root mean square error of approximation (RMSEA), and the standardized root mean square residual (SRMR), relying on conventional fit criteria (72). The 35 items and 7 factors retained in the EFA were submitted to CFA in samples 1, 2, and 3. Following Gerbing and Hamilton (73), we used CFA to refine the EFA solution identified in sample 1. Next, we cross-validated the outcomes of this CFA in samples 2 and 3. Configural invariance was examined separately in samples 2 and 3. We used Multiple-groups CFA to assess metric and scalar invariance across samples 2 and 3. We removed all observations with missing data for one or several of the 35 items subjected to CFA in sample 1, and for the 25 items subjected to CFA in samples 2 and 3. This slightly reduced sample 1 from n= 2,051 to n= 2,009, sample 2 from n= 2,135 to n= 2,054, and sample 3 from n=489 to n=449. One item concerning perceived gender discrimination in education had high rates of missing data in sample 3 (N>100). Hence, we restricted the cross-validation in sample 3, and the multiple-groups CFA in samples 2 and 3, to the remaining 24 variables. To avoid a Heywood estimate on the Social support factor, we followed Chen et al. (74) and constrained the error variance of one item (socsupchores) to 0.001 in samples 2 and 3.

The initial 35-item solution based on the EFA did not perform satisfactorily with respect to global model fit in sample 1 (χ^2^= 5897.38, df= 539, p= 0.00, TLI= 0.81, RMSEA= 0.07, SRMR= 0.07). To obtain acceptable model fit, we examined the factor loadings, removing all items with loadings ≤ 0.5. The “trimmed” solution, consisting of 25 items, exhibited good fit to the data (χ^2^= 1362.53, df= 254, p= 0.00, TLI= 0.95, RMSEA= 0.05, SRMR= 0.04) (Table S40). The cross-validation of this final model in samples 2 and 3, with no equality constraints, also exhibited reasonable fit to the data, indicating configural invariance (sample 2: χ^2^= 1440.7, df= 255, p= 0.00, TLI= 0.94, RMSEA= 0.05, SRMR= 0.04; sample 3: χ^2^= 496.5, df= 232, p= 0.00,TLI= 0.93, RMSEA=0.05, 3: SRMR=0.05) (Table S41).

A more restricted multiple-groups CFA with factor loadings assumed to be equal across samples 2 and 3 also supported metric invariance (χ^2^= 1899.849, df= 481, p= 0.00, TLI= 0.94, RMSEA= 0.03, SRMR= 0.04) (Table S42). Further restrictions with both factor loadings and intercepts assumed to be equal across samples 2 and 3 also indicated reasonable fit compared to prior models, supporting scalar invariance (χ^2^= 2167.051, df= 505, p= 0.00, TLI= 0.93, RMSEA= 0.04, SRMR= 0.04) (Table S43). As a sensitivity check, we also ran the multiple groups CFA in samples 2 and 3 with the item on perceived gender discrimination in education included (sample 2= 2,054; sample 3= 348) and obtained comparable model fit (metric invariance: χ^2^= 2149.723, df= 528, p= 0.00, TLI= 0.93, RMSEA= 0.04, SRMR= 0.04; scalar invariance: χ^2^= 2380.150, df= 553, p= 0.00, TLI= 0.93, RMSEA= 0.04, SRMR= 0.04) (Tables S44-S45).

## Reliability

The reliabilities of the factors implied by the final CFA solution were assessed in all samples using Raykov’s ρ. ρ was computed using James Gaskin’s ‘Validity master tool’ (75), and following conventional criteria (76), we considered values >0.60 desirable.

## Data Availability

Data and code needed to evaluate the conclusions are available here: https://osf.io/7yje9/

## Data and materials availability

Data and code needed to evaluate the conclusions are available here: **https://osf.io/7yje9/**.

## Acknowledgments

Stanford Humanities Center; Stanford School of Humanities and Science, Stanford University funded this research. The funder had no involvement in this study.

## Supplementary Information for

Gender-Related Variables for Health Research

## Supplementary Information Text

### Materials

#### Questionnaire: The Stanford Gender-Related Variables for Health Research

Please specify your current employment status:

▢ Employed Full-time
▢ Employed Part-time
▢ Homemaker
▢ Retired
▢ Student
▢ Unemployed

People see themselves in different ways. There are no right or wrong answers to the following questions. We just want to know what’s true for *you*. Choose the answer that best describes you.

i. In general, how prepared are you to take risks?
  ◯ Not at all prepared
  ◯ Slightly prepared
  ◯ Moderately prepared
  ◯ Very prepared
  ◯ Completely prepared
ii. How prepared are you to take risks….

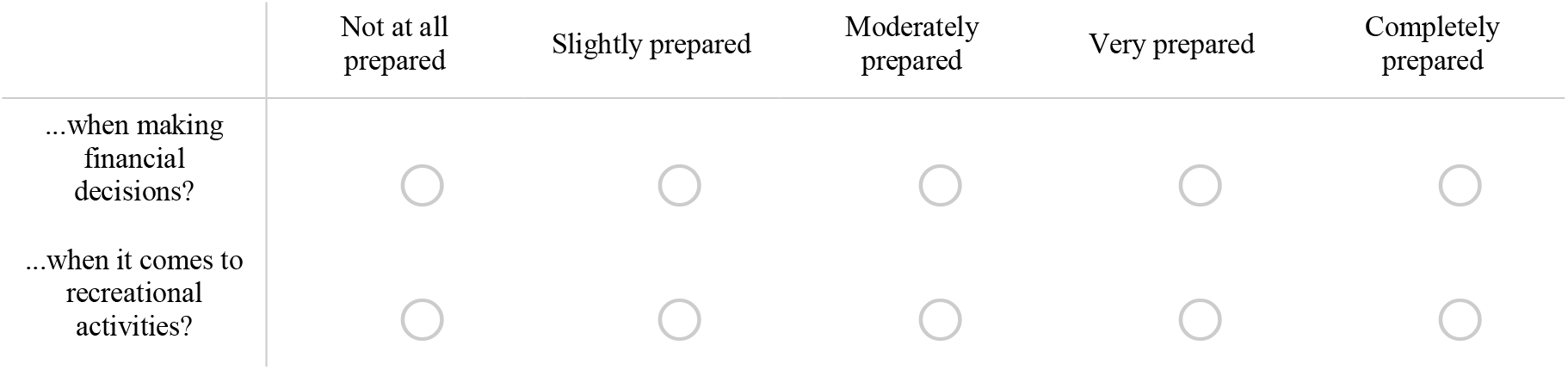 People see themselves in different ways. There are no right or wrong answers to the following questions. We just want to know what’s true for *you*. Choose the answer that best describes you.

i. How important is it for you to be independent?
  ◯ Extremely important
  ◯ Very important
  ◯ Moderately important
  ◯ Slightly important
  ◯ Not at all important

How important is it for you to solve your problems on your own?

◯ Extremely important
◯ Very important
◯ Moderately important
◯ Slightly important
◯ Not at all important

People see themselves in different ways. There are no right or wrong answers to the following questions. We just want to know what’s true for *you*. Choose the answer that best describes you.

iii. How often do friends talk to you about their problems?
  ◯ Never
  ◯ Once in a while
  ◯ Sometimes
  ◯ Most of the time
  ◯ Always
  People see themselves in different ways. There are no right or wrong answers to the following questions. We just want to know what’s true for *you*. Choose the answer that best describes you.

i. How often do you talk to your friends about your problems?
  ◯ Never
  ◯ Once in a while
  ◯ Sometimes
  ◯ Most of the time
  ◯ Always
ii. How easy is it for you to express what you are feeling to others?
  ◯ Not at all easy
  ◯ Slightly easy
  ◯ Moderately easy
  ◯ Very easy
  ◯ Extremely easy

Are you currently responsible for taking care of someone in need? By “taking care of someone in need” we mean providing unpaid assistance and support to someone who has physical or psychological needs, such as a child, elder, partner, or disabled family member.

◯ I am currently responsible for taking care of someone in need
◯ I have been responsible for taking care of someone in the past
◯ I have never been responsible for taking care of someone in need
◯ I am currently responsible for taking care of someone in need, and I have been responsible for taking care of someone in the past

*Display This Question*

*If Are you currently responsible for taking care of someone in need? By “taking care of someone in n*… *!*= *I have never been responsible for taking care of someone in need*

In the past year, how often did you feel **emotionally** exhausted because of your caretaking responsibilities?

◯ Never
◯ Once in a while
◯ Sometimes
◯ Most of the time
◯ Always

In the past year, how often did you feel **physically** exhausted because of your caretaking responsibilities?

◯ Never
◯ Once in a while
◯ Sometimes
◯ Most of the time
◯ Always

In the past year, how often have your caretaking responsibilities caused you to worry about the future?

◯ Never
◯ Once in a while
◯ Sometimes
◯ Most of the time
◯ Always

We are interested in how you spend your time on an average weekday, Monday through Friday. Please provide your best estimate. On average, how many hours per weekday do you spend on the following.

Work (paid work, studying, internships, etc.):____________

Taking care of someone in need (caring for children, elders, partners in need, etc.):____________

We are interested in how you feel about your current job, including your daily work activities as an employee or student. For each of the following questions, select the answer that best describes your work activities. If you have several jobs, please think about the job that you spend most hours doing per week.

i. How often does your job require working fast?
  ◯ Never
  ◯ Once in a while
  ◯ Sometimes
  ◯ Most of the time
  ◯ Always
ii. How often does your job involve repetitive tasks?
  ◯ Never
  ◯ Once in a while
  ◯ Sometimes
  ◯ Most of the time
  ◯ Always
iii. How often do you feel **emotionally** exhausted from your work activities?
  ◯ Never
  ◯ Once in a while
  ◯ Sometimes
  ◯ Most of the time
  ◯ Always
iv. How often do you feel **physically** exhausted from your work activities?
  ◯ Never
  ◯ Once in a while
  ◯ Sometimes
  ◯ Most of the time
  ◯ Always
  People sometimes look to others for companionship, assistance, or other types of physical or emotional support. The following questions ask you about the support available to you when you need it. Choose the answer that best describes your situation.

i. In the past year, how often did you have someone…

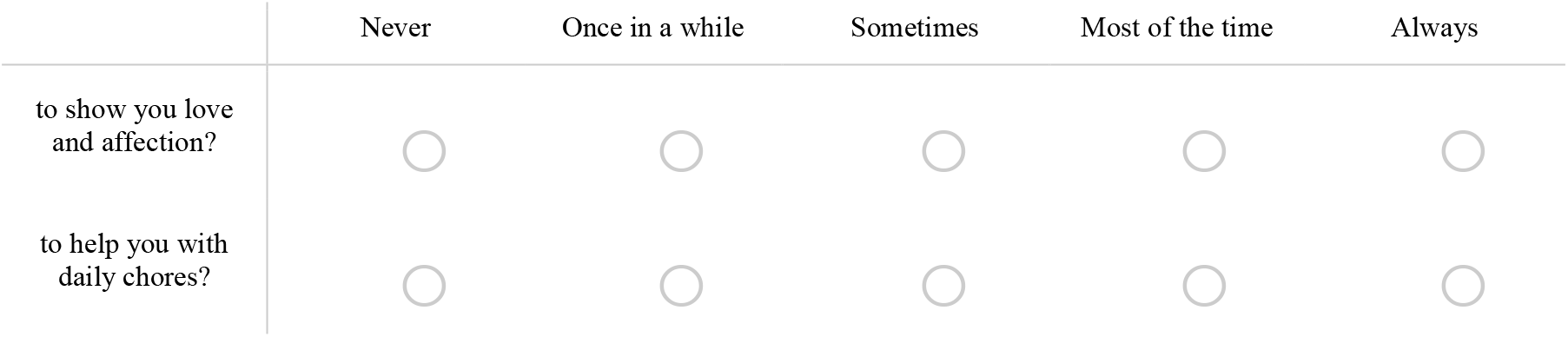 The following questions ask about how often you have felt discriminated against because of your gender. You may not be certain about your answers to these questions, but we would like you to choose the answer that best describes your experience.

i. Because of your gender, how often have you felt discriminated against?
  ◯ Never
  ◯ Once in a while
  ◯ Sometimes
  ◯ Most of the time
  ◯ Always
ii. Because of your gender how often have you felt discriminated against…

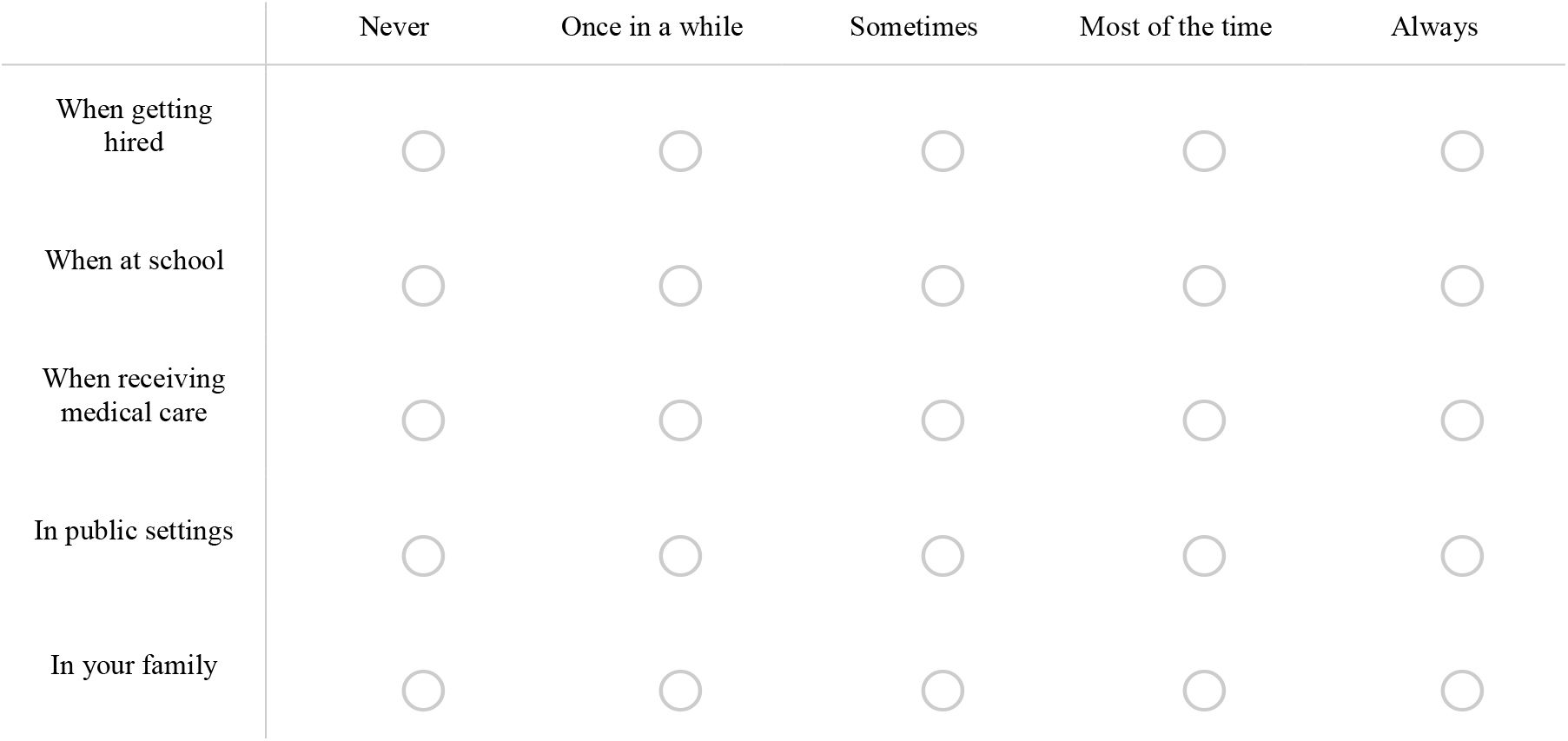

What was your birth sex?

◯ Male
◯ Female
◯ Intersex
◯ Other: Please specify:_____________________________
◯ Prefer not to state

What is your gender? Please select all that apply.

▢ Man
▢ Woman
▢ Gender fluid/Non-binary
▢ Other. Please specify:_____________________________
▢ Prefer not to state

In what year were you born?

▼ 2000 … 1900

Which categories describe you? (You may choose more than one.)

▢ White
▢ Hispanic, Latino, or Spanish
▢ Black or African American
▢ Asian
▢ Native American or Alaska Native
▢ Middle Eastern or North African
▢ Native Hawaiian or Pacific Islander
▢ Other

What was your income last calendar year?

Please combine all incomes. “Incomes” include wages, salaries, small business earnings, social security, armed forces pay, special cash bonuses and subsistence allowances.

◯ Less than $10,000
◯ $10,000 - $19,999
◯ $20,000 - $29,999
◯ $30,000 - $39,999
◯ $40,000 - $49,999
◯ $50,000 - $59,999
◯ $60,000 - $69,999
◯ $70,000 - $79,999
◯ $80,000 - $89,999
◯ $90,000 - $99,999
◯ $100,000 - $149,999
◯ $150,000 - $199,999
◯ More than $200,000
◯ Prefer not to say

What is the highest degree or level of school you have completed? If currently enrolled, please report the highest degree received.

◯ No schooling completed
◯ Preschool to 8th grade
◯ Some high school, no diploma
◯ High school graduate, diploma or equivalent (GED)
◯ Some college credit, no degree
◯ Trade/technical/vocational training
◯ Associate degree
◯ Bachelor’s degree
◯ Master’s degree
◯ Professional degree
◯ Doctorate degree What is your height?
◯ Feet_________________________________________
◯ Inches_________________________________________ What is your weight (in lbs)? How many cigarettes do you smoke per day? *Not including e-cigarettes*.
◯ Average #:_________________________________________ How many times do you vape or use an e-cigarette product per day?
________________________________________

*Display This Question:*

*If What was your birth sex?* = *Male*

In the past 3 months, how often have you had 5 or more drinks on one occasion?

◯ Never
◯ Less than monthly
◯ Monthly
◯ Weekly
◯ Daily or almost daily

*Display This Question*

*If What was your birth sex?* = *Female*

*In the past 3 months, how often have you had 4 or more drinks on one occasion?*

◯ Never
◯ Less than monthly
◯ Monthly
◯ Weekly
◯ Daily or almost daily

In general, would you say your health is…

◯ Excellent
◯ Very good
◯ Good
◯ Fair
◯ Poor

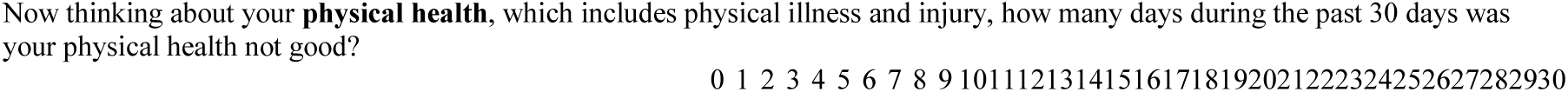
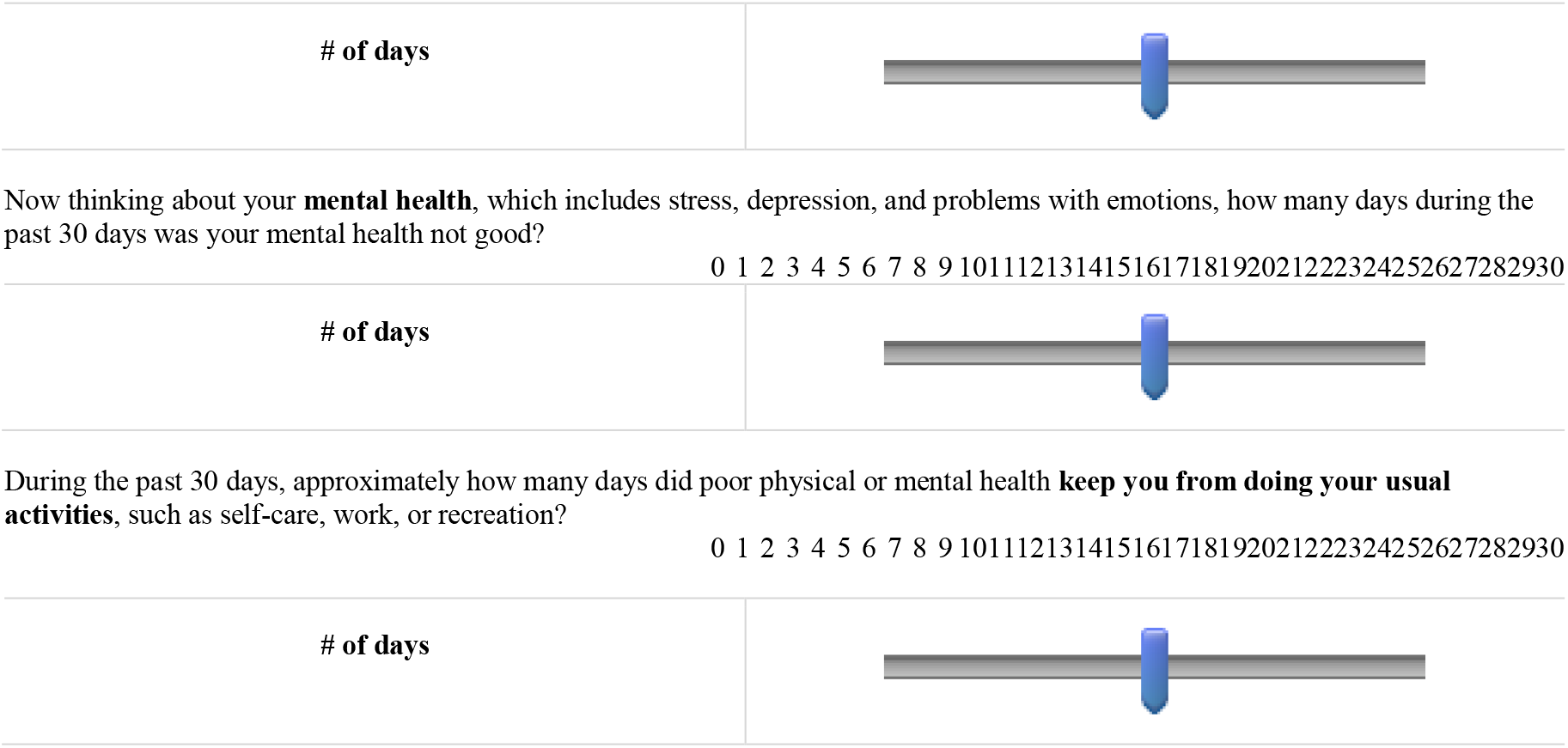

## Methods: Documentation of Literature survey

Here we document the systematic literature search that guided our development of the gender variables. The description of the literature survey is divided into five steps. Step 1 specifies the test-specific literature searches in PsycInfo, PsycNet, and PubMed with respect to database specific search limitations, search terms and selection criteria. Step 2 lists all of the gender measures derived from the articles identified through the searches in PsycInfo, PsycNet and PubMed. In Step 3, we sort the identified gender measures into three overarching categories for analyzing gender (norms, identity and relations), and use citation frequencies to determine the prevalence and use of each scale in the literature. In Step 4, we identify the core traits and characteristics of relevance to each of the three categories of gender-related traits, gender norms, and gender relations. Finally, Step 5 documents the search strategy used to identify additional construct specific measures of relevance to our gender variables.

### Step 1: Test-specific literature search in PsycINFO, PsycNET, and PubMed

This section specifies our search strategy. We carried out systematic searches in PsychINFO, PsycNET, and PubMED. We restricted the time span to the period from January 1975 through 2015.

### Search limitations

We used the following database-specific search limitations:

#### PsycINFO

Document type: Research literature in peer reviewed scholarly journals; Language: English; Population: Human; Classification: **“**Personality Scales & Inventories”; “Tests & Testing”.

#### PsycNet

Age-group: Adulthood (18 years or older); Document type: Abstract collection, Peer-reviewed articles; PsycNET Classifiers: **“**Tests and testing”; “Personality Scales & Inventories”; “Clinical Psychological Testing”; “Health Psychology Testing”; “Educational measurement”; “Occupational and employment testing”; “Consumer opinion and attitudes”.

#### PUBMED

Ages: all ages; Species: Humans; Language: English; Article types: all.

1. The search was narrowed to the following gender/sex related MeSH-terms: “gender identity”; “sex”; “sexism”; “interpersonal relations”; “female”, “male”. We excluded: “sexual and gender disorders”; “gender dysphoria”; “sex reassignment surgery”; “sex reassignment procedures.
2. We narrowed our search to the following “test-related” MeSH terms: “psychological tests”; “neuropsychological tests”; “personality tests”; “psychometrics”; “pain measurement”; ‘behavior rating scale”; “thematic apperception test”; “test anxiety scale”; “personal construct theory”; “Wechsler scales”; “personality assessment”.

### Search strings

We used the following search strings in the three databases:

#### PsycINFO

(“Gender Identity” OR “Gender Gap” OR “Gender Equality” OR “Sex Role Attitudes” OR “Sex Roles”)

#### PsycNet

(“Gender” OR “Sex”) NOT (“sexual”)

#### PubMed

(Psychological tests[mh] OR Neuropsychological tests[mh] OR Personality tests[mh] OR Psychometrics[mh] OR Pain measurement[mh] OR Behavior rating scale[mh] OR Thematic Apperception Test[mh] OR Test anxiety scale[mh] OR Personal Construct Theory[mh] OR Wechsler Scales[mh] OR Personality Assessment[mh]) AND (Sex[mh] OR Gender identity[mh] OR Interpersonal relations[mh] OR Sexism[mh] OR women[mh] OR men[mh] or female [mh] OR male [mh]) AND (scale*[ti] OR measure*[ti] OR compos*[ti] OR inventor*[ti] OR index*[ti] OR indic*[ti] OR score*[ti] OR psychomet*[ti] OR instrument*[ti] OR batter*[ti] OR assess*[ti] OR test*[ti] OR rating*[ti]) AND (Gender*[ti] OR sex[ti] OR sexes[ti] OR mascu*[ti] OR femin*[ti] OR male[ti] OR males[ti] OR female*[ti] OR man[ti] OR woman[ti] OR men[ti]] OR women[ti] OR androgy*[ti] OR boy[ti] OR girl[ti] OR boys[ti] OR girls[ti]).

The literature on gender- and sex-related scales is extremely extensive and for reasons of feasibility, it will not be possible to capture everything through a broad search in PubMed.

An important caveat related to our selected search string in PubMed is the emphasis on gender/ sex related search terms in abstract titles. To provide a concrete example: Our search string may not capture potentially relevant scales that do not include the following terms in their titles: (Gender*[ti] OR sex[ti] OR sexes[ti] OR mascu*[ti] OR femin*[ti] OR male[ti] OR males[ti] OR female*[ti] OR man[ti] OR woman[ti] OR men[ti]] OR women[ti] OR androgy*[ti] OR boy[ti] OR girl[ti] OR boys[ti] OR girls[ti]). However, for influential scales we will likely capture several follow up studies adapting or testing the validity of these scales. Test of coverage is captured in a later step.

The search strings presented above returned 519 studies in PsycINFO, 432 studies in PsycNET and 2030 studies in PubMed (Fig. S1).

### Selection criteria

We read through abstract and titles and used the following selection criteria to identify relevant publications:

All studies with scales or tests measuring aspects of gender are eligible, regardless of purpose of use in article (+); articles using non-gender related measures to illuminate gender differences on a given trait or topic (-); articles focusing on children or adolescents (17 years old or younger) (-); articles capturing scales measuring issues of gender norms or relations (e.g. child-rearing, romantic relationships, division of domestic labor) (+); articles capturing scales concerning sexuality, sexual health, sexual violence, domestic violence and issues of gender- and sex development in childhood and adolescence (-) and body image (-).

### Eligble publications

Using these, we identified 405 eligible publications: 315 in PsycINFO, 65 in PsychNET and 89 in PubMed. Of these, 64 publications were dublicates or triplicates (Fig. S1)

### Step 2: Identifying measures of gender

As a second step in the review process, we searched through all of the 405 eligible references to identify relevant gender measures employed in each paper. We derived 127 unique gender measures through this approach. Here, we list all of the 127 gender measures and the publications in which they were used. Four additional gender measures were identified through literature reviews published in books. These additional measures are reported at the bottom of this section. The gender measures are listed in alphabetized order. Adherence to Extreme Gender Role Beliefs:

## Step 3: Sorting and determining the impact of gender measures

As a third step in the review process, we sorted the 131 gender measures into three overarching categories for analyzing gender (norms, identity and relations). Given the large number of gender measures, we decided to use citation frequencies to determine the prevalence and use of each measure in the literature. All measures receiving at least 20 citations in Google Scholar within the last ten years were selected for further investigation, of which 74 scales met the criteria. All articles published from 2006-2015 were retained for further investigation. All articles published in 2005 or earlier, with less than 20 citations in Google Scholar were assorted into the “Ineligible Scales” category. This category also comprises measures that we found irrelevant after reading through abstract and full text. The scales assorted into each category (norms, identity and relations) are ranked based on their citation impact. Ineligible scales are listed at the bottom of this section.

## Gender norms (N=3)

**Coparenting and Family Rating System**

**Original scale:** McHale, J. P., Kuersten-Hogan, R., & Lauretti, A. (2000). Evaluating coparenting and family-level dynamics during infancy and early childhood: The Coparenting and Family Rating System. *Family observational coding systems: Resources for systemic research*, 151-170.

Web of Science (2006->): Missing

Google scholar (2006->): 57

**Parental Child-rearing Behaviour Scale**

**Original scale:** Meunier, J. C., & Roskam, I. (2007). Psychometric properties of a parental childrearing behavior scale for French-speaking parents, children, and adolescents. *European Journal of Psychological Assessment, 23*(2), 113-124.

Web of Science (2006->): 17 (full database), 3 (health-related subject categories)

Google scholar (2006->): 29

**The Orientation towards Domestic Labor**

**Original scale:** Hawkins, A. J., Marshall, C. M., & Allen, S. M. (1998). The Orientation Toward Domestic Labor Questionnaire: Exploring dual-earner wives’ sense of fairness about family work. *Journal of Family Psychology, 12*(2), 244.

Web of Science (2006->): 7 (full database)

Google scholar (2006->): 28

## Gender-related traits (N=31)

**Bem Sex Role Inventory**

**Original scale:** Bem, S. L. (1974). The measurement of psychological androgyny. *Journal of consulting and clinical psychology, 42*, 155-162 Web of Science (2006->): 802 (full database), 100 (health research) Google scholar (2006->): 3670

**Minnesota Multiphasic Personality Inventory (broader, including masc. and fem.)**

**Original scale:** Hathaway, S. R., & McKinley, J. C. (1951). Minnesota Multiphasic Personality Inventory; manual (Revised).

Web of Science (2006->): Missing

Google scholar (2006->): 937

**Conformity to Masculine Norms Inventory**

**Original scale:** Mahalik, J. R., Locke, B. D., Ludlow, L. H., Diemer, M. A., Scott, R. P., Gottfried, M., & Freitas, G. (2003). Development of the Conformity to Masculine Norms Inventory. *Psychology of Men & Masculinity, 4*(1), 3.

Web of Science (2006->) Missing

Google scholar (2006->): 676

**Gender Role Conflict Scale**

**Original scale:** O’Neil, J. M., Helms, B. J., Gable, R. K., David, L., & Wrightsman, L. S. (1986). Gender-Role Conflict Scale: College men’s fear of femininity. *Sex roles,14*(5-6), 335-350.

Web of Science (2006->): 185 (full database), 20 (health research)

Google scholar (2006->): 576

**Personal Attributes Questionnaire**

**Original scale:** Spence, J. T., Helmreich, R. L., & Stapp, J. (1974). The Personal Attributes Questionnaire: A measure of sex role stereotypes and masculinity-femininity.

Web of Science (2006->): Missing

Google scholar (2006->): 428

**Male Role Norms Scale**

**Original scale:** Thompson, E. H., & Pleck, J. H. (1986). The structure of male role norms. *The American Behavioral Scientist, 29*(5), 531.

Web of Science (2006->): 142 (full database) 15 (health)

Google scholar (2006->): 368

**Masculinity Ideology**

**Original scale:** Pleck, J. H., Sonenstein, F. L., & Ku, L. C. (1993). Masculinity ideology: Its impact on adolescent males’ heterosexual relationships. *Journal of Social issues, 49*(3), 11-29.

Web of Science (2006->): 131 (full database), 69 (health research)

Google scholar (2006->): 312

**Californian Psychological Inventory**

**Original scale:** Gough, H. G. (1975). *Manual for the California psychological inventory*. Consulting Psychologists Press.

Web of Science (2006->): Missing

Google scholar (2006->): 314

**Masculine Gender Role Stress**

**Original scale:** Eisler, R. M., & Skidmore, J. R. (1987). Masculine gender role stress scale development and component factors in the appraisal of stressful situations. *Behavior Modification, 11*(2), 123-136.

Web of Science (2006->): 100 (full database), 19 (health)

Google scholar (2006->): 301

**Traditional Machismo and Caballerismo Scale**

**Original scale:** Arciniega, G. M., Anderson, T. C., Tovar-Blank, Z. G., & Tracey, T. J. (2008). Toward a fuller conception of Machismo: Development of a traditional Machismo and Caballerismo Scale. *Journal of Counseling Psychology, 55*(1), 19.

Web of Science (2006->): 100 (full database), 33 (health research)

Google scholar (2006->): 276

**Conformity to Feminine Norms Inventory**

**Original scale:** Mahalik, J. R., Morray, E. B., Coonerty-Femiano, A., Ludlow, L. H., Slattery, S. M., & Smiler, A. (2005). Development of the conformity to feminine norms inventory. *Sex Roles, 52*(7-8), 417-435.

Web of Science (2006->): 82 (full database), 17 (health research)

Google scholar (2006->): 241

**Male Role Norms Inventory**

**Original scale:** Levant, R. F., Hirsch, L. S., Celentano, E., & Cozza, T. M. (1992). The male role: An investigation of contemporary norms. *Journal of Mental Health Counseling*.

Web of Science (2006->): Missing

Google scholar (2006->): 191

**Hypermasculinity Inventory**

**Original scale:** Mosher, D. L., & Sirkin, M. (1984). Measuring a macho personality constellation. *Journal of Research in Personality, 18*(2), 150-163.

Web of Science (2006=>): 83 (full database), 4 (health research)

Citations Google (2006=>): 251

**Brannon Masculinity Scale**

**Original scale:** Brannon, R., & Juni, S. (1984). A scale for measuring attitudes about masculinity. *Psychological Documents, 14*.

Web of Science (2006->): 2 (full database)

Google scholar (2006->): 141

**Feminine Gender Role Stress**

**Original scale:** Gillespie, B. L., & Eisler, R. M. (1992). Development of the feminine gender role stress scale: A cognitive-behavioral measure of stress, appraisal, and coping for women. *Behavior Modification, 16*(3), 426-438.

Web of Science (2006->): 31 (full database)

Google scholar (2006->): 78

**Hyperfemininity**

**Original scale:** Murnen, S. K., & Byrne, D. (1991). Hyperfeminity: Measurement and Initial Validation of the Construct. *The Journal of Sex Research*, 479-489.

Web of Science (2006->): 31 (full database) 4 (health)

Google scholar (2006->): 76

**Adjective Check List (Masculinity and Femininity scales)**

**Original scale:** Zuckerman, M. (1960). The development of an affect adjective check list for the measurement of anxiety. *Journal of Consulting Psychology, 24*(5), 457.

Web of Science (2006->): 14 (full database)

Google scholar (2006->): 72

**Normative Male Alexithymia Scale**

**Original scale:** Levant, R. F., Good, G. E., Cook, S. W., O’Neil, J. M., Smalley, K. B., Owen, K., & Richmond, K. (2006). The normative Male Alexithymia Scale: Measurement of a gender-linked syndrome. *Psychology of Men & Masculinity, 7*(4), 212.

Web of Science (2006->): Missing

Google scholar (2006->): 65

**Japanese Gender Role Index**

**Original scale:** Sugihara, Y., & Katsurada, E. (2002). Gender role development in Japanese culture: Diminishing gender role differences in a contemporary society. *Sex roles, 47*(9-10), 443-452.

Web of Science (2006->): 13 (full database) 2 (health)

Google scholar (2006->): 44

**Femininity Ideology Scale**

**Original scale:** Levant, R., Richmond, K., Cook, S., House, A. T., & Aupont, M. (2007). The femininity ideology scale: Factor structure, reliability, convergent and discriminant validity, and social contextual variation. *Sex Roles, 57*, 373–383.

Web of Science (2006=>): 15 (full database), 3 (health research)

Google scholar (2006=>): 42

**Gender Role Journey Measure**

**Original scale:** O’Neil, J. M., Egan, J., Owen, S. V., & Murry, V. M. (1993). The gender role journey measure: Scale development and psychometric evaluation. *Sex Roles,28*(3-4), 167-185.

Web of science (2006=>): 10 (full database), 1 (health research)

Google scholar (2006=>): 43

**Auburn Differential Masculinity Inventory**

**Original scale:** Burk, L. R., Burkhart, B. R., & Sikorski, J. F. (2004). Construction and Preliminary Validation of the Auburn Differential Masculinity Inventory. *Psychology of Men & Masculinity, 5*(1), 4.

Web of Science (2006->): Missing

Google scholar (2006=>): 36

**PRF Andro Scale (Personality Research Form)**

**Original scale:** Berzins, J. I., Welling, M. A., & Wetter, R. E. (1978). A new measure of psychological androgyny based on the Personality Research Form. *Journal of consulting and clinical psychology, 46*(1), 126.

Web of Science (2006->): 8 (full database), 1 (health research)

Google scholar (2006->): 33

**Sex-Role Identity Scale**

**Original scale:** Storms, M. D. (1979). Sex role identity and its relationships to sex role attributes and sex role stereotypes. *Journal of Personality and Social Psychology, 37*(10), 1779.

Web of science (2006=>): 15 (full database), 2 (health research)

Google scholar (2006=>): 27

**New Masculine Gender Role Discrepancy**

**Original scale:** Rummell, C. M., & Levant, R. F. (2014). Masculine gender role discrepancy strain and self-esteem. *Psychology of Men & Masculinity, 15*(4), 419.

Too new for citations

**Russian Male Norms Inventory**

**Original scale:** Janey, B. A., Kim, T., Jampolskaja, T., Khuda, A., Larionov, A., Maksimenko, A., … & Shipilova, A. (2013). Development of the Russian Male Norms Inventory. *Psychology of Men & Masculinity, 14*(2), 138.

Too new for citations

**Measurement of Men’s Perceived Inexpressiveness**

**Original scale:** Wong, Y. J., Horn, A. J., Gomory, A. M., & Ramos, E. (2013). Measure of Men’s Perceived Inexpressiveness Norms (M2PIN): Scale development and psychometric properties. *Psychology of Men & Masculinity, 14*(3), 288.

Too new for citations

**Positive Negative Sex-role Inventory**

**Original scale:** Berger, A., & Krahé, B. (2013). Negative attributes are gendered too: Conceptualizing and measuring positive and negative facets of sex-role identity. *European Journal of Social Psychology, 43*(6), 516-531.

Too new for citations

**Social Roles Questionnaire**

**Original scale:** Baber, K. M., & Tucker, C. J. (2006). The social roles questionnaire: A new approach to measuring attitudes toward gender. *Sex Roles, 54*(7-8), 459-467.

Too new for citations

**Indian Gender role identity**

Basu, J. (2010). Development of the Indian gender role identity scale. *Journal of the Indian Academy of Applied Psychology, 36*(1), 25-34.

Too new for citations

**Inventory of Subjective Masculinity**

Wong, Y. J., Shea, M., Lafollette, J. R., Hickman, S. J., Cruz, N., & Boghokian, T. (2011). The inventory of subjective masculinity experiences: Development and psychometric properties. *The Journal of Men’s Studies,19*(3), 236-255.

Too new for citations

## Gender Relations (N=40)

**The Ambivalent Sexism Inventory**

**Original scale:** Glick, P., & Fiske, S. T. (1996). The ambivalent sexism inventory: Differentiating hostile and benevolent sexism. *Journal of personality and social psychology, 70*(3), 491.

Web of Science (2006=>): 792 (full database), 41 (health research)

Google scholar (2006=>): 1960

**Modern and Old-fashioned Sexism**

**Original scale:** Swim, J. K., Aikin, K. J., Hall, W. S., & Hunter, B. A. (1995). Sexism and racism: Old-fashioned and modern prejudices. *Journal of Personality and Social Psychology, 68*, 199–214 Web of Science (2006=>):306 (full database), 12 (health research)

Google scholar (2006=>): 801

**Dyadic Adjustment Scale across Gender**

Original scale: Christensen, A., & Heavey, C. L. (1990). Gender and social structure in the demand/withdraw pattern of marital conflict. *Journal of personality and social psychology, 59*(1), 73.

Web of science (2006=>): 193 (full database), 10 (health research)

Google scholar (2006=>): 488

**Multidimensional Measure of Sexual Minority Identity**

**Original scale:** Mohr, J., & Fassinger, R. (2000). Measuring dimensions of lesbian and gay male experience. *Measurement and Evaluation in Counseling and Development, 33*(2), 66-66.

Web of science (2006=>): 123 (full database), 32 (health research)

Google scholar (2006=>): 346

**Relationship Assessment Scale**

**Original scale:** Hendrick, S. S., Dicke, A., & Hendrick, C. (1998). The relationship assessment scale. *Journal of Social and Personal Relationships, 15*(1), 137-142.

Web of science (2006=>): 124 (full database), 34 (health research)

Google scholar (2006=>): 337

**Neosexism Scale**

**Original scale:** Tougas, F., Brown, R., Beaton, A. M., & Joly, S. (1995). Neosexism: Plus ça change, plus c’est pareil. *Personality and Social Psychology Bulletin, 21*(8), 842-849.

Web of science (2006=>): 111 (full database), 1 (health research)

Google scholar (2006=>): 308

**Attitudes toward Women Scale**

**Original scale:** Spence, J. T., Helmreich, R., & Stapp, J. (1973). A short version of the Attitudes toward Women Scale (AWS). *Bulletin of the Psychonomic Society,2*(4), 219-220.

Web of Science (2006->): 104 (full database), 41 (health research)

Google scholar (2006->): 289

**Routine and Strategic Relational Maintenance Scale**

Stafford, L., Dainton, M., & Haas, S. (2000). Measuring routine and strategic relational maintenance: Scale revision, sex versus gender roles, and the prediction of relational characteristics. *Communications Monographs, 67*(3), 306-323.

Web of Science (2006->): Missing

Google scholar (2006->): 211

**Sex-Role Stereotypes**

**Original scale:** Rosenkrantz, P., Vogel, S., Bee, H., Broverman, I., & Broverman, D. M. (1968). Sex-role stereotypes and self-concepts in college students. Journal of consulting and clinical psychology, 32(3), 287.

Web of Science (2006->): 43 (full database),

Google scholar (2006->): 210

**Relationship Belief Inventory**

**Original scale:** Eidelson, R. J., & Epstein, N. (1982). Cognition and relationship maladjustment: Development of a measure of dysfunctional relationship beliefs. *Journal of consulting and clinical psychology, 50*(5), 715.

Web of science (2006=>): 46 (full database), 1 (health research)

Google scholar (2006=>): 196

**The Romantic Beliefs Scale**

**Original scale:** Sprecher, S., & Metts, S. (1989). Development of the Romantic Beliefs Scale and examination of the effects of gender and gender-role orientation.*Journal of Social and Personal Relationships, 6*(4), 387-411.

Web of science (2006=>): 26 (full database), 2 (health research)

Google scholar (2006=>): 133

**Relationship Authenticity**

**Original scale:** Lopez, F. G., & Rice, K. G. (2006). Preliminary development and validation of a measure of relationship authenticity. *Journal of Counseling Psychology, 53*(3), 362.

Web of science (2006=>): 53 (full database), 5 (health research)

Google scholar (2006=>): 123

**Sex-Role Egalitarianism Scale**

**Original scale:** Beere, C. A., King, D. W., Beere, D. B., & King, L. A. (1984). The Sex-Role Egalitarianism Scale: A measure of attitudes toward equality between the sexes. *Sex Roles, 10*(7-8), 563-576.

Web of science (2006=>): 41 (full database), 5 (health research)

Google scholar (2006=>): 122

**Quick Discrimination Index**

**Original scale:** Ponterotto, J. G., Burkard, A., Rieger, B. P., Grieger, I., D’Onofrio, A., Dubuisson, A., Heenehan, M., Millstein, B., Parisi, M., Rath, J. F., & Sax, G. (1995). Development and initial validation of the Quick Discrimination Index (QDI). Educational and Psychological Measurement, 55, 1026-1031.

Web of science (2006=>): 31 (full database), 2 (health research)

Google scholar (2006=>): 110

**Attitudes toward Traditional and Egalitarian Sex-Roles**

**Original scale:** Larsen, K. S., & Long, E. (1988). Attitudes toward sex-roles: Traditional or egalitarian?. Sex Roles, 19(1-2), 1-12.

Web of science (2006=>): 43 (full database), 2 (health research)

Google scholar (2006=>): 110

**Mutual Psychological Development Questionnaire**

**Original scale:** Genero, N. P., Miller, J. B., Surrey, J., & Baldwin, L. M. (1992). Measuring perceived mutuality in close relationships: Validation of the Mutual Psychological Development Questionnaire. *Journal of Family Psychology, 6*(1), 36.

Web of science (2006=>): Missing

Google scholar (2006=>): 103

**Swedish Classical and Modern Sexism Scales**

**Original scale:** Ekehammar, B., Akrami, N., & Araya, T. (2000). Development and validation of Swedish classical and modern sexism scales. *Scandinavian journal of psychology, 41*(4), 307-314. Web of science (2006=>): 19 (full database), 3 (health research)

Google scholar (2006=>): 65

**Sex-Role Ideology**

**Original scale:** Kalin, R., & Tilby, P. J. (1978). Development and validation of a sex-role ideology scale. *Psychological reports, 42*(3), 731-738.

Web of Science (2006->): 19 (full database), 2 (health research)

Google scholar (2006->): 59

**Quality in Relationships Inventory within Couples**

**Original scale:** Pierce, G. R. (1994). The Quality of Relationships Inventory: Assessing the interpersonal context of social support.

Web of science (2006=>): Missing

Google scholar (2006=>): 52

**Partner Behaviour Inventory**

**Original scale:** Doss, B. D., & Christensen, A. (2006). Acceptance in romantic relationships: the frequency and acceptability of partner behavior inventory. *Psychological Assessment, 18*(3), 289. Web of science (2006=>): 13 (full database), 4 (health research)

Google scholar (2006=>): 45

**Managing Affect and Differences Scale**

**Original scale:** Arellano, C. M., & Markman, H. J. (1995). The Managing Affect and Differences Scale (MADS): A self-report measure assessing conflict management in couples. *Journal of Family Psychology, 9*(3), 319

Web of science (2006=>): 8 (full database)

Google scholar (2006=>): 33

**Women as Managers Scale**

**Original scale:** Peters, L. H., Terborg, J. R., & Taynor, J. (1974). Women as Managers Scale:(WAMS): a Measure of Attitudes Toward Women in Management Positions. Journal Supplement Abstract Service of the American Psychological Association.

Web of science (2006=>): Missing

Google scholar (2006=>): 54

**Gender Role Egalitarian Attitudes**

**Original Scale:** Chang, L. (1999). Gender role egalitarian attitudes in Beijing, Hong Kong, Florida, and Michigan. *Journal of Cross-Cultural Psychology, 30*(6), 722-741.

Web of Science (2006->): 22 (full database) 1 (health)

Google scholar (2006->): 54

**Adherence to Extreme Gender Role Beliefs**

**Original scale:** Hamburger, M. E., Hogben, M., McGowan, S., & Dawson, L. J. (1996). Assessing Hypergender Ideologies: Development and Initial Validation of a Gender-Neutral Measure of Adherence to Extreme Gender-Role Beliefs. Journal of Research in Personality, 30(2), 157-178.

Web of Science (2006->): 23 (full database) 3 (health)

Google scholar (2006->): 45

**Gender Attitude Inventory**

**Original scale:** Ashmore, R. D., Del Boca, F. K., & Bilder, S. M. (1995). Construction and validation of the Gender Attitude Inventory, a structured inventory to assess multiple dimensions of gender attitudes. *Sex roles, 32*(11-12), 753-785.

Web of Science (2006->): 15 (full database)

Google scholar (2006->): 45

**Sex-Role Orientation**

**Original scale:** Brogan, D., & Kutner, N. G. (1976). Measuring sex-role orientation: A normative approach. *Journal of Marriage and the Family*, 31-40.

Web of science (2006=>): 12 (full database) 4 (health)

Google scholar (2006=>): 44

**Gender ideology as an identity**

**Original scale:** Kroska, A. (2000). Conceptualizing and measuring gender ideology as an identity. *Gender & Society, 14*(3), 368-394.

Web of Science (2006->): 12 (full database) 2 (health)

Google scholar (2006->): 36

**Gender Role Beliefs Scale**

**Original scale:** Kerr, P. S., & Holden, R. R. (1996). Development of the gender role beliefs scale (GRBS). Journal of Social Behavior and Personality, 11(5), 3.

Web of Science (2006->): 9 (full database)

Google scholar (2006->): 34

**Male Attitude Norms Inventory II**

**Original scale:** Luyt, R. (2005). The Male Attitude Norms Inventory-II A Measure of Masculinity Ideology in South Africa. *Men and Masculinities, 8*(2), 208-229.

Web of science (2006=>): Missing

Google scholar (2006=>): 36

**Sexist Attitudes toward Women**

**Original scale:** Benson, P. L., Institute, S., & Vincent, S. (1980). Development and validation of the sexist attitudes toward women stale (SATWS). Psychology of Women Quarterly, 5(2), 276-291. Web of science (2006=>): 8 (full database), 4 (health)

Google scholar (2006=>): 31

**Race and Gender-Specific Stress Measure for African-American Women**

**Original scale:** Jackson, F. M. (2005). The Development of a Race and Gender-Specific Stress Measure for African-American Women: Jackson, Hogue, Phillips Contextualized Stress Measure. *Ethnicity & disease, 15*(4), 594-600.

Web of science (2006=>): 16 (full database), 10 (health research)

Google scholar (2006=>): 26

**Egalitarian Sex Role Attitudes**

Suzuki, A. (1991). Egalitarian sex role attitudes: Scale development and comparison of American and Japanese women. Sex Roles, 24(5-6), 245-259.

Web of science (2006=>): 15 (full database), 4 (health research)

Google scholar (2006=>): 26

**Schedule of Sexist Events**

**Original scale:** Yoder, J. D., & McDonald, T. W. (1998). Measuring sexist discrimination in the workplace: Support for the validity of the schedule of sexist events. *Psychology of Women Quarterly, 22*, 487–491.

Google scholar (2006=>): 22

Web of Science (2006=>): 8 (full database) 1 (health research)

**German Relationship Assessment Scale**

**Original scale:** Dinkel, A., & Balck, F. (2005). An evaluation of the German relationship assessment scale. *Swiss Journal of Psychology, 64*(4), 259-263.

Web of science (2006=>): 6 (full database), 1 (health research)

Google scholar (2006=>): 22

**Concord-Index (measure of family relationships)**

**Original scale:** Lee, P. H., Stewart, S. M., Lun, V., Bond, M. H., Yu, X., & Lam, T. H. (2012). Validating the concord index as a measure of family relationships in China.*Journal of Family Psychology, 26*(6), 906.

Too new for citations

**Conflict Disengagement Inventory (CDI)**

**Original scale:** Sanford, K. (2014). A latent change score model of conflict resolution in couples Are negative behaviors bad, benign, or beneficial?. *Journal of Social and Personal Relationships*, 0265407513518156.

Too new for citations

**Gender Minority Stress and Resilience**

**Original scale:** Testa, R. J., Habarth, J., Peta, J., Balsam, K., & Bockting, W. (2015). Development of the Gender Minority Stress and Resilience Measure. *Psychology of Sexual Orientation and Gender Diversity, 2*(1), 65.

Too new for citations

**Gender-Based Attitudes Toward Marriage and Child Rearing**

**Original scale:** Adams, M., Coltrane, S., & Parke, R. D. (2007). Cross-ethnic applicability of the gender-based attitudes toward marriage and child rearing scales. *Sex roles,56*(5-6), 325-339.

Too new for citations

**Patriarchal Beliefs Scale**

**Original scale:** Yoon, E., Adams, K., Hogge, I., Bruner, J. P., Surya, S., & Bryant, F. B. (2015). Development and validation of the Patriarchal Beliefs Scale. *Journal of counseling psychology, 62*(2), 264.

Too new for citations

**African American Men’s Gendered Racism Stress Inventory**

**Original scale:** Schwing, A. E., Wong, Y. J., & Fann, M. D. (2013). Development and validation of the African American Men’s Gendered Racism Stress Inventory. *Psychology of Men & Masculinity, 14*(1), 16.

Too new for citations

## Ineligible Scales (N=57)

**NEO Personality Inventory (broader scale)**

**Original scale:** Costa, P. T., & McCrae, R. R. (1985). *The NEO personality inventory: Manual, form S and form R*. Psychological Assessment Resources.

Web of Science (2006->): Missing

Google scholar (2006->): 2200

**Justification:** Doesn’t measure gender.

**Conformity to Traditional Masculinity and Relationship Satisfaction**

**Original scale:** Burn, S. M., & Ward, A. Z. (2005). Men’s Conformity to Traditional Masculinity and Relationship Satisfaction. *Psychology of Men & Masculinity, 6*(4), 254.

Web of Science (2006->): Missing

Google scholar (2006->): 99

**Justification:** Its traits and dimensions are based on the Conformity to Masculine Norms scale. This scale is already included.

**Gender Diagnosticity**

**Original scale:** Lippa, R., & Connelly, S. (1990). Gender diagnosticity: A new Bayesian approach to gender-related individual differences. *Journal of Personality and Social Psychology, 59*(5), 1051. Web of Science (2006->): 44 (full database), 5 (health research)

Google scholar (2006->): 61

**Justification:** Measure used to predict not measure gender

**Gender Differences in Sexual Attitudes**

**Original scale:** Hendrick, S., Hendrick, C., Slapion-Foote, M. J., & Foote, F. H. (1985). Gender differences in sexual attitudes. *Journal of Personality and Social Psychology,48*(6), 1630.

Web of Science (2006->): 21 (full database), 9 (health research)

Google scholar (2006->):61

**Justification:** Focuses on sexuality, not gender.

**Masculine Gender Identity In females**

**Original scale:** Blanchard, R., & Freund, K. (1983). Measuring masculine gender identity in females. *Journal of Consulting and Clinical Psychology, 51*(2), 205.

Web of Science (2006->): 9 (full database) 5 (health)

Google scholar (2006->): 21

**Justification:** Specifically designed to measure transgender behavior. Items (behaviors and sentiments) do not capture gender related traits and characteristics.

**Male Assessment of Self-Objectification**

**Original scale:** Daniel, S., Bridges, S. K., & Martens, M. P. (2014). The development and validation of the Male Assessment of Self-Objectification (MASO). *Psychology of Men & Masculinity, 15*(1), 78.

Too new for citations

**Justification:** Measures Body-related aspects of gender (not deemed relevant for our measure)

**Masculine Body Ideal Distress Scale**

**Original scale:** Kimmel, S. B., & Mahalik, J. R. (2004). Measuring masculine body ideal distress: Development of a measure. *International Journal of Men’s Health,3*(1), 1.

Too new for citations

**Justification:** Measures Body-related aspects of gender (not deemed relevant for our measure)

**Subjective Masculinity stress**

**Original scale:** Wong, Y. J., Shea, M., Hickman, S. J., LaFollette, J. R., Cruz, N., & Boghokian, T. (2013). The Subjective Masculinity Stress Scale: Scale development and psychometric properties. *Psychology of Men & Masculinity,14*(2), 148.

Too new for citations

**Justification:** Does not rely on predefined gender characteristics and traits. Respondents define their own “gender stressors” and hereafter rate them.

**Men’s Perceived Inexpressiveness Norms**

Too new for citations

**Justification:** Measures expressivity in men, not gender.

**Reference Group Identity Dependence Scale**

**Original scale:** Wade, J. C., & Gelso, C. J. (1998). Reference Group Identity Dependence Scale A Measure of Male Identity. *The Counseling Psychologist, 26*(3), 384-412.

Web of Science (2006->): 11 (full database), 1 (health research)

Google scholar (2006->): 28

**Justification:** Does not address gender traits and characteristics

**A Gender-Based Measurement Invariance Study of the Sociocultural Attitudes toward Appearance Questionnaire**

Original scale

Wheeler, D. L., Vassar, M., & Hale, W. D. (2011). A gender-based measurement invariance study of the Sociocultural Attitudes Toward Appearance Questionnaire-3. *Body image, 8*(2), 168-172.

Too new for citations

**Justification:** Measures Body-related aspects of gender (not deemed relevant for our measure)

**LGBT Ally Identity Measure**

**Original scale:** Jones, K. N., Brewster, M. E., & Jones, J. A. (2014). The creation and validation of the LGBT Ally Identity Measure. *Psychology of Sexual Orientation and Gender Diversity, 1*(2), 181.

Too new for citations

**Justification:** Does not measure gender

**O’ Kelly Women’s Belief Scale**

**Original scale:** O’Kelly, M. (2011). Psychometric properties of the O’Kelly women’s belief scales. *Journal of Rational-Emotive & Cognitive-Behavior Therapy, 29*(3), 145-157.

Too new for citations

**Justification:** Does not address gender traits and characteristics

**Lesbian and Gay Identity Scale**

Too new for citations

**Justification:** Doesn’t measure gender

**Wiggins Interpersonal Behavior Circle**

**Original scale:** Wiggins, J. S. (1979). A psychological taxonomy of trait-descriptive terms: The interpersonal domain. *Journal of personality and social psychology, 37*(3), 395.

Too new for citations

**Justification:** Doesn’t measure gender

**Social Sex-Role Inventory (DSI)**

**Original scale:** Shively, M. G., & De Cecco, J. P. (1977). Components of sexual identity. *Journal of homosexuality, 3*(1), 41-48.

Web of Science (2006->): 23 (full database), 7 (health research)

Google scholar (2006->): 142

**Justification:** Doesn’t measure gender

**Masculine Gender Role Discrepancy**

**Original scale:** Pleck, J. H. (1995). The gender role strain paradigm: An update. Web of Science (2006->): Missing

Google scholar (2006->): 590

**Justification:** This is a theory, not a scale

**Omnibus Personality Inventory**

**Original scale:** Heist, P., & Yonge, G. D. (1968). *Omnibus personality inventory*. Psychological Corporation.

**Justification:** Fewer than 20 citations since 2006

**Women’s Role Strain Inventory**

**Original scale:** Lengacher, C. A. (1993). Development of a predictive model for role strain in registered nurses returning to school. *The Journal of nursing education, 32*(7), 301-308.

**Justification:** Fewer than 20 citations since 2006

**Australian Sex-Role Scale**

**Original scale:** Antill, J. K., Cunningham, J. D., Russell, G., & Thompson, N. L. (1981). An Australian sex-role scale. *Australian Journal of Psychology, 33*(2), 169-183.

**Justification:** Fewer than 20 citations since 2006

**Home Career Conflict Measure**

**Original scale:** Farmer, H. S. (1984). Development of a measure of home-career conflict related to career motivation in college women. *Sex Roles, 10*(9-10), 663-675.

**Justification:** Fewer than 20 citations since 2006

**Sex-Role Blending**

**Original scale:** Heilbrun, A. B. (1981). Gender differences in the functional linkage between androgyny, social cognition, and competence. *Journal of Personality and Social psychology, 41*(6), 1106.

**Justification:** Fewer than 20 citations since 2006

**Chinese Gender Role Stress**

**Original scale:** Tang, C. S. K., & Lau, B. H. B. (1995). The assessment of gender role stress for Chinese. *Sex Roles, 33*(7-8), 587-595.

**Justification:** Fewer than 20 citations since 2006

**Feminine Gender Identity Scale**

**Original scale**

Freund, K., Nagler, E., Langevin, R., Zajac, A., & Steiner, B. (1974). Measuring feminine gender identity in homosexual males. *Archives of Sexual Behavior,3*(3), 249-260.

**Justification:** Fewer than 20 citations since 2006

**Self and Peer-Related Scale of Female and Male Roles and Attributes**

**Original scale:** Chang, L., & McBride-Chang, C. (1997). Self-and peer-ratings of female and male roles and attributes. *The Journal of social psychology, 137*(4), 527-529.

**Justification:** Fewer than 20 citations since 2006

**Behavioral Self-Report of Femininity**

**Original scale:** Greene, K. S., & Gynther, M. D. (1994). Another femininity scale? *Psychological reports, 75*(1), 163-170.

**Justification:** Fewer than 20 citations since 2006

**The Femininity Study**

**Original scale:** Thorne, F. C. (1977). The measurement of femininity. *Journal of clinical psychology, 33*(S1), 5-10.

**Justification:** Fewer than 20 citations since 2006

**Undesirable Characteristics Scale**

**Original scale:** Socially-undesirable sex-correlated characteristics: implications for androgyny and adjustment. *Journal of Consulting and Clinical Psychology*, 45, 1185-1186.

**Justification:** Fewer than 20 citations since 2006

**Measure of Adult Stereotypic Sex-Role Concepts of Masculinity**

**Original scale:** Newman, R. C. (1976). Development and Standardization of Measures of Stereotypic Sex-Role Concepts and of Sex-Role Adoption in Adults. *Psychological reports, 39*(2), 623-630.

**Justification:** Fewer than 20 citations since 2006

**Michigan Gender Identity Test (MIGIT)**

**Original scale:** Dull, C. Y., Catford, J. C., Guiora, A. Z., Beit-Hallahmi, B., Paluszny, M., & Cooley, R. E. (1975). The Michigan gender identity test (MIGIT). *Comprehensive psychiatry, 16*(6), 581-592.

**Justification:** Fewer than 20 citations since 2006

**Sex Rep**

**Original scale:** Baldwin, A. C., Critelli, J. W., Stevens, L. C., & Russell, S. (1986). Androgyny and sex role measurement: A personal construct approach. *Journal of Personality and Social Psychology, 51*(5), 1081.

**Justification:** Fewer than 20 citations since 2006

**Attitudes toward Masculinity Transcendence Scale**

**Original scale:** Moreland, J., & Van Tuinen, M. (1978). The attitude toward masculinity transcendence scale. *Unpublished manuscript, Southern Illinois University at Carbondale*.

**Justification:** Fewer than 20 citations since 2006

**Scales for Investigation of the Dual-Career Family**

**Original scale:** Pendleton, B. F., Poloma, M. M., & Garland, T. N. (1980). Scales for investigation of the dual-career family. *Journal of Marriage and the Family*, 269-276.

**Justification:** Fewer than 20 citations since 2006

**Sex-Role Antecedents Scale**

**Original scale:** Mast, D. L., & Herron, W. G. (1986). The sex-role antecendents scales. *Perceptual and Motor Skills, 63*(1), 27-56.

**Justification:** Fewer than 20 citations since 2006

**Multi-dimensional Sex-role Inventory**

**Original scale:** Bernard, L. C., & Wood, J. (1990). Further observations on the multidimensional aspects of masculinity-femininity: The multidimensional sex role inventory-revised. *Journal of Social Behavior and Personality, 5*(4), 205.

**Justification:** Fewer than 20 citations since 2006

**Masculine role inventory**

**Original scale:** Snell Jr, W. E. (1986). The masculine role inventory: Components and correlates. *Sex Roles, 15*(7-8), 443-455.

**Justification:** Fewer than 20 citations since 2006

**Multifaceted Gender identity questionnaire**

**Original scale:** Willemsen, T. M., & Fischer, A. H. (1999). Assessing multiple facets of gender identity: The Gender Identity Questionnaire. *Psychological Reports, 84*(2), 561-562.

**Justification:** Fewer than 20 citations since 2006

**Masculine Behavior**

**Original scale:** Snell Jr, W. E. (1989). Development and validation of the Masculine Behavior Scale: A measure of behaviors stereotypically attributed to males vs. females.*Sex Roles, 21*(11-12), 749-767.

**Justification:** Fewer than 20 citations since 2006

**Masculine and Feminine Self-Disclosure Scale**

**Original scale:** Snell Jr, W. E., Belk, S. S., & Hawkins II, R. C. (1986). The Masculine and Feminine Self-Disclosure Scale: The politics of masculine and feminine self-presentation. *Sex Roles, 15*(5-6), 249-267.

**Justification:** Fewer than 20 citations since 2006

**Male female relations questionnaire**

**Original scale:** Sherman, P. J., & Spence, J. T. (1997). A COMPARISON OF TWO COHORTS OF COLLEGE STUDENTS IN RESPONSES TO THE MALE-FEMALE RELATIONS QUESTIONNAIRE. *Psychology of Women Quarterly, 21*(2), 265-278.

**Justification:** Fewer than 20 citations since 2006

**Machismo**

**Original scale:** Villemez, W. J., & Touhey, J. C. (1977). A measure of individual differences in sex stereotyping and sex discrimination: The “Macho” scale. *Psychological Reports, 41*(2), 411-415. **Justification:** Fewer than 20 citations since 2006

**Feminine Gender Identity Scale**

**Original scale:** Willemsen, T. M., & Fischer, A. H. (1999). Assessing multiple facets of gender identity: The Gender Identity Questionnaire. *Psychological Reports, 84*(2), 561-562.

**Justification:** Fewer than 20 citations since 2006

**Womanist identity attitude scale**

**Original scale:** Moradi, B., Yoder, J. D., & Berendsen, L. L. (2004). An evaluation of the psychometric properties of the womanist identity attitudes scale. *Sex Roles,50*(3-4), 253-266.

**Justification:** Fewer than 20 citations since 2006

**Assessing the Dynamics of Gender in Couples and Families (Gendergram)**

**Original scale**

White, M. B., & Tyson-Rawson, K. J. (1995). Assessing the dynamics of gender in couples and families: The gendergram. *Family Relations*, 253-260.

**Justification:** Fewer than 20 citations since 2006

**Attitudes toward Multiple Role Planning**

**Original scale**

Weitzman, L. M., & Fitzgerald, L. F. (1996). The development and initial validation of scales to assess attitudes toward multiple role planning. *Journal of Career Assessment, 4*(3), 269-284.

**Justification:** Fewer than 20 citations since 2006

**Traditional vs. Egalitarian Roles in Marriages**

**Original scale**

Altrocchi, J., & Crosby, R. D. (1989). Clarifying and measuring the concept of traditional vs. egalitarian roles in marriages. *Sex roles, 20*(11-12), 639-648.

**Justification:** Fewer than 20 citations since 2006

**Women in Society Questionnaire**

**Original scale:** Walker, L. (1994). Attitudes to minorities: Survey evidence of Western Australians’ attitudes to Aborigines, Asians, and women. *Australian Journal of Psychology, 46*(3), 137-143 **Justification:** Fewer than 20 citations since 2006

**Attitudes to females’ social roles**

**Original scale: Slade, P**., **& Jenner, F. A. (1978). Questionnaire measuring attitudes to females’ social roles. Psychological Reports, 43(2), 351-354**.

**Justification:** Fewer than 20 citations since 2006

**Maferr Inventory of Feminine Values**

**Original scale:** Steinmann, A. G. (1979). Maferr Inventory of Feminine Values: Specimen Set (and) Manual Series for the Interpretation of the Maferr Inventory of Feminine Values (MIFV).

**Justification:** Fewer than 20 citations since 2006

**Tridimensional Sexism Scale**

**Original scale:** Rombough, S., & Ventimiglia, J. C. (1981). Sexism: A tri-dimensional phenomenon. *Sex Roles, 7*(7), 747-755.

**Justification:** Fewer than 20 citations since 2006

**Sex Role Questionnaire**

**Original scale:** Jean, P. J., & Reynolds, C. R. (1980). Development of the Bias in Attitudes Survey: A sex-role questionnaire. *Journal of Psychology, 104*(2), 269.

**Justification:** Fewer than 20 citations since 2006

**Traditional and Liberated Males’ Attitudes**

**Original scale:** Fiebert, M. S. (1983). Measuring traditional and liberated males’ attitudes. Perceptual and motor skills.

**Justification:** Fewer than 20 citations since 2006

**Superficiality and the Dimensionality of Sexism**

**Original scale:** Korth, B. (1978). Superficiality and the Dimensionality of Sexism. Applied Psychological Measurement, 2(1), 51-61.

**Justification:** Fewer than 20 citations since 2006

**The Traditional-Liberated Content Scale**

**Original scale:** Fiebert, M. S., & Vera, W. (1985). Test-retest reliability of a male sex-role attitude survey: The traditional-liberated content scale. *Perceptual and motor skills, 60*(1).

**Justification:** Fewer than 20 citations since 2006

**Multidimensional Aversion to Women who Work Scale**

**Original scale:** Valentine, S. (2001). Development of a brief multidimensional aversion to women who work scale. *Sex Roles, 44*(11-12), 773-787.

**Justification:** Fewer than 20 citations since 2006

**Wellesley Role Orientation Scale**

**Original scal**

Alper, T. G. (1973). The relationship between role orientation and achievement motivation in college women. *Journal of Personality*.

**Justification:** Fewer than 20 citations since 2006

**Sex Role Behavior Scale**

**Original scale:** Orlofsky, J. L., Ramsden, M. W., & Cohen, R. S. (1982). Development of the revised sex-role behavior scale. *Journal of Personality Assessment, 46*, 632–638.

**Justification:** Fewer than 20 citations since 2006

## Step 4: Developing a core list of gender variables

This section describes how we identified the core gender variables for each of the three categories gender-related traits, gender norms and gender relations.

## Identifying core variables for gender norms

Through the literature search, we identified three eligible measures pertaining to the gender norms category. The first two measures (see below) concern child-rearing and co-parenting behavior. These aspects will be captured with a slightly broader variable focusing on various forms of caregiving. We hereby extend the restricted focus on “nuclear” family formations proposed by existing measures. The third measure focuses on dual-career wives’ orientation towards domestic labour. We also intend to capture this aspect with a broader variable focusing on time-use for both women and men. An additional variable focusing work-related physical and psychological strain will also be included under this category.

Caregiving

- Parental Child-rearing Behavior Scale
- Co-parenting and family rating system

Time-use

- The orientation towards domestic labor

## Identifying core variables for gender-related traits

In developing our final list of gender-related traits, we began by harvesting all relevant characteristics and traits covered by the 31 eligible gender-related traits scales identified in Step 3 (Table S28). Next, we aggregated similar and closely related gender traits and characteristics into 17 clusters, each capturing distinct aspects of an individual’s gender identity, using inductive coding categories. Numerous cluster-solutions (i.e. solutions varying on the total number of clusters since characteristics and traits can be merged or split into smaller or larger clusters) were scrutinized as part of this step. Consensus was ultimately established on the 17-cluster solution as the most intuitively meaningful and feasible way of structuring the many traits and characteristics into key gender-related traits variables for health research (Table S29). We then ranked the clusters based on occurrences (i.e. number of characteristics and traits pertaining to each cluster) to obtain a closer understanding of their prevalence across existing scales. All in all, this strategy helped us make sense of the literature and carve out a condensed list of core gender-related traits variables. Specifically, we selected the four most prevalent clusters emerging from our coding. Risk-taking is not among the “top-scorers,” but has been included as a fifth variable given its well-documented links to health outcomes.

1. Emotionality and expressiveness (37 occurrences) (Captured by *Expressive*)
2. Empathy, caring and nurturing (37 occurrences) (Captured by *Empathetic/caring/nurturing*)
3. Self-reliance and independence (18 occurrences) (Captured by *Self-reliant/independent*)
4. Striving for status, competitiveness (17 occurrences) (Captured by *Competitive/striving*)
5. Risk-taking (5 occurrences) (Captured by *Sensation-seeking*/*risk-taking*)

## Identifying core variables for gender relations

The literature search resulted in 40 scales pertaining to the gender relations category. Specifically, these scales capture two core variables for gender relations: equality in family relationships and discrimination. An additional variable focusing more broadly on social support will also be included under this category.

Gender discrimination

- Gender Minority Stress and Resilience
- Multidimensional Measure of Sexual Minority Identity
- The Ambivalent Sexism Inventory
- Race and Gender-Specific Stress Measure for African-American Women
- Schedule of Sexist Events
- African American Men’s Gendered Racism Stress Inventory

Sexism and attitudes

- Patriarchal Beliefs Scale
- Modern and Old-Fashioned Sexism
- Sex-Role Ideology
- Gender Ideology as an Identity
- Gender Attitude Inventory
- Quick Discrimination Index
- Sex-Role Stereotypes
- Attitudes toward Traditional and Egalitarian Sex-Roles
- Women as Managers Scale
- Neosexism Scale
- Swedish Classical and Modern Sexism Scale
- Sexist Attitudes toward Women
- Sex-Role Egalitarianism Scale
- Attitudes toward Women Scale
- Gender Role Egalitarian Attitudes Scale
- Adherence to Extreme Gender Role Beliefs
- Sex-Role Orientation
- Gender Role Beliefs Scale
- Male Attitude Norms Inventory II
- Egalitarian Sex Role Attitudes

Equality in family relationships

- Dyadic Adjustment Scale across Gender
- Quality in Relationships Inventory within Couples
- Conflict Disengagement Inventory (CDI)
- German Relationship Assessment Scale (*Same as Relationship Assessment Scale)
- Partner Behavior Inventory
- Relationship Assessment Scale
- Relationship Belief Inventory
- Managing Affect and Differences Scale
- Concord-Index (measure of family relationships)
- Mutual Psychological Development Questionnaire (measuring perceived mutuality in close relationships)
- Relationship Authenticity (Authenticity in Relationships Scale)
- The Romantic Beliefs Scale
- Routine and Strategic Relational Maintenance Scale
- Gender-Based Attitudes Toward Marriage and Child Rearing

## Step 5: Identifying measures for each gender variable

This section documents the search strategy used to identify additional construct specific measures of relevance to our gender variables. We for searched for meta-analyses in PsycINFO and PubMed and restricted the time-span to the period 2006 through 2015.

## Search limitations

We used the following database-specific search limitations:

### PsycINFO

Age-group: Adulthood (18 years or older); Record type: Peer-reviewed journal, Abstract collection, Peer-reviewed articles; Methodology: Meta-Analysis, Metasynthesis, systematic review; Language: English; Population: Human; PsycINFO Classifiers: “Tests and testing”; “Personality Scales &Inventories”; “Clinical Psychological Testing”; “Health Psychology Testing”; “Educational measurement”; “Occupational and employment testing”; “Consumer opinion and attitudes”.

### PUBMED

Ages: all ages; Species: Humans; Language: English; Article types: all.

## Search strings

The search terms used in this part of the literature search are listed in Table S30.

Type or paste text here. This should be additional explanatory text such as an extended technical description of results, full details of mathematical models, etc.

### Heading

#### Subhead

Type or paste text here. You may break this section up into subheads as needed (e.g., one section on “Materials” and one on “Methods”).

**Fig S1.**
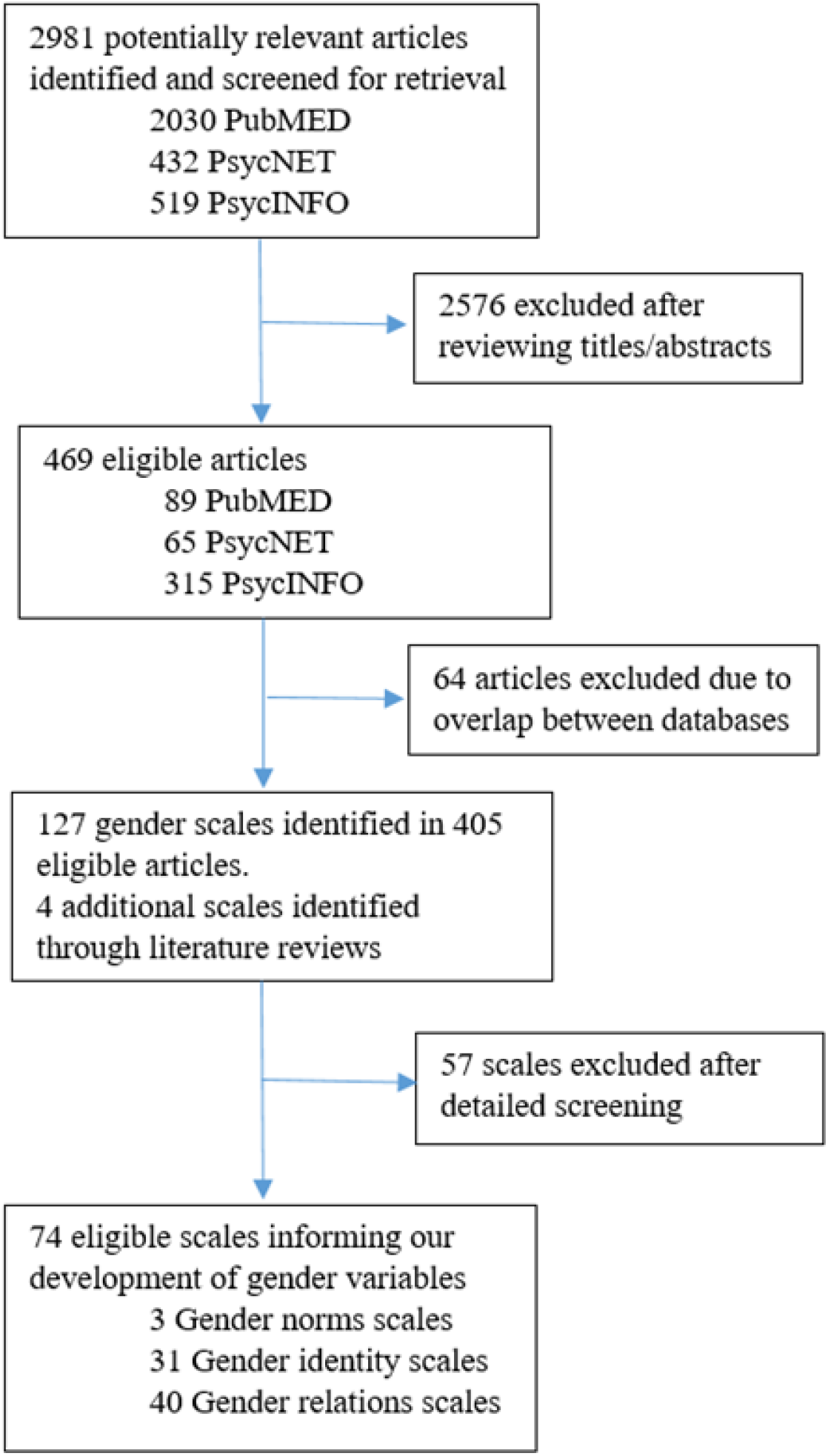
Flowchart of article inclusion and exclusion in the literature search.

**Fig S2.**
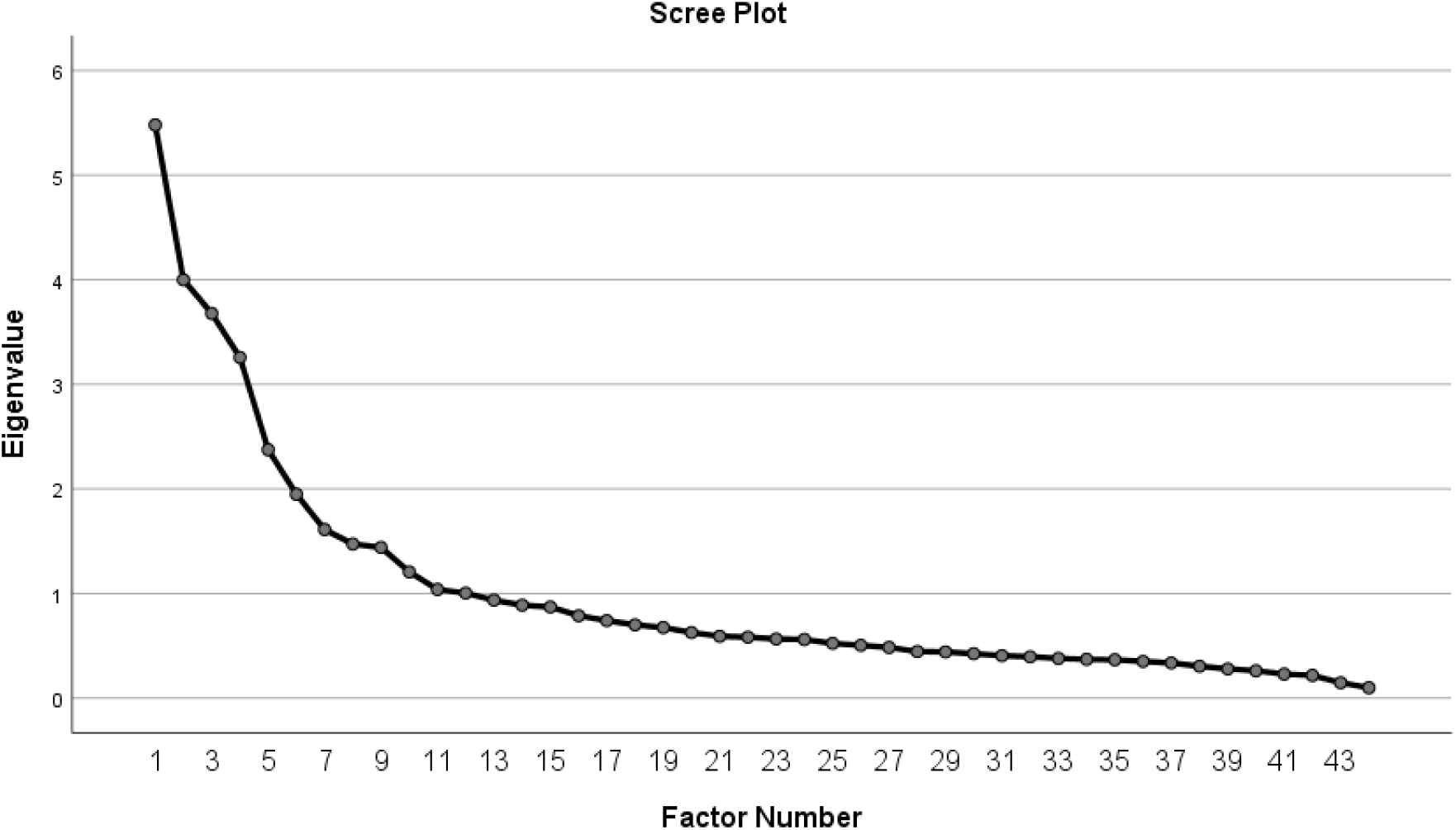
Screeplot of the factor analysis reported in Table S34.

**Table S1.**
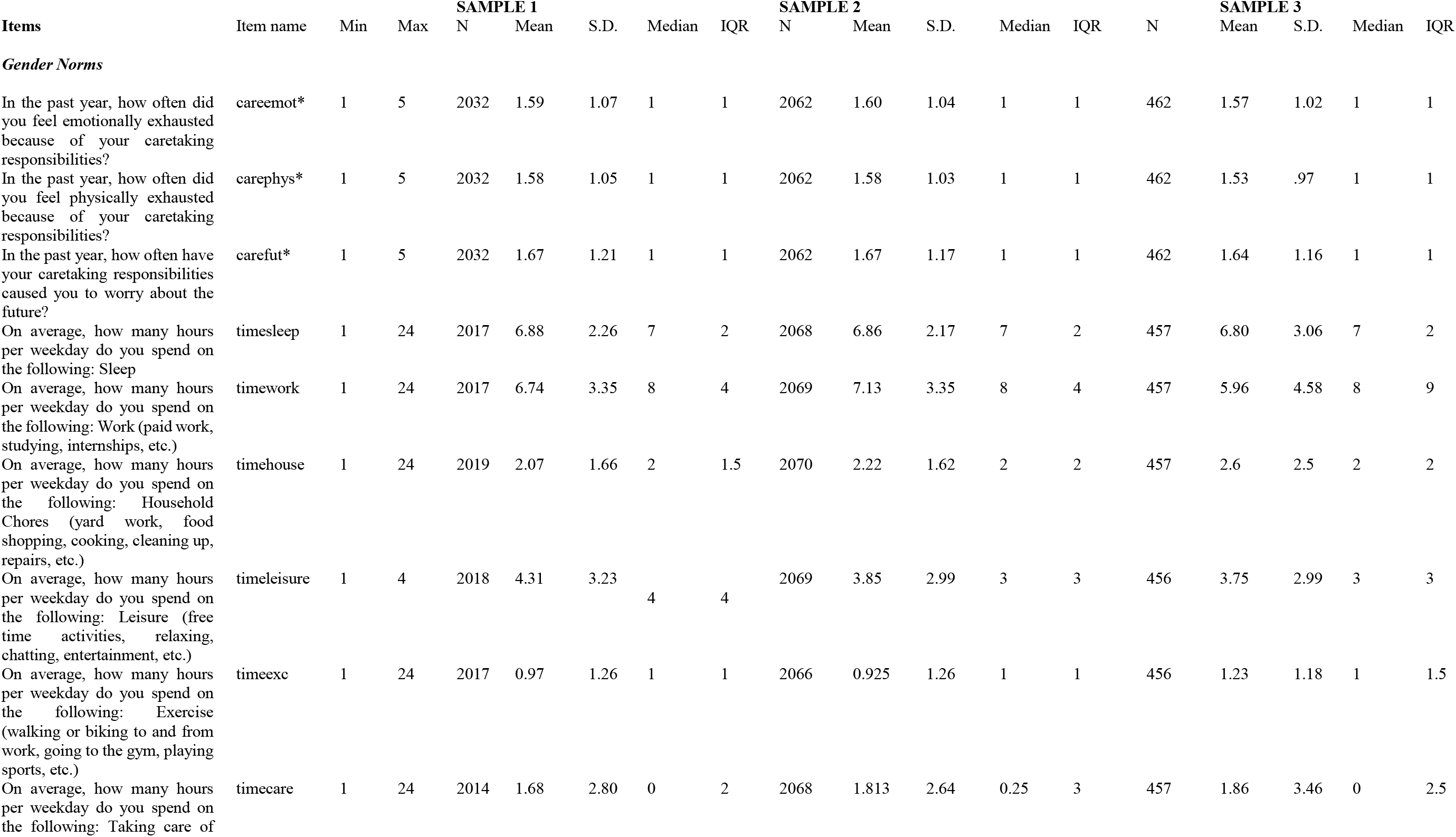
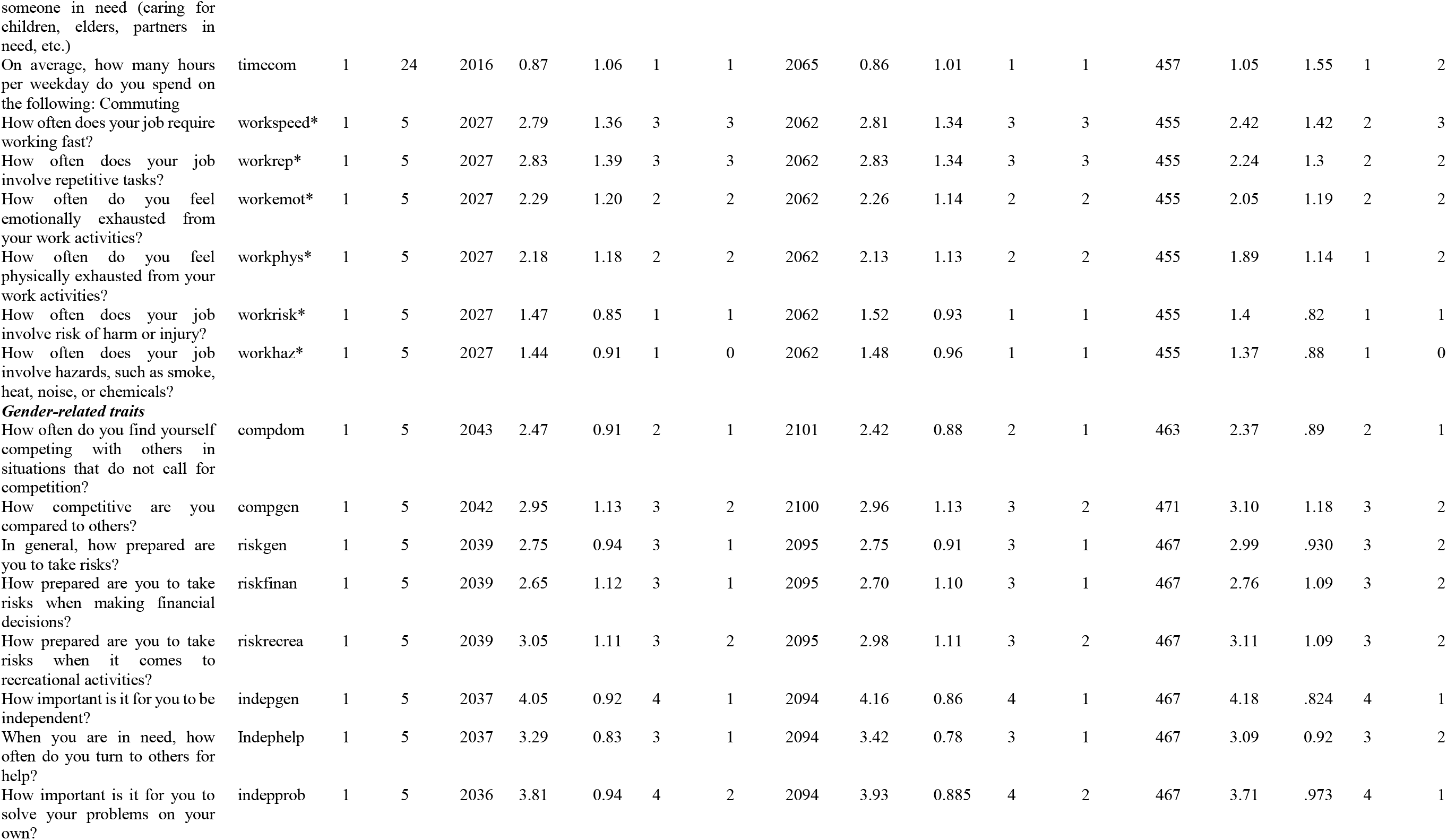
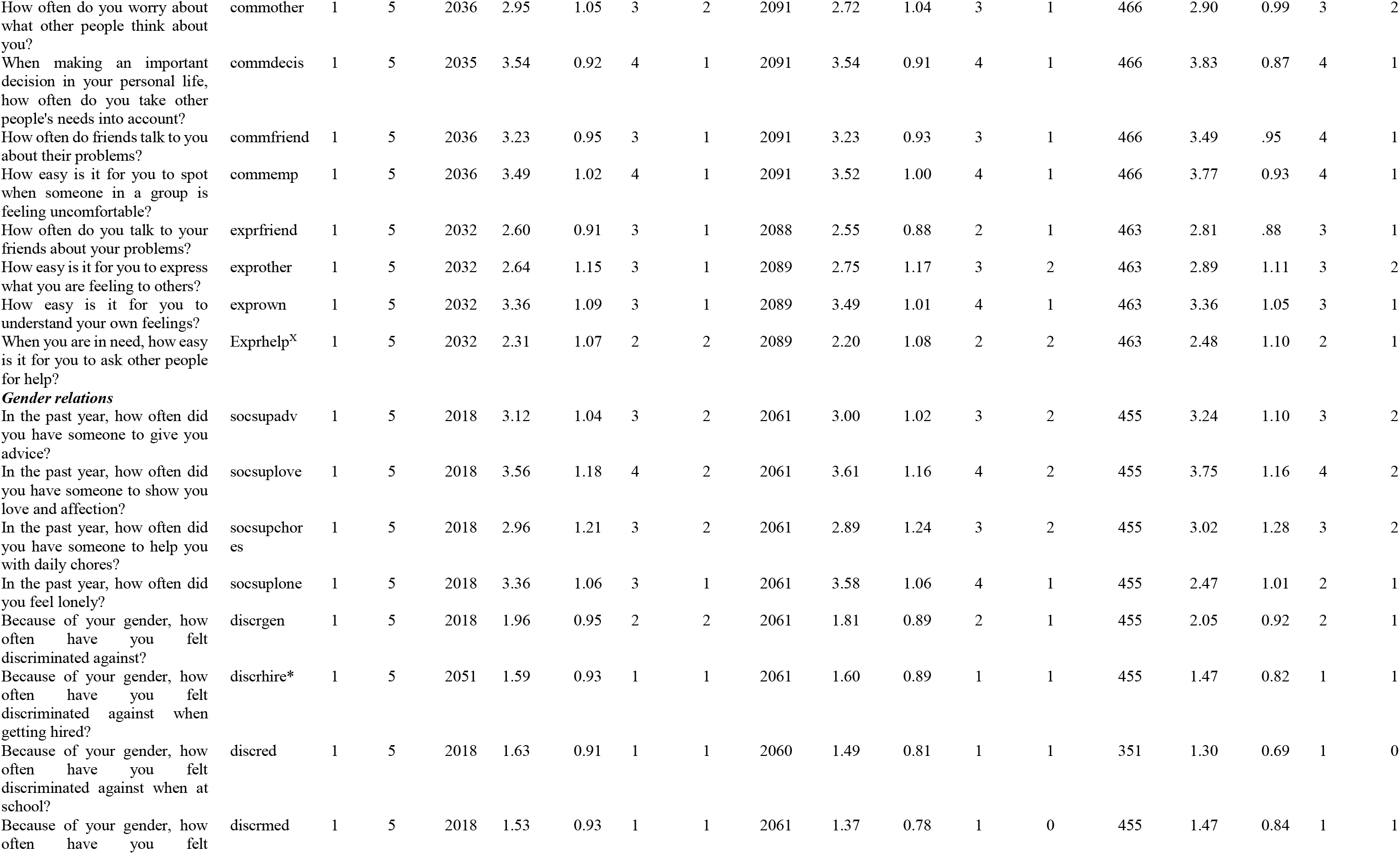
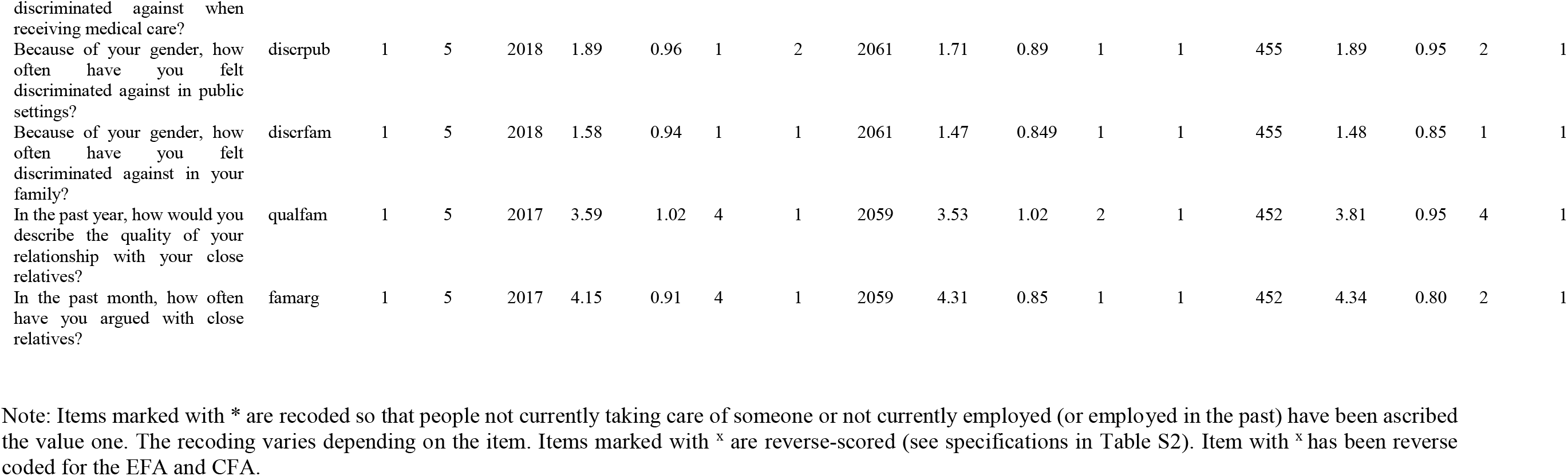
Item phrasing and descriptive statistics for the 44 potentially relevant gender-related items.

**Table S2.**
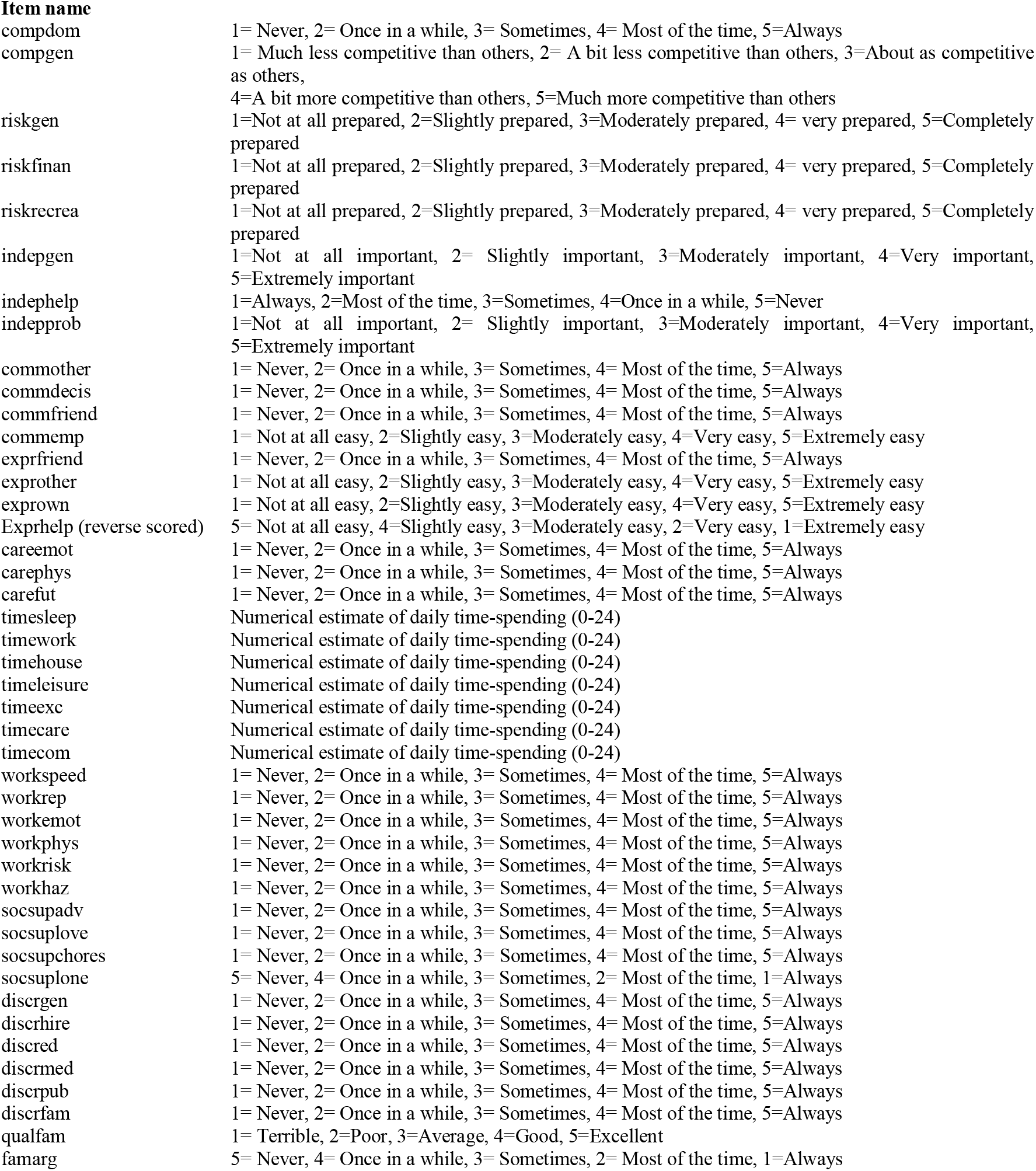
Response options for all 44 items included in the exploratory factor analyses.

**Table S3.** Correlations between the factors in samples 1, 2 and 3.

| **Sample 1** | Independent | Emotional Intelligence | Social Support | Risk-taking | Caregiver strain | Work strain | Discrimination |
| --- | --- | --- | --- | --- | --- | --- | --- |
| Independent | 1.000 |  |  |  |  |  |  |
| Emotional Intelligence | -0.101 | 1.000 |  |  |  |  |  |
| Social Support | -0.105 | 0.364 | 1.000 |  |  |  |  |
| Risk-taking | 0.256 | 0.236 | 0.145 | 1.000 |  |  |  |
| Caregiver strain | 0.071 | 0.133 | 0.101 | 0.043 | 1.000 |  |  |
| Work strain | 0.151 | 0.129 | 0.032 | 0.219 | 0.105 | 1.000 |  |
| Discrimination | 0.031 | 0.207 | 0.021 | 0.049 | 0.13 | 0.094 | 1.000 |
| **Sample 2** | Independent | Emotional Intelligence | Social Support | Risk-taking | Caregiver strain | Work strain | Discrimination |
| Independent | 1.000 |  |  |  |  |  |  |
| Emotional Intelligence | -0.196 | 1.000 |  |  |  |  |  |
| Social Support | -0.077 | 0.314 | 1.000 |  |  |  |  |
| Risk-taking | 0.174 | 0.18 | 0.149 | 1.000 |  |  |  |
| Caregiver strain | 0.058 | 0.055 | 0.036 | 0.032 | 1.000 |  |  |
| Work strain | 0.105 | 0.076 | 0.036 | 0.133 | 0.101 | 1.000 |  |
| Discrimination | -0.087 | 0.182 | -0.053 | 0.001 | 0.132 | 0.108 | 1.000 |
| **Sample 3** | Independent | Emotional Intelligence | Social Support | Risk-taking | Caregiver strain | Work strain | Discrimination |
| Independent | 1.000 |  |  |  |  |  |  |
| Emotional Intelligence | -.261 | 1.000 |  |  |  |  |  |
| Social Support | -.224 | .340 | 1.000 |  |  |  |  |
| Risk-taking | .104 | .194 | .009 | 1.000 |  |  |  |
| Caregiver strain | -.076 | .055 | .041 | -.048 | 1.000 |  |  |
| Work strain | .089 | .065 | -.010 | -.027 | .108 | 1.000 |  |
| Discrimination | .012 | .118 | .033 | -.018 | .220 | .068 | 1.000 |

Note: Sample 1= 2009, Sample 2=2054, Sample 3= 449. The discrimination variable in sample 3 was computed without the discred variable.

**Table S4.** Negative binomial regression predicting number of days with poor physical health (during past 30 days) (with gender identity as covariate).

|  | *SAMPLE 1* |  |  | *SAMPLE 2* |  |  |
| --- | --- | --- | --- | --- | --- | --- |
|  | Incidence ratio | rate | 95% CI | Incidence ratio | rate | 95% CI |
| Caregiver strain |  | 1.20 | 1.12 1.29 |  | 1.11 | 1.04 1.18 |
| Work strain |  | 0.96 | 0.88 1.05 |  | 1.01 | 0.93 1.10 |
| Independence |  | 0.93 | 0.85 1.01 |  | 0.85 | 0.75 0.95 |
| Risk-taking |  | 0.95 | 0.87 1.04 |  | 1.00 | 0.92 1.08 |
| Emotional intelligence |  | 0.93 | 0.85 1.03 |  | 0.92 | 0.85 1.01 |
| Social support |  | 0.95 | 0.89 1.03 |  | 0.94 | 0.89 1.00 |
| Discrimination |  | 1.45 | 1.33 1.58 |  | 1.43 | 1.31 1.57 |
| Year of birth |  | 0.99 | 0.98 1.00 |  | 0.99 | 0.98 0.99 |
| Personal Income (\$10,000-\$19,999) | | 1.02 | 0.80 1.29 | | 1.07 | 0.86 1.33 |
| Personal Income (\$20,000-\$29,999) | | 0.94 | 0.73 1.21 | | 1.01 | 0.81 1.25 |
| Personal Income (\$30,000-\$39,999) | | 0.84 | 0.64 1.10 | | 0.87 | 0.68 1.11 |
| Personal Income (\$40,000-\$49,999) | | 1.01 | 0.76 1.33 | | 0.71 | 0.56 0.91 |
| Personal Income (\$50,000-\$59,999) | | 0.93 | 0.68 1.27 | | 0.68 | 0.50 0.92 |
| Personal Income (\$60,000-\$69,999) | | 0.83 | 0.60 1.16 | | 0.70 | 0.52 0.95 |
| Personal Income (\$70,000-\$79,999) | | 0.82 | 0.54 1.23 | | 0.61 | 0.44 0.85 |
| Personal Income (\$80,000-\$89,999) | | 0.65 | 0.41 1.02 | | 0.45 | 0.32 0.62 |
| Personal Income (\$90,000-\$99,999) | | 1.03 | 0.56 1.92 | | 0.80 | 0.54 1.19 |
| Personal Income (\$100,000-\$149,999) | | 0.78 | 0.52 1.17 | | 0.67 | 0.47 0.96 |
| Personal Income (More than \$150,000) | | 1.36 | 0.88 2.10 | | 0.55 | 0.29 1.03 |
| Educational Level (High school graduate) |  | 1.04 | 0.64 1.70 |  | 0.88 | 0.50 1.55 |
| Educational Level (Some college credit, no degree) |  | 0.88 | 0.55 1.40 |  | 1.12 | 0.65 1.92 |
| Educational Level (Trade/Technical/Vocational education) |  | 0.86 | 0.48 1.54 |  | 0.83 | 0.46 1.51 |
| Educational Level (Associate degree) |  | 0.79 | 0.48 1.30 |  | 1.00 | 0.58 1.74 |
| Educational Level (Bachelor's degree) |  | 0.75 | 0.47 1.20 |  | 0.85 | 0.50 1.46 |
| Educational Level (Master's degree) |  | 0.89 | 0.54 1.48 |  | 0.84 | 0.48 1.48 |
| Educational Level (professional degree) |  | 0.71 | 0.24 2.05 |  | 0.92 | 0.43 1.95 |
| Educational Level (Doctorate degree) |  | 0.68 | 0.37 1.25 |  | 0.69 | 0.35 1.33 |
| Ethnicity (White) |  | 1.02 | 0.77 1.36 |  | 0.96 | 0.74 1.24 |
| Ethnicity (Hispanic, Latinx, Spanish) |  | 0.98 | 0.71 1.37 |  | 0.96 | 0.70 1.31 |
| Ethnicity (Black, African American) |  | 0.94 | 0.67 1.30 |  | 0.86 | 0.66 1.12 |
| Ethnicity (Asian) |  | 0.83 | 0.60 1.15 |  | 0.87 | 0.61 1.24 |
| Ethnicity (Native) |  | 0.91 | 0.67 1.24 |  | 1.64 | 1.12 2.41 |
| Woman |  | 0.95 | 0.82 1.11 |  | 0.93 | 0.80 1.07 |
| Non-binary/Gender fluid + Gender=other |  | 0.96 | 0.58 1.60 |  | 0.44 | 0.17 1.15 |
| **Goodness of Fit** |  |  |  |  |  |  |
| Deviance/df |  | 1.114 |  |  | 1.120 |  |
| Pearson Chi-Square/df |  | 1.240 |  |  | 1.159 |  |
| Loglikelihood |  | -4848.379 |  |  | -5161.248 |  |
| AIC |  | 9768.758 |  |  | 10394.496 |  |
| **Omnibus tests** |  |  |  |  |  |  |
| Chi-square |  | 183.600 |  |  | 204.826 |  |
| df |  | 34 |  |  | 34 |  |
| N |  | 1881 |  |  | 2008 |  |

Note: Participants reporting Native Hawaiian/Pacific Islander, Middle east/North Africa or Other as ethnicity are the reference group for the ethnicity variables. Personal Income < \$10,000 is the reference group for the "Personal Income" variables. Educational level < high school diploma is the reference group for the "Educational Level" variables. The models are computed with robust covariance matrix estimation.

**Table S5.** Negative Binomial regression predicting number of days with poor mental health (during past 30 days) (with gender identity as covariate).

|  | *SAMPLE 1* |  |  | *SAMPLE 2* |  |  |
| --- | --- | --- | --- | --- | --- | --- |
|  | Incidence ratio | rate | 95% CI | Incidence ratio | rate | 95% CI |
| Caregiver strain |  | 1.21 | 1.15 1.27 |  | 1.20 | 1.14 1.27 |
| Work strain |  | 1.03 | 0.96 1.11 |  | 1.12 | 1.04 1.21 |
| Independence |  | 0.98 | 0.91 1.04 |  | 0.83 | 0.74 0.93 |
| Risk-taking |  | 0.86 | 0.80 0.93 |  | 0.99 | 0.91 1.08 |
| Emotional intelligence |  | 0.97 | 0.90 1.05 |  | 0.86 | 0.79 0.94 |
| Social support |  | 0.86 | 0.81 0.90 |  | 0.81 | 0.76 0.86 |
| Discrimination |  | 1.42 | 1.33 1.51 |  | 1.36 | 1.24 1.49 |
| Year of birth |  | 1.02 | 1.01 1.02 |  | 1.02 | 1.01 1.02 |
| Personal Income (\$10,000-\$19,999) | | 0.98 | 0.82 1.16 | | 1.03 | 0.84 1.25 |
| Personal Income (\$20,000-\$29,999) | | 0.75 | 0.62 0.91 | | 0.89 | 0.71 1.10 |
| Personal Income (\$30,000-\$39,999) | | 0.78 | 0.63 0.96 | | 0.85 | 0.68 1.05 |
| Personal Income (\$40,000-\$49,999) | | 0.76 | 0.62 0.95 | | 0.63 | 0.50 0.80 |
| Personal Income (\$50,000-\$59,999) | | 0.81 | 0.65 1.01 | | 0.51 | 0.38 0.68 |
| Personal Income (\$60,000-\$69,999) | | 0.72 | 0.56 0.92 | | 0.54 | 0.40 0.73 |
| Personal Income (\$70,000-\$79,999) | | 0.69 | 0.50 0.96 | | 0.55 | 0.39 0.76 |
| Personal Income (\$80,000-\$89,999) | | 0.75 | 0.52 1.08 | | 0.77 | 0.47 1.26 |
| Personal Income (\$90,000-\$99,999) | | 0.98 | 0.66 1.45 | | 0.61 | 0.38 0.97 |
| Personal Income (\$100,000-\$149,999) | | 0.59 | 0.42 0.82 | | 0.62 | 0.44 0.88 |
| Personal Income (More than \$150,000) | | 1.00 | 0.67 1.48 | | 0.43 | 0.25 0.74 |
| Educational Level (High school graduate) |  | 1.06 | 0.72 1.58 |  | 1.50 | 0.86 2.61 |
| Educational Level (Some college credit, no degree) |  | 1.04 | 0.72 1.52 |  | 1.68 | 0.98 2.85 |
| Educational Level (Trade/Technical/Vocational education) |  | 1.15 | 0.73 1.81 |  | 1.90 | 1.03 3.53 |
| Educational Level (Associate degree) |  | 1.01 | 0.67 1.52 |  | 1.61 | 0.93 2.77 |
| Educational Level (Bachelor's degree) |  | 0.88 | 0.61 1.28 |  | 1.53 | 0.90 2.60 |
| Educational Level (Master's degree) |  | 1.07 | 0.72 1.59 |  | 1.61 | 0.92 2.81 |
| Educational Level (professional degree) |  | 0.71 | 0.36 1.41 |  | 1.84 | 0.90 3.77 |
| Educational Level (Doctorate degree) |  | 0.91 | 0.57 1.46 |  | 1.74 | 0.87 3.48 |
| Ethnicity (White) |  | 1.53 | 1.24 1.89 |  | 1.32 | 1.00 1.73 |
| Ethnicity (Hispanic, Latinx, Spanish) |  | 1.06 | 0.85 1.32 |  | 1.10 | 0.83 1.47 |
| Ethnicity (Black, African American) |  | 1.27 | 1.00 1.62 |  | 0.94 | 0.71 1.24 |
| Ethnicity (Asian) |  | 1.13 | 0.88 1.45 |  | 1.08 | 0.75 1.54 |
| Ethnicity (Native) |  | 1.06 | 0.80 1.41 |  | 1.88 | 1.27 2.79 |
| Woman |  | 0.95 | 0.84 1.07 |  | 1.20 | 1.04 1.39 |
| Non-binary/Gender fluid + Gender=other |  | 1.24 | 0.96 1.61 |  | 0.92 | 0.43 1.98 |
| **Goodness of Fit** |  |  |  |  |  |  |
| Deviance/df |  | 1.173 |  |  | 1.131 |  |
| Pearson Chi-Square/df |  | 1.016 |  |  | 1.106 |  |
| Loglikelihood |  | -5669.909 |  |  | -5571.861 |  |
| AIC |  | 11411.818 |  |  | 11215.721 |  |
| **Omnibus tests** |  |  |  |  |  |  |
| Chi-square |  | 285.752 |  |  | 316.349 |  |
| df |  | 34 |  |  | 34 |  |
| N |  | 1883 |  |  | 2005 |  |

Note: Participants reporting Native Hawaiian/Pacific Islander, Middle east/North Africa or Other as ethnicity are the reference group for the ethnicity variables. Personal Income < \$10,000 is the reference group for the "Personal Income" variables. Educational level < high school diploma is the reference group for the "Educational Level" variables. The models are computed with robust covariance matrix estimation.

**Table S6.** Negative binomial regression predicting number of days where poor mental or physical health prevented the respondent from doing usual activities (during past 30 days) (with gender identity as covariate).

|  | *SAMPLE 1* |  |  | *SAMPLE 2* |  |  |
| --- | --- | --- | --- | --- | --- | --- |
|  | Incidence ratio | rate | 95% CI | Incidence ratio | rate | 95% CI |
| Caregiver strain |  | 1.13 | 1.05 1.22 |  | 1.10 | 1.02 1.19 |
| Work strain |  | 0.88 | 0.80 0.97 |  | 0.98 | 0.90 1.08 |
| Independence |  | 1.02 | 0.94 1.12 |  | 0.88 | 0.77 1.01 |
| Risk-taking |  | 0.93 | 0.85 1.02 |  | 1.02 | 0.93 1.11 |
| Emotional intelligence |  | 0.99 | 0.89 1.10 |  | 0.92 | 0.83 1.01 |
| Social support |  | 0.92 | 0.86 0.99 |  | 0.87 | 0.81 0.94 |
| Discrimination |  | 1.74 | 1.59 1.90 |  | 1.57 | 1.41 1.74 |
| Year of birth |  | 1.01 | 1.00 1.01 |  | 1.00 | 1.00 1.01 |
| Personal Income (\$10,000-\$19,999) | | 1.01 | 0.80 1.27 | | 0.90 | 0.70 1.16 |
| Personal Income (\$20,000-\$29,999) | | 0.90 | 0.70 1.16 | | 0.92 | 0.72 1.17 |
| Personal Income (\$30,000-\$39,999) | | 0.76 | 0.56 1.02 | | 0.77 | 0.59 1.01 |
| Personal Income (\$40,000-\$49,999) | | 0.92 | 0.68 1.23 | | 0.65 | 0.48 0.87 |
| Personal Income (\$50,000-\$59,999) | | 0.76 | 0.56 1.04 | | 0.52 | 0.37 0.73 |
| Personal Income (\$60,000-\$69,999) | | 0.63 | 0.44 0.90 | | 0.68 | 0.47 0.97 |
| Personal Income (\$70,000-\$79,999) | | 0.67 | 0.42 1.05 | | 0.62 | 0.43 0.89 |
| Personal Income (\$80,000-\$89,999) | | 0.99 | 0.57 1.70 | | 0.64 | 0.40 1.03 |
| Personal Income (\$90,000-\$99,999) | | 1.00 | 0.51 1.95 | | 0.70 | 0.44 1.11 |
| Personal Income (\$100,000-\$149,999) | | 0.91 | 0.58 1.42 | | 0.70 | 0.46 1.06 |
| Personal Income (More than \$150,000) | | 1.57 | 0.95 2.60 | | 0.75 | 0.38 1.47 |
| Educational Level (High school graduate) |  | 1.07 | 0.64 1.78 |  | 0.99 | 0.51 1.92 |
| Educational Level (Some college credit, no degree) |  | 0.85 | 0.52 1.39 |  | 1.14 | 0.61 2.13 |
| Educational Level (Trade/Technical/Vocational education) |  | 1.32 | 0.74 2.36 |  | 0.81 | 0.40 1.65 |
| Educational Level (Associate degree) |  | 0.82 | 0.48 1.40 |  | 0.96 | 0.51 1.82 |
| Educational Level (Bachelor's degree) |  | 0.87 | 0.53 1.41 |  | 0.77 | 0.42 1.44 |
| Educational Level (Master's degree) |  | 0.72 | 0.43 1.22 |  | 0.70 | 0.37 1.33 |
| Educational Level (professional degree) |  | 0.24 | 0.10 0.56 |  | 0.51 | 0.22 1.18 |
| Educational Level (Doctorate degree) |  | 0.64 | 0.36 1.15 |  | 0.61 | 0.28 1.31 |
| Ethnicity (White) |  | 1.11 | 0.84 1.48 |  | 0.89 | 0.65 1.21 |
| Ethnicity (Hispanic, Latinx, Spanish) |  | 0.88 | 0.66 1.18 |  | 1.00 | 0.70 1.43 |
| Ethnicity (Black, African American) |  | 0.99 | 0.72 1.37 |  | 0.76 | 0.54 1.06 |
| Ethnicity (Asian) |  | 0.77 | 0.56 1.05 |  | 0.85 | 0.56 1.31 |
| Ethnicity (Native) |  | 0.93 | 0.62 1.39 |  | 2.03 | 1.26 3.28 |
| Woman |  | 0.88 | 0.74 1.03 |  | 1.09 | 0.92 1.29 |
| Non-binary/Gender fluid + Gender=other |  | 1.30 | 0.82 2.04 |  | 0.62 | 0.16 2.45 |
| **Goodness of Fit** |  |  |  |  |  |  |
| Deviance/df |  | 1.081 |  |  | 1.057 |  |
| Pearson Chi-Square/df |  | 1.165 |  |  | 1.148 |  |
| Loglikelihood |  | -4584.605 |  |  | -4640.077 |  |
| AIC |  | 9241.210 |  |  | 9352.154 |  |
| **Omnibus tests** |  |  |  |  |  |  |
| Chi-square |  | 250.517 |  |  | 222.470 |  |
| df |  | 34 |  |  | 34 |  |
| N |  | 1879 |  |  | 2007 |  |

Note: Participants reporting Native Hawaiian/Pacific Islander, Middle east/North Africa or Other as ethnicity are the reference group for the ethnicity variables. Personal Income < \$10,000 is the reference group for the "Personal Income" variables. Educational level < high school diploma is the reference group for the "Educational Level" variables. The models are computed with robust covariance matrix estimation.

**Table S7.** Logistic regression predicting general health status (excellent, very good, good= 0, fair, poor= 1) (with gender identity as covariate)

|  | *SAMPLE 1* |  |  | *SAMPLE 2* |  |  |
| --- | --- | --- | --- | --- | --- | --- |
|  | Incidence ratio | rate | 95% CI | Incidence ratio | rate | 95% CI |
| Caregiver strain |  | 1.28 | 1.12 1.47 |  | 1.17 | 1.03 1.34 |
| Work strain |  | 0.87 | 0.73 1.05 |  | 0.99 | 0.83 1.17 |
| Independence |  | 0.91 | 0.77 1.06 |  | 0.71 | 0.56 0.90 |
| Risk-taking |  | 0.56 | 0.47 0.67 |  | 0.69 | 0.58 0.81 |
| Emotional intelligence |  | 0.92 | 0.76 1.11 |  | 0.89 | 0.75 1.06 |
| Social support |  | 0.83 | 0.73 0.95 |  | 0.78 | 0.69 0.88 |
| Discrimination |  | 1.43 | 1.18 1.73 |  | 1.57 | 1.29 1.89 |
| Year of birth |  | 0.98 | 0.97 0.99 |  | 0.98 | 0.97 0.99 |
| Personal Income (\$10,000-\$19,999) | | 0.79 | 0.52 1.20 | | 1.19 | 0.78 1.80 |
| Personal Income (\$20,000-\$29,999) | | 0.88 | 0.57 1.36 | | 0.83 | 0.54 1.28 |
| Personal Income (\$30,000-\$39,999) | | 0.61 | 0.37 1.02 | | 0.82 | 0.51 1.32 |
| Personal Income (\$40,000-\$49,999) | | 0.76 | 0.46 1.28 | | 0.62 | 0.37 1.04 |
| Personal Income (\$50,000-\$59,999) | | 0.62 | 0.37 1.05 | | 0.52 | 0.29 0.93 |
| Personal Income (\$60,000-\$69,999) | | 0.86 | 0.48 1.55 | | 0.78 | 0.41 1.49 |
| Personal Income (\$70,000-\$79,999) | | 0.61 | 0.28 1.32 | | 0.63 | 0.33 1.22 |
| Personal Income (\$80,000-\$89,999) | | 0.19 | 0.04 0.81 | | 0.52 | 0.20 1.33 |
| Personal Income (\$90,000-\$99,999) | | 0.30 | 0.06 1.51 | | 1.01 | 0.46 2.23 |
| Personal Income (\$100,000-\$149,999) | | 0.11 | 0.02 0.48 | | 0.60 | 0.30 1.20 |
| Personal Income (More than \$150,000) | | 0.09 | 0.01 0.71 | | 0.73 | 0.23 2.33 |
| Educational Level (High school graduate) |  | 0.45 | 0.19 1.09 |  | 0.84 | 0.28 2.54 |
| Educational Level (Some college credit, no degree) |  | 0.58 | 0.25 1.32 |  | 0.93 | 0.32 2.68 |
| Educational Level (Trade/Technical/Vocational education) |  | 0.56 | 0.19 1.60 |  | 0.79 | 0.25 2.51 |
| Educational Level (Associate degree) |  | 0.42 | 0.17 1.05 |  | 1.01 | 0.34 2.96 |
| Educational Level (Bachelor's degree) |  | 0.41 | 0.18 0.95 |  | 0.50 | 0.17 1.45 |
| Educational Level (Master's degree) |  | 0.47 | 0.19 1.15 |  | 0.39 | 0.13 1.19 |
| Educational Level (professional degree) |  | 0.35 | 0.06 2.04 |  | 0.24 | 0.05 1.24 |
| Educational Level (Doctorate degree) |  | 0.17 | 0.04 0.74 |  | 0.63 | 0.17 2.35 |
| Ethnicity (White) |  | 1.95 | 1.08 3.53 |  | 0.99 | 0.52 1.87 |
| Ethnicity (Hispanic, Latinx, Spanish) |  | 0.83 | 0.42 1.63 |  | 1.06 | 0.55 2.04 |
| Ethnicity (Black, African American) |  | 2.31 | 1.20 4.42 |  | 0.90 | 0.46 1.77 |
| Ethnicity (Asian) |  | 1.69 | 0.88 3.27 |  | 0.76 | 0.32 1.82 |
| Ethnicity (Native) |  | 0.46 | 0.18 1.16 |  | 0.86 | 0.38 1.97 |
| Woman |  | 0.57 | 0.42 0.78 |  | 0.65 | 0.48 0.87 |
| Non-binary/Gender fluid + Gender=other |  | 1.08 | 0.40 2.93 |  | 0.33 | 0.03 3.69 |
| **Goodness of Fit** |  |  |  |  |  |  |
| Deviance/df |  | 0.826 |  |  | 0.877 |  |
| Pearson Chi-Square/df |  | 1.015 |  |  | 1.015 |  |
| Loglikelihood |  | -766.888 |  |  | -867.561 |  |
| AIC |  | 1603.776 |  |  | 1805.123 |  |
| **Omnibus tests** |  |  |  |  |  |  |
| Chi-square |  | 210.935 |  |  | 191.691 |  |
| df |  | 34 |  |  | 34 |  |
| N |  | 1891 |  |  | 2013 |  |

Note: Participants reporting Native Hawaiian/Pacific Islander, Middle east/North Africa or Other as ethnicity are the reference group for the ethnicity variables. Personal Income < \$10,000 is the reference group for the "Personal Income" variables. Educational level < high school diploma is the reference group for the "Educational Level" variables. The models are computed with robust covariance matrix estimation.

**Table S8.** Logistic regression predicting vaping (not vaping=0, vaping=1) (with gender identity as covariate).

|  | *SAMPLE 1* |  |  | *SAMPLE 2* |  |  |
| --- | --- | --- | --- | --- | --- | --- |
|  | Incidence ratio | rate | 95% CI | Incidence ratio | rate | 95% CI |
| Caregiver strain |  | 1.06 | 0.90 1.25 |  | 1.10 | 0.93 1.29 |
| Work strain |  | 1.21 | 0.96 1.53 |  | 1.29 | 1.05 1.59 |
| Independence |  | 1.18 | 0.96 1.45 |  | 1.05 | 0.77 1.44 |
| Risk-taking |  | 1.18 | 0.97 1.45 |  | 1.00 | 0.80 1.24 |
| Emotional intelligence |  | 1.39 | 1.12 1.72 |  | 1.05 | 0.84 1.31 |
| Social support |  | 0.98 | 0.84 1.14 |  | 0.89 | 0.76 1.04 |
| Discrimination |  | 1.40 | 1.16 1.70 |  | 1.47 | 1.18 1.83 |
| Year of birth |  | 0.98 | 0.97 1.00 |  | 1.01 | 1.00 1.03 |
| Personal Income (\$10,000-\$19,999) | | 1.89 | 0.95 3.76 | | 1.20 | 0.60 2.38 |
| Personal Income (\$20,000-\$29,999) | | 1.63 | 0.83 3.21 | | 1.65 | 0.87 3.11 |
| Personal Income (\$30,000-\$39,999) | | 2.56 | 1.34 4.90 | | 1.03 | 0.52 2.06 |
| Personal Income (\$40,000-\$49,999) | | 2.04 | 0.99 4.20 | | 1.08 | 0.52 2.21 |
| Personal Income (\$50,000-\$59,999) | | 2.41 | 1.21 4.80 | | 1.47 | 0.71 3.03 |
| Personal Income (\$60,000-\$69,999) | | 2.87 | 1.38 5.98 | | 1.30 | 0.57 2.98 |
| Personal Income (\$70,000-\$79,999) | | 1.65 | 0.72 3.78 | | 0.89 | 0.34 2.32 |
| Personal Income (\$80,000-\$89,999) | | 0.69 | 0.16 3.06 | | 1.02 | 0.31 3.30 |
| Personal Income (\$90,000-\$99,999) | | 4.67 | 1.84 11.85 | | 0.99 | 0.32 3.05 |
| Personal Income (\$100,000-\$149,999) | | 2.19 | 0.96 4.98 | | 0.90 | 0.35 2.30 |
| Personal Income (More than \$150,000) | | 7.25 | 3.29 16.00 | | 0.45 | 0.05 3.75 |
| Educational Level (High school graduate) |  | 0.36 | 0.13 0.94 |  | 0.85 | 0.16 4.60 |
| Educational Level (Some college credit, no degree) |  | 0.30 | 0.12 0.78 |  | 0.81 | 0.16 4.19 |
| Educational Level (Trade/Technical/Vocational education) |  | 0.38 | 0.11 1.33 |  | 0.45 | 0.07 2.89 |
| Educational Level (Associate degree) |  | 0.28 | 0.10 0.79 |  | 1.18 | 0.23 6.08 |
| Educational Level (Bachelor's degree) |  | 0.18 | 0.07 0.46 |  | 0.66 | 0.13 3.38 |
| Educational Level (Master's degree) |  | 0.26 | 0.10 0.69 |  | 0.45 | 0.08 2.46 |
| Educational Level (professional degree) |  | 0.30 | 0.06 1.42 |  | 0.50 | 0.06 4.48 |
| Educational Level (Doctorate degree) |  | 0.21 | 0.06 0.71 |  | 0.49 | 0.06 4.41 |
| Ethnicity (White) |  | 1.69 | 0.93 3.05 |  | 1.04 | 0.53 2.05 |
| Ethnicity (Hispanic, Latinx, Spanish) |  | 1.18 | 0.60 2.30 |  | 1.35 | 0.62 2.92 |
| Ethnicity (Black, African American) |  | 1.47 | 0.70 3.12 |  | 0.73 | 0.35 1.53 |
| Ethnicity (Asian) |  | 1.08 | 0.53 2.22 |  | 0.73 | 0.28 1.86 |
| Ethnicity (Native) |  | 1.48 | 0.60 3.66 |  | 1.66 | 0.69 4.01 |
| Woman |  | 0.42 | 0.30 0.60 |  | 0.74 | 0.50 1.09 |
| Non-binary/Gender fluid + Gender=other |  | 0.44 | 0.09 2.23 |  | 0.64 | 0.08 5.30 |
| **Goodness of Fit** |  |  |  |  |  |  |
| Deviance/df |  | 0.822 |  |  | 0.571 |  |
| Pearson Chi-Square/df |  | 1.034 |  |  | 1.014 |  |
| Loglikelihood |  | -761.661 |  |  | -564.438 |  |
| AIC |  | 1593.322 |  |  | 1198.877 |  |
| **Omnibus tests** |  |  |  |  |  |  |
| Chi-square |  | 269.616 |  |  | 60.175 |  |
| df |  | 34 |  |  | 34 |  |
| N |  | 1888 |  |  | 2012 |  |

Note: Participants reporting Native Hawaiian/Pacific Islander, Middle east/North Africa or Other as ethnicity are the reference group for the ethnicity variables. Personal Income < \$10,000 is the reference group for the "Personal Income" variables. Educational level < high school diploma is the reference group for the "Educational Level" variables. The models are computed with robust covariance matrix estimation.

**Table S9.**
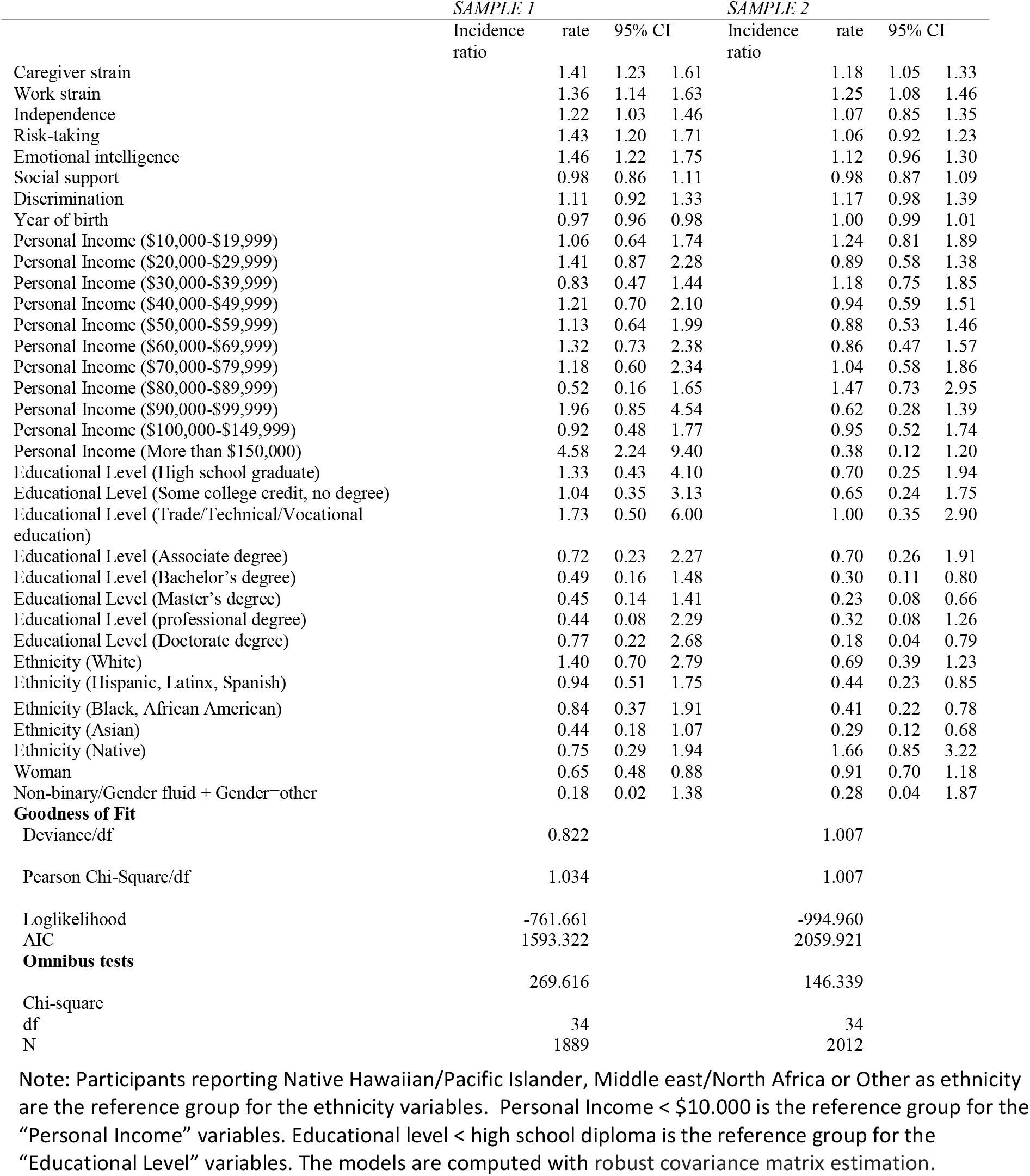
Logistic regression predicting smoking (not smoking=0, smoking=1) (with gender identity as covariate).

|  | *SAMPLE 1* |  |  | *SAMPLE 2* |  |  |
| --- | --- | --- | --- | --- | --- | --- |
|  | Incidence ratio | rate | 95% CI | Incidence ratio | rate | 95% CI |
| Caregiver strain |  | 1.41 | 1.23 1.61 |  | 1.18 | 1.05 1.33 |
| Work strain |  | 1.36 | 1.14 1.63 |  | 1.25 | 1.08 1.46 |
| Independence |  | 1.22 | 1.03 1.46 |  | 1.07 | 0.85 1.35 |
| Risk-taking |  | 1.43 | 1.20 1.71 |  | 1.06 | 0.92 1.23 |
| Emotional intelligence |  | 1.46 | 1.22 1.75 |  | 1.12 | 0.96 1.30 |
| Social support |  | 0.98 | 0.86 1.11 |  | 0.98 | 0.87 1.09 |
| Discrimination |  | 1.11 | 0.92 1.33 |  | 1.17 | 0.98 1.39 |
| Year of birth |  | 0.97 | 0.96 0.98 |  | 1.00 | 0.99 1.01 |
| Personal Income (\$10,000-\$19,999) | | 1.06 | 0.64 1.74 | | 1.24 | 0.81 1.89 |
| Personal Income (\$20,000-\$29,999) | | 1.41 | 0.87 2.28 | | 0.89 | 0.58 1.38 |
| Personal Income (\$30,000-\$39,999) | | 0.83 | 0.47 1.44 | | 1.18 | 0.75 1.85 |
| Personal Income (\$40,000-\$49,999) | | 1.21 | 0.70 2.10 | | 0.94 | 0.59 1.51 |
| Personal Income (\$50,000-\$59,999) | | 1.13 | 0.64 1.99 | | 0.88 | 0.53 1.46 |
| Personal Income (\$60,000-\$69,999) | | 1.32 | 0.73 2.38 | | 0.86 | 0.47 1.57 |
| Personal Income (\$70,000-\$79,999) | | 1.18 | 0.60 2.34 | | 1.04 | 0.58 1.86 |
| Personal Income (\$80,000-\$89,999) | | 0.52 | 0.16 1.65 | | 1.47 | 0.73 2.95 |
| Personal Income (\$90,000-\$99,999) | | 1.96 | 0.85 4.54 | | 0.62 | 0.28 1.39 |
| Personal Income (\$100,000-\$149,999) | | 0.92 | 0.48 1.77 | | 0.95 | 0.52 1.74 |
| Personal Income (More than \$150,000) | | 4.58 | 2.24 9.40 | | 0.38 | 0.12 1.20 |
| Educational Level (High school graduate) |  | 1.33 | 0.43 4.10 |  | 0.70 | 0.25 1.94 |
| Educational Level (Some college credit, no degree) |  | 1.04 | 0.35 3.13 |  | 0.65 | 0.24 1.75 |
| Educational Level (Trade/Technical/Vocational education) |  | 1.73 | 0.50 6.00 |  | 1.00 | 0.35 2.90 |
| Educational Level (Associate degree) |  | 0.72 | 0.23 2.27 |  | 0.70 | 0.26 1.91 |
| Educational Level (Bachelor's degree) |  | 0.49 | 0.16 1.48 |  | 0.30 | 0.11 0.80 |
| Educational Level (Master's degree) |  | 0.45 | 0.14 1.41 |  | 0.23 | 0.08 0.66 |
| Educational Level (professional degree) |  | 0.44 | 0.08 2.29 |  | 0.32 | 0.08 1.26 |
| Educational Level (Doctorate degree) |  | 0.77 | 0.22 2.68 |  | 0.18 | 0.04 0.79 |
| Ethnicity (White) |  | 1.40 | 0.70 2.79 |  | 0.69 | 0.39 1.23 |
| Ethnicity (Hispanic, Latinx, Spanish) |  | 0.94 | 0.51 1.75 |  | 0.44 | 0.23 0.85 |
| Ethnicity (Black, African American) |  | 0.84 | 0.37 1.91 |  | 0.41 | 0.22 0.78 |
| Ethnicity (Asian) |  | 0.44 | 0.18 1.07 |  | 0.29 | 0.12 0.68 |
| Ethnicity (Native) |  | 0.75 | 0.29 1.94 |  | 1.66 | 0.85 3.22 |
| Woman |  | 0.65 | 0.48 0.88 |  | 0.91 | 0.70 1.18 |
| Non-binary/Gender fluid + Gender=other |  | 0.18 | 0.02 1.38 |  | 0.28 | 0.04 1.87 |
| **Goodness of Fit** |  |  |  |  |  |  |
| Deviance/df |  | 0.822 |  |  | 1.007 |  |
| Pearson Chi-Square/df |  | 1.034 |  |  | 1.007 |  |
| Loglikelihood |  | -761.661 |  |  | -994.960 |  |
| AIC |  | 1593.322 |  |  | 2059.921 |  |
| **Omnibus tests** |  |  |  |  |  |  |
| Chi-square |  | 269.616 |  |  | 146.339 |  |
| df |  | 34 |  |  | 34 |  |
| N |  | 1889 |  |  | 2012 |  |

Note: Participants reporting Native Hawaiian/Pacific Islander, Middle east/North Africa or Other as ethnicity are the reference group for the ethnicity variables. Personal Income < \$10,000 is the reference group for the "Personal Income" variables. Educational level < high school diploma is the reference group for the "Educational Level" variables. The models are computed with robust covariance matrix estimation.

**Table S10.** Logistic regression predicting binge drinking (less than monthly=0, monthly, weekly, and daily=1) (with gender identity as covariate).

|  | *SAMPLE 1* |  |  | *SAMPLE 2* |  |  |
| --- | --- | --- | --- | --- | --- | --- |
|  | Incidence ratio | rate | 95% CI | Incidence ratio | rate | 95% CI |
| Caregiver strain |  | 0.92 | 0.80 1.05 |  | 0.91 | 0.80 1.04 |
| Work strain |  | 1.28 | 1.09 1.52 |  | 1.03 | 0.88 1.21 |
| Independence |  | 1.08 | 0.93 1.24 |  | 1.19 | 0.94 1.49 |
| Risk-taking |  | 1.53 | 1.31 1.78 |  | 1.25 | 1.08 1.45 |
| Emotional intelligence |  | 1.18 | 1.00 1.40 |  | 1.04 | 0.89 1.22 |
| Social support |  | 1.01 | 0.90 1.13 |  | 0.87 | 0.78 0.97 |
| Discrimination |  | 1.17 | 0.99 1.37 |  | 1.12 | 0.93 1.34 |
| Year of birth |  | 1.02 | 1.01 1.03 |  | 1.03 | 1.02 1.04 |
| Personal Income (\$10,000-\$19,999) | | 1.03 | 0.65 1.62 | | 1.27 | 0.78 2.04 |
| Personal Income (\$20,000-\$29,999) | | 1.15 | 0.73 1.82 | | 1.94 | 1.23 3.06 |
| Personal Income (\$30,000-\$39,999) | | 1.20 | 0.76 1.90 | | 1.33 | 0.81 2.17 |
| Personal Income (\$40,000-\$49,999) | | 1.31 | 0.81 2.13 | | 1.79 | 1.08 2.96 |
| Personal Income (\$50,000-\$59,999) | | 0.93 | 0.56 1.53 | | 1.37 | 0.80 2.36 |
| Personal Income (\$60,000-\$69,999) | | 1.21 | 0.71 2.06 | | 1.98 | 1.11 3.55 |
| Personal Income (\$70,000-\$79,999) | | 0.83 | 0.45 1.54 | | 1.44 | 0.78 2.65 |
| Personal Income (\$80,000-\$89,999) | | 1.17 | 0.53 2.58 | | 1.15 | 0.52 2.57 |
| Personal Income (\$90,000-\$99,999) | | 1.68 | 0.73 3.86 | | 2.29 | 1.13 4.62 |
| Personal Income (\$100,000-\$149,999) | | 1.16 | 0.63 2.12 | | 1.93 | 1.05 3.57 |
| Personal Income (More than \$150,000) | | 2.08 | 1.04 4.16 | | 2.42 | 0.99 5.88 |
| Educational Level (High school graduate) |  | 1.20 | 0.40 3.59 |  | 0.50 | 0.15 1.65 |
| Educational Level (Some college credit, no degree) |  | 1.67 | 0.58 4.79 |  | 0.90 | 0.29 2.82 |
| Educational Level (Trade/Technical/Vocational education) |  | 2.37 | 0.71 7.93 |  | 0.85 | 0.25 2.92 |
| Educational Level (Associate degree) |  | 1.36 | 0.45 4.12 |  | 0.65 | 0.20 2.09 |
| Educational Level (Bachelor's degree) |  | 1.52 | 0.53 4.36 |  | 0.63 | 0.20 1.97 |
| Educational Level (Master's degree) |  | 1.80 | 0.62 5.28 |  | 0.57 | 0.18 1.86 |
| Educational Level (professional degree) |  | 3.24 | 0.77 13.57 |  | 0.58 | 0.14 2.50 |
| Educational Level (Doctorate degree) |  | 1.76 | 0.52 5.91 |  | 0.65 | 0.15 2.73 |
| Ethnicity (White) |  | 1.89 | 1.08 3.30 |  | 0.92 | 0.53 1.60 |
| Ethnicity (Hispanic, Latinx, Spanish) |  | 1.64 | 0.96 2.79 |  | 0.83 | 0.47 1.46 |
| Ethnicity (Black, African American) |  | 0.75 | 0.36 1.54 |  | 1.04 | 0.56 1.92 |
| Ethnicity (Asian) |  | 0.58 | 0.30 1.12 |  | 0.54 | 0.27 1.08 |
| Ethnicity (Native) |  | 0.46 | 0.19 1.09 |  | 0.79 | 0.38 1.64 |
| Woman |  | 0.57 | 0.44 0.74 |  |  |  |
| Non-binary/Gender fluid + Gender=other |  | 0.53 | 0.18 1.56 |  |  |  |
| Gender Dummy (woman=1) |  |  |  |  | 0.78 | 0.60 1.03 |
| **Goodness of Fit** |  |  |  |  |  |  |
| Deviance/df |  | 1.001 |  |  | 0.571 |  |
| Pearson Chi-Square/df |  | 1.017 |  |  | 1.014 |  |
| Loglikelihood |  | -923.706 |  |  | -564.438 |  |
| AIC |  | 1917.411 |  |  | 1198.877 |  |
| **Omnibus tests** |  | 212.582 |  |  | 60.175 |  |
| Chi-square |  |  |  |  |  |  |
| df |  | 34 |  |  | 34 |  |
| N |  | 1888 |  |  | 2000 |  |

Note: Participants reporting Native Hawaiian/Pacific Islander, Middle east/North Africa or Other as ethnicity are the reference group for the ethnicity variables. Personal Income < \$10,000 is the reference group for the "Personal Income" variables. Educational level < high school diploma is the reference group for the "Educational Level" variables. It was not possible to estimate the effect of "Non-binary/Gender fluid + Gender=other" in Sample 2. Instead, we included a dichotomous gender variable (man=0, woman=1). The models are computed with robust covariance matrix estimation.

**Table S11.**
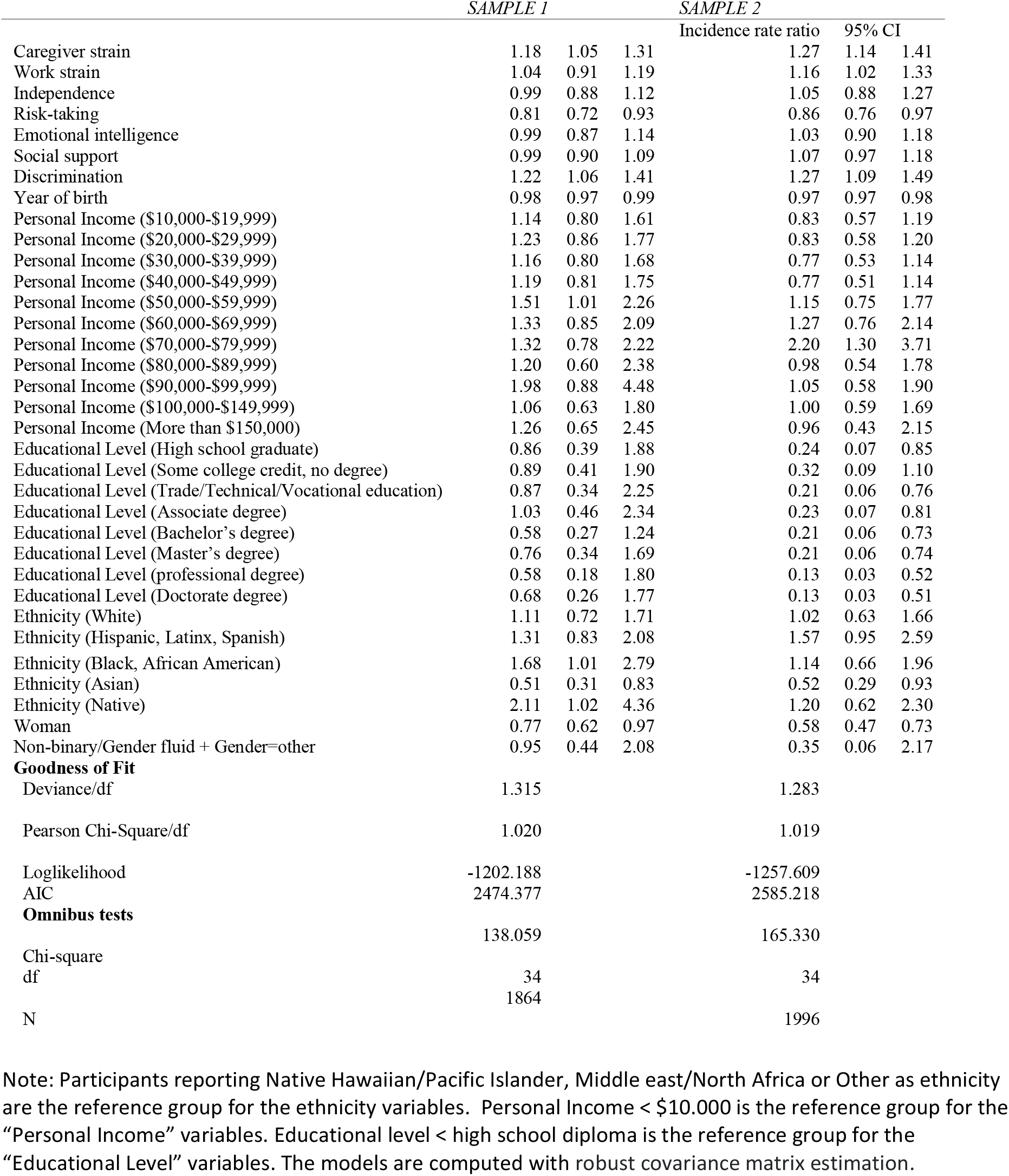
Logistic regression predicting overweight (BMI<25=0, BMI≥25 =1) (with gender identity as covariate).

**Table S12.** Negative binomial regression predicting number of days with poor physical health (during past 30 days) (with sex as covariate).

|  | *SAMPLE 1* |  |  | *SAMPLE 2* |  |  |
| --- | --- | --- | --- | --- | --- | --- |
|  | Incidence ratio | rate | 95% CI | Incidence ratio | rate | 95% CI |
| Caregiver strain |  | 1.20 | 1.12 1.29 |  | 1.11 | 1.04 1.18 |
| Work strain |  | 0.96 | 0.87 1.05 |  | 1.02 | 0.94 1.11 |
| Independence |  | 0.93 | 0.85 1.01 |  | 0.84 | 0.75 0.95 |
| Risk-taking |  | 0.95 | 0.87 1.04 |  | 1.00 | 0.92 1.08 |
| Emotional intelligence |  | 0.93 | 0.84 1.03 |  | 0.93 | 0.85 1.01 |
| Social support |  | 0.95 | 0.89 1.02 |  | 0.94 | 0.89 1.00 |
| Discrimination |  | 1.46 | 1.34 1.59 |  | 1.43 | 1.30 1.57 |
| Year of birth |  | 0.99 | 0.98 1.00 |  | 0.99 | 0.98 0.99 |
| Personal Income (\$10,000-\$19,999) | | 1.01 | 0.79 1.29 | | 1.07 | 0.86 1.34 |
| Personal Income (\$20,000-\$29,999) | | 0.94 | 0.73 1.21 | | 1.01 | 0.82 1.25 |
| Personal Income (\$30,000-\$39,999) | | 0.83 | 0.64 1.09 | | 0.87 | 0.68 1.12 |
| Personal Income (\$40,000-\$49,999) | | 1.00 | 0.75 1.33 | | 0.71 | 0.56 0.91 |
| Personal Income (\$50,000-\$59,999) | | 0.92 | 0.67 1.26 | | 0.68 | 0.50 0.93 |
| Personal Income (\$60,000-\$69,999) | | 0.83 | 0.60 1.15 | | 0.70 | 0.52 0.95 |
| Personal Income (\$70,000-\$79,999) | | 0.81 | 0.54 1.22 | | 0.62 | 0.44 0.86 |
| Personal Income (\$80,000-\$89,999) | | 0.66 | 0.42 1.03 | | 0.45 | 0.32 0.63 |
| Personal Income (\$90,000-\$99,999) | | 1.03 | 0.55 1.91 | | 0.76 | 0.51 1.15 |
| Personal Income (\$100,000-\$149,999) | | 0.77 | 0.51 1.16 | | 0.68 | 0.48 0.96 |
| Personal Income (More than \$150,000) | | 1.26 | 0.81 1.95 | | 0.55 | 0.29 1.03 |
| Educational Level (High school graduate) |  | 1.04 | 0.64 1.70 |  | 0.88 | 0.50 1.55 |
| Educational Level (Some college credit, no degree) |  | 0.88 | 0.55 1.40 |  | 1.11 | 0.65 1.90 |
| Educational Level (Trade/Technical/Vocational education) |  | 0.87 | 0.49 1.55 |  | 0.84 | 0.46 1.51 |
| Educational Level (Associate degree) |  | 0.80 | 0.48 1.32 |  | 1.00 | 0.58 1.74 |
| Educational Level (Bachelor's degree) |  | 0.76 | 0.47 1.20 |  | 0.85 | 0.50 1.46 |
| Educational Level (Master's degree) |  | 0.90 | 0.54 1.48 |  | 0.85 | 0.48 1.48 |
| Educational Level (professional degree) |  | 0.72 | 0.25 2.10 |  | 0.92 | 0.44 1.96 |
| Educational Level (Doctorate degree) |  | 0.69 | 0.38 1.26 |  | 0.69 | 0.36 1.33 |
| Ethnicity (White) |  | 1.07 | 0.80 1.43 |  | 0.96 | 0.74 1.25 |
| Ethnicity (Hispanic, Latinx, Spanish) |  | 1.01 | 0.72 1.40 |  | 0.97 | 0.71 1.32 |
| Ethnicity (Black, African American) |  | 0.97 | 0.70 1.35 |  | 0.87 | 0.66 1.14 |
| Ethnicity (Asian) |  | 0.86 | 0.62 1.19 |  | 0.88 | 0.62 1.26 |
| Ethnicity (Native) |  | 0.91 | 0.67 1.23 |  | 1.62 | 1.11 2.38 |
| Sex (male=0, female=1) |  | 0.94 | 0.81 1.10 |  | 0.93 | 0.80 1.08 |
| **Goodness of Fit** |  |  |  |  |  |  |
| Deviance/df |  | 1.114 |  |  | 1.120 |  |
| Pearson Chi-Square/df |  | 1.240 |  |  | 1.159 |  |
| Loglikelihood |  | -4827.875 |  |  | -5152.368 |  |
| AIC |  | 9725.751 |  |  | 10374.737 |  |
| **Omnibus tests** |  |  |  |  |  |  |
| Chi-square |  | 179.945 |  |  | 200.473 |  |
| df |  | 33 |  |  | 33 |  |
| N |  | 1874 |  |  | 2005 |  |

Note: Participants reporting Native Hawaiian/Pacific Islander, Middle east/North Africa or Other as ethnicity are the reference group for the ethnicity variables. Personal Income < \$10,000 is the reference group for the "Personal Income" variables. Educational level < high school diploma is the reference group for the "Educational Level" variables. The models are computed with robust covariance matrix estimation.

**Table S13.** Negative Binomial regression predicting number of days with poor mental health (during past 30 days) (with sex as covariate).

|  | *SAMPLE 1* |  |  | *SAMPLE 2* |  |  |
| --- | --- | --- | --- | --- | --- | --- |
|  | Incidence ratio | rate | 95% CI | Incidence ratio | rate | 95% CI |
| Caregiver strain |  | 1.21 | 1.14 1.27 |  | 1.20 | 1.14 1.27 |
| Work strain |  | 1.03 | 0.96 1.11 |  | 1.12 | 1.04 1.21 |
| Independence |  | 0.97 | 0.91 1.04 |  | 0.83 | 0.74 0.94 |
| Risk-taking |  | 0.86 | 0.80 0.92 |  | 0.99 | 0.91 1.08 |
| Emotional intelligence |  | 0.97 | 0.89 1.05 |  | 0.86 | 0.78 0.93 |
| Social support |  | 0.86 | 0.81 0.90 |  | 0.81 | 0.76 0.86 |
| Discrimination |  | 1.44 | 1.34 1.53 |  | 1.35 | 1.24 1.48 |
| Year of birth |  | 1.02 | 1.01 1.02 |  | 1.02 | 1.01 1.02 |
| Personal Income (\$10,000-\$19,999) | | 0.98 | 0.82 1.17 | | 1.03 | 0.84 1.26 |
| Personal Income (\$20,000-\$29,999) | | 0.74 | 0.61 0.90 | | 0.89 | 0.72 1.10 |
| Personal Income (\$30,000-\$39,999) | | 0.78 | 0.63 0.95 | | 0.85 | 0.68 1.05 |
| Personal Income (\$40,000-\$49,999) | | 0.76 | 0.61 0.95 | | 0.64 | 0.50 0.80 |
| Personal Income (\$50,000-\$59,999) | | 0.81 | 0.65 1.01 | | 0.51 | 0.38 0.68 |
| Personal Income (\$60,000-\$69,999) | | 0.71 | 0.56 0.91 | | 0.55 | 0.41 0.74 |
| Personal Income (\$70,000-\$79,999) | | 0.69 | 0.49 0.96 | | 0.55 | 0.40 0.76 |
| Personal Income (\$80,000-\$89,999) | | 0.77 | 0.53 1.10 | | 0.77 | 0.47 1.26 |
| Personal Income (\$90,000-\$99,999) | | 0.97 | 0.65 1.45 | | 0.63 | 0.39 1.01 |
| Personal Income (\$100,000-\$149,999) | | 0.59 | 0.42 0.82 | | 0.63 | 0.44 0.89 |
| Personal Income (More than \$150,000) | | 0.93 | 0.62 1.38 | | 0.44 | 0.25 0.75 |
| Educational Level (High school graduate) |  | 1.07 | 0.72 1.58 |  | 1.50 | 0.87 2.61 |
| Educational Level (Some college credit, no degree) |  | 1.05 | 0.72 1.54 |  | 1.67 | 0.98 2.85 |
| Educational Level (Trade/Technical/Vocational education) |  | 1.16 | 0.73 1.82 |  | 1.90 | 1.03 3.51 |
| Educational Level (Associate degree) |  | 1.02 | 0.67 1.54 |  | 1.61 | 0.93 2.76 |
| Educational Level (Bachelor's degree) |  | 0.89 | 0.61 1.29 |  | 1.53 | 0.90 2.59 |
| Educational Level (Master's degree) |  | 1.08 | 0.72 1.61 |  | 1.61 | 0.92 2.80 |
| Educational Level (professional degree) |  | 0.73 | 0.37 1.44 |  | 1.83 | 0.90 3.74 |
| Educational Level (Doctorate degree) |  | 0.93 | 0.58 1.49 |  | 1.73 | 0.86 3.47 |
| Ethnicity (White) |  | 1.59 | 1.29 1.96 |  | 1.32 | 1.01 1.73 |
| Ethnicity (Hispanic, Latinx, Spanish) |  | 1.07 | 0.86 1.33 |  | 1.11 | 0.83 1.47 |
| Ethnicity (Black, African American) |  | 1.31 | 1.03 1.67 |  | 0.94 | 0.72 1.24 |
| Ethnicity (Asian) |  | 1.16 | 0.91 1.48 |  | 1.09 | 0.76 1.56 |
| Ethnicity (Native) |  | 1.06 | 0.80 1.41 |  | 1.89 | 1.27 2.80 |
| Sex (male=0, female=1) |  | 0.94 | 0.83 1.06 |  | 1.21 | 1.05 1.40 |
| **Goodness of Fit** |  |  |  |  |  |  |
| Deviance/df |  | 1.172 |  |  | 1.131 |  |
| Pearson Chi-Square/df |  | 1.019 |  |  | 1.104 |  |
| Loglikelihood |  | -5645.574 |  |  | -5566.070 |  |
| AIC |  | 11361.148 |  |  | 11202.139 |  |
| **Omnibus tests** |  |  |  |  |  |  |
| Chi-square |  | 284.801 |  |  | 315.270 |  |
| df |  | 33 |  |  | 33 |  |
| N |  | 1896 |  |  | 2004 |  |

Note: Participants reporting Native Hawaiian/Pacific Islander, Middle east/North Africa or Other as ethnicity are the reference group for the ethnicity variables. Personal Income < \$10,000 is the reference group for the "Personal Income" variables. Educational level < high school diploma is the reference group for the "Educational Level" variables. The models are computed with robust covariance matrix estimation.

**Table S14.** Negative binomial regression predicting number of days where poor mental or physical health prevented the respondent from doing usual activities (during past 30 days) (with sex as covariate).

|  | *SAMPLE 1* |  |  | *SAMPLE 2* |  |  |
| --- | --- | --- | --- | --- | --- | --- |
|  | Incidence ratio | rate | 95% CI | Incidence ratio | rate | 95% CI |
| Caregiver strain |  | 1.13 | 1.04 1.21 |  | 1.11 | 1.03 1.19 |
| Work strain |  | 0.88 | 0.80 0.96 |  | 0.99 | 0.90 1.08 |
| Independence |  | 1.02 | 0.94 1.12 |  | 0.87 | 0.77 1.00 |
| Risk-taking |  | 0.93 | 0.85 1.01 |  | 1.02 | 0.94 1.12 |
| Emotional intelligence |  | 0.98 | 0.88 1.10 |  | 0.92 | 0.84 1.02 |
| Social support |  | 0.92 | 0.86 0.99 |  | 0.87 | 0.81 0.94 |
| Discrimination |  | 1.76 | 1.61 1.93 |  | 1.56 | 1.40 1.73 |
| Year of birth |  | 1.01 | 1.00 1.01 |  | 1.00 | 1.00 1.01 |
| Personal Income (\$10,000-\$19,999) | | 1.01 | 0.80 1.27 | | 0.91 | 0.70 1.17 |
| Personal Income (\$20,000-\$29,999) | | 0.89 | 0.70 1.14 | | 0.93 | 0.73 1.18 |
| Personal Income (\$30,000-\$39,999) | | 0.75 | 0.56 1.01 | | 0.78 | 0.59 1.02 |
| Personal Income (\$40,000-\$49,999) | | 0.91 | 0.68 1.22 | | 0.65 | 0.48 0.87 |
| Personal Income (\$50,000-\$59,999) | | 0.76 | 0.56 1.03 | | 0.52 | 0.37 0.73 |
| Personal Income (\$60,000-\$69,999) | | 0.63 | 0.44 0.89 | | 0.67 | 0.47 0.97 |
| Personal Income (\$70,000-\$79,999) | | 0.66 | 0.42 1.04 | | 0.62 | 0.43 0.89 |
| Personal Income (\$80,000-\$89,999) | | 1.01 | 0.59 1.74 | | 0.65 | 0.40 1.04 |
| Personal Income (\$90,000-\$99,999) | | 0.99 | 0.51 1.93 | | 0.62 | 0.40 0.95 |
| Personal Income (\$100,000-\$149,999) | | 0.90 | 0.57 1.41 | | 0.70 | 0.46 1.06 |
| Personal Income (More than \$150,000) | | 1.48 | 0.88 2.50 | | 0.75 | 0.38 1.47 |
| Educational Level (High school graduate) |  | 1.08 | 0.65 1.79 |  | 1.00 | 0.52 1.93 |
| Educational Level (Some college credit, no degree) |  | 0.86 | 0.53 1.41 |  | 1.13 | 0.61 2.10 |
| Educational Level (Trade/Technical/Vocational education) |  | 1.33 | 0.75 2.39 |  | 0.82 | 0.40 1.66 |
| Educational Level (Associate degree) |  | 0.84 | 0.49 1.42 |  | 0.96 | 0.51 1.82 |
| Educational Level (Bachelor's degree) |  | 0.88 | 0.54 1.43 |  | 0.78 | 0.42 1.44 |
| Educational Level (Master's degree) |  | 0.74 | 0.44 1.25 |  | 0.71 | 0.37 1.34 |
| Educational Level (professional degree) |  | 0.25 | 0.11 0.58 |  | 0.53 | 0.23 1.25 |
| Educational Level (Doctorate degree) |  | 0.67 | 0.37 1.20 |  | 0.61 | 0.28 1.33 |
| Ethnicity (White) |  | 1.15 | 0.87 1.53 |  | 0.90 | 0.65 1.23 |
| Ethnicity (Hispanic, Latinx, Spanish) |  | 0.90 | 0.67 1.20 |  | 1.01 | 0.71 1.43 |
| Ethnicity (Black, African American) |  | 1.03 | 0.74 1.43 |  | 0.77 | 0.55 1.08 |
| Ethnicity (Asian) |  | 0.78 | 0.57 1.06 |  | 0.88 | 0.57 1.34 |
| Ethnicity (Native) |  | 0.93 | 0.62 1.40 |  | 2.04 | 1.26 3.30 |
| Sex (male=0, female=1) |  | 0.88 | 0.74 1.04 |  | 1.11 | 0.94 1.31 |
| **Goodness of Fit** |  |  |  |  |  |  |
| Deviance/df |  | 1.081 |  |  | 1.057 |  |
| Pearson Chi-Square/df |  | 1.170 |  |  | 1.143 |  |
| Loglikelihood |  | -4564.397 |  |  | -4630.156 |  |
| AIC |  | 9198.795 |  |  | 9330.312 |  |
| **Omnibus tests** |  |  |  |  |  |  |
| Chi-square |  | 246.377 |  |  | 223.091 |  |
| df |  | 33 |  |  | 33 |  |
| N |  | 1872 |  |  | 2004 |  |

Note: Participants reporting Native Hawaiian/Pacific Islander, Middle east/North Africa or Other as ethnicity are the reference group for the ethnicity variables. Personal Income < \$10,000 is the reference group for the "Personal Income" variables. Educational level < high school diploma is the reference group for the "Educational Level" variables. The models are computed with robust covariance matrix estimation.

**Table S15.**
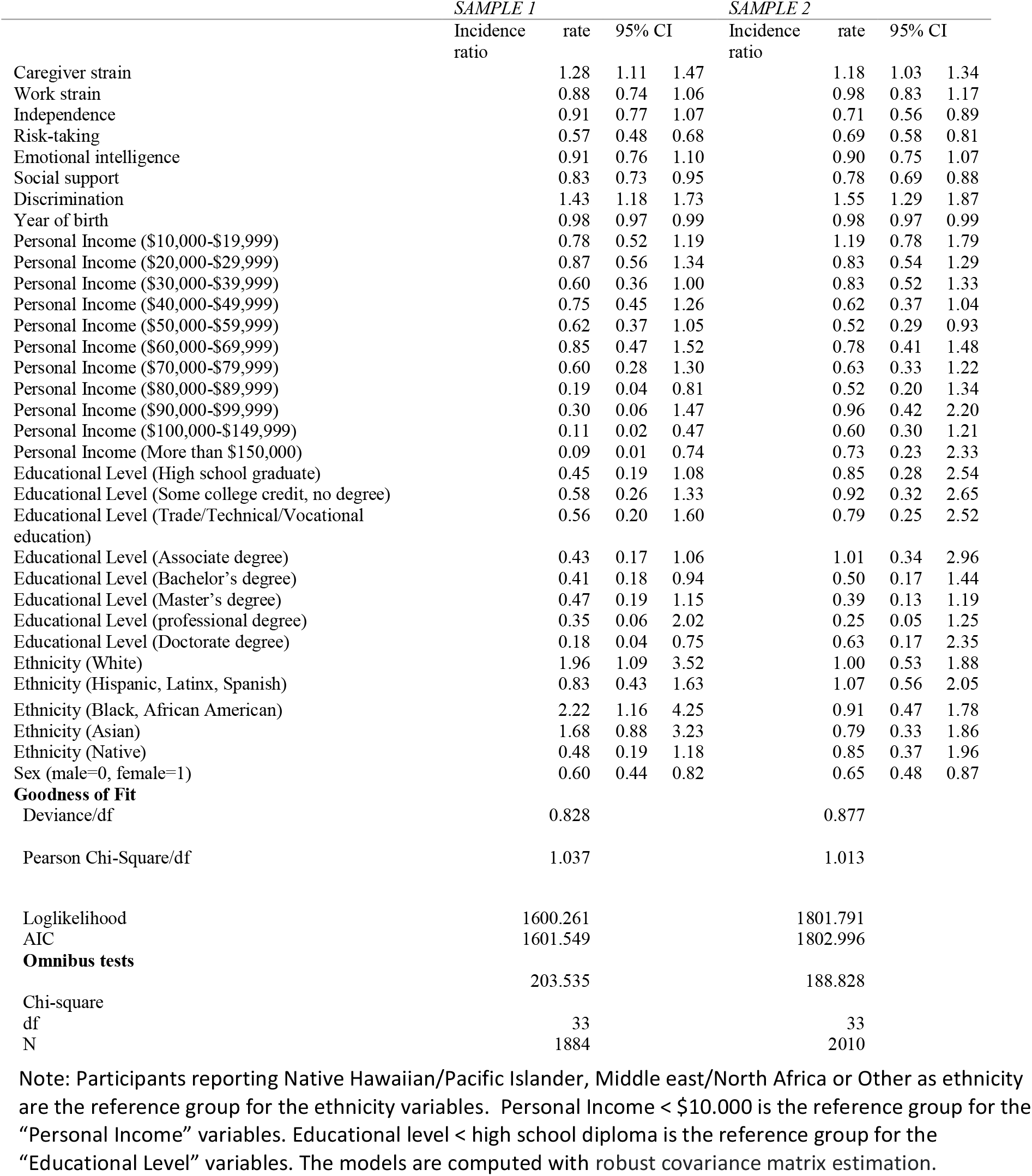
Logistic regression predicting general health status (excellent, very good, good= 0, fair, poor= 1) (with sex as covariate).

|  | *SAMPLE 1* |  |  | *SAMPLE 2* |  |  |
| --- | --- | --- | --- | --- | --- | --- |
|  | Incidence ratio | rate | 95% CI | Incidence ratio | rate | 95% CI |
| Caregiver strain |  | 1.28 | 1.11 1.47 |  | 1.18 | 1.03 1.34 |
| Work strain |  | 0.88 | 0.74 1.06 |  | 0.98 | 0.83 1.17 |
| Independence |  | 0.91 | 0.77 1.07 |  | 0.71 | 0.56 0.89 |
| Risk-taking |  | 0.57 | 0.48 0.68 |  | 0.69 | 0.58 0.81 |
| Emotional intelligence |  | 0.91 | 0.76 1.10 |  | 0.90 | 0.75 1.07 |
| Social support |  | 0.83 | 0.73 0.95 |  | 0.78 | 0.69 0.88 |
| Discrimination |  | 1.43 | 1.18 1.73 |  | 1.55 | 1.29 1.87 |
| Year of birth |  | 0.98 | 0.97 0.99 |  | 0.98 | 0.97 0.99 |
| Personal Income (\$10,000-\$19,999) | | 0.78 | 0.52 1.19 | | 1.19 | 0.78 1.79 |
| Personal Income (\$20,000-\$29,999) | | 0.87 | 0.56 1.34 | | 0.83 | 0.54 1.29 |
| Personal Income (\$30,000-\$39,999) | | 0.60 | 0.36 1.00 | | 0.83 | 0.52 1.33 |
| Personal Income (\$40,000-\$49,999) | | 0.75 | 0.45 1.26 | | 0.62 | 0.37 1.04 |
| Personal Income (\$50,000-\$59,999) | | 0.62 | 0.37 1.05 | | 0.52 | 0.29 0.93 |
| Personal Income (\$60,000-\$69,999) | | 0.85 | 0.47 1.52 | | 0.78 | 0.41 1.48 |
| Personal Income (\$70,000-\$79,999) | | 0.60 | 0.28 1.30 | | 0.63 | 0.33 1.22 |
| Personal Income (\$80,000-\$89,999) | | 0.19 | 0.04 0.81 | | 0.52 | 0.20 1.34 |
| Personal Income (\$90,000-\$99,999) | | 0.30 | 0.06 1.47 | | 0.96 | 0.42 2.20 |
| Personal Income (\$100,000-\$149,999) | | 0.11 | 0.02 0.47 | | 0.60 | 0.30 1.21 |
| Personal Income (More than \$150,000) | | 0.09 | 0.01 0.74 | | 0.73 | 0.23 2.33 |
| Educational Level (High school graduate) |  | 0.45 | 0.19 1.08 |  | 0.85 | 0.28 2.54 |
| Educational Level (Some college credit, no degree) |  | 0.58 | 0.26 1.33 |  | 0.92 | 0.32 2.65 |
| Educational Level (Trade/Technical/Vocational education) |  | 0.56 | 0.20 1.60 |  | 0.79 | 0.25 2.52 |
| Educational Level (Associate degree) |  | 0.43 | 0.17 1.06 |  | 1.01 | 0.34 2.96 |
| Educational Level (Bachelor's degree) |  | 0.41 | 0.18 0.94 |  | 0.50 | 0.17 1.44 |
| Educational Level (Master's degree) |  | 0.47 | 0.19 1.15 |  | 0.39 | 0.13 1.19 |
| Educational Level (professional degree) |  | 0.35 | 0.06 2.02 |  | 0.25 | 0.05 1.25 |
| Educational Level (Doctorate degree) |  | 0.18 | 0.04 0.75 |  | 0.63 | 0.17 2.35 |
| Ethnicity (White) |  | 1.96 | 1.09 3.52 |  | 1.00 | 0.53 1.88 |
| Ethnicity (Hispanic, Latinx, Spanish) |  | 0.83 | 0.43 1.63 |  | 1.07 | 0.56 2.05 |
| Ethnicity (Black, African American) |  | 2.22 | 1.16 4.25 |  | 0.91 | 0.47 1.78 |
| Ethnicity (Asian) |  | 1.68 | 0.88 3.23 |  | 0.79 | 0.33 1.86 |
| Ethnicity (Native) |  | 0.48 | 0.19 1.18 |  | 0.85 | 0.37 1.96 |
| Sex (male=0, female=1) |  | 0.60 | 0.44 0.82 |  | 0.65 | 0.48 0.87 |
| **Goodness of Fit** |  |  |  |  |  |  |
| Deviance/df |  | 0.828 |  |  | 0.877 |  |
| Pearson Chi-Square/df |  | 1.037 |  |  | 1.013 |  |
| Loglikelihood |  | 1600.261 |  |  | 1801.791 |  |
| AIC |  | 1601.549 |  |  | 1802.996 |  |
| **Omnibus tests** |  |  |  |  |  |  |
| Chi-square |  | 203.535 |  |  | 188.828 |  |
| df |  | 33 |  |  | 33 |  |
| N |  | 1884 |  |  | 2010 |  |

Note: Participants reporting Native Hawaiian/Pacific Islander, Middle east/North Africa or Other as ethnicity are the reference group for the ethnicity variables. Personal Income < \$10.000 is the reference group for the "Personal Income" variables. Educational level < high school diploma is the reference group for the "Educational Level" variables. The models are computed with robust covariance matrix estimation.

**Table S16.** Logistic regression predicting smoking (not smoking=0, smoking=1) (with sex as covariate).

|  | *SAMPLE 1* |  |  | *SAMPLE 2* |  |  |
| --- | --- | --- | --- | --- | --- | --- |
|  | Incidence ratio | rate | 95% CI | Incidence ratio | rate | 95% CI |
| Caregiver strain |  | 1.42 | 1.24 1.62 |  | 1.18 | 1.05 1.33 |
| Work strain |  | 1.37 | 1.15 1.64 |  | 1.26 | 1.08 1.46 |
| Independence |  | 1.22 | 1.03 1.46 |  | 1.07 | 0.85 1.35 |
| Risk-taking |  | 1.45 | 1.21 1.72 |  | 1.05 | 0.91 1.21 |
| Emotional intelligence |  | 1.46 | 1.22 1.75 |  | 1.12 | 0.96 1.30 |
| Social support |  | 0.98 | 0.86 1.12 |  | 0.97 | 0.87 1.09 |
| Discrimination |  | 1.09 | 0.90 1.31 |  | 1.16 | 0.97 1.38 |
| Year of birth |  | 0.97 | 0.96 0.98 |  | 1.00 | 0.99 1.01 |
| Personal Income (\$10,000-\$19,999) | | 1.04 | 0.63 1.71 | | 1.22 | 0.80 1.86 |
| Personal Income (\$20,000-\$29,999) | | 1.41 | 0.87 2.28 | | 0.89 | 0.58 1.37 |
| Personal Income (\$30,000-\$39,999) | | 0.82 | 0.47 1.43 | | 1.16 | 0.74 1.82 |
| Personal Income (\$40,000-\$49,999) | | 1.20 | 0.69 2.07 | | 0.94 | 0.59 1.51 |
| Personal Income (\$50,000-\$59,999) | | 1.10 | 0.62 1.94 | | 0.88 | 0.54 1.46 |
| Personal Income (\$60,000-\$69,999) | | 1.31 | 0.72 2.37 | | 0.86 | 0.47 1.57 |
| Personal Income (\$70,000-\$79,999) | | 1.17 | 0.59 2.31 | | 1.04 | 0.58 1.85 |
| Personal Income (\$80,000-\$89,999) | | 0.50 | 0.16 1.59 | | 1.47 | 0.73 2.95 |
| Personal Income (\$90,000-\$99,999) | | 1.94 | 0.84 4.48 | | 0.64 | 0.29 1.43 |
| Personal Income (\$100,000-\$149,999) | | 0.90 | 0.47 1.73 | | 0.95 | 0.52 1.75 |
| Personal Income (More than \$150,000) | | 4.84 | 2.36 9.94 | | 0.38 | 0.12 1.21 |
| Educational Level (High school graduate) |  | 1.34 | 0.43 4.12 |  | 0.69 | 0.25 1.90 |
| Educational Level (Some college credit, no degree) |  | 1.02 | 0.34 3.08 |  | 0.65 | 0.24 1.75 |
| Educational Level (Trade/Technical/Vocational education) |  | 1.74 | 0.50 6.06 |  | 1.00 | 0.35 2.89 |
| Educational Level (Associate degree) |  | 0.72 | 0.23 2.28 |  | 0.70 | 0.26 1.91 |
| Educational Level (Bachelor's degree) |  | 0.49 | 0.16 1.47 |  | 0.30 | 0.11 0.79 |
| Educational Level (Master's degree) |  | 0.45 | 0.14 1.40 |  | 0.23 | 0.08 0.66 |
| Educational Level (professional degree) |  | 0.42 | 0.08 2.25 |  | 0.32 | 0.08 1.26 |
| Educational Level (Doctorate degree) |  | 0.75 | 0.22 2.60 |  | 0.18 | 0.04 0.78 |
| Ethnicity (White) |  | 1.31 | 0.65 2.63 |  | 0.71 | 0.40 1.25 |
| Ethnicity (Hispanic, Latinx, Spanish) |  | 0.90 | 0.48 1.68 |  | 0.46 | 0.24 0.87 |
| Ethnicity (Black, African American) |  | 0.78 | 0.34 1.79 |  | 0.43 | 0.23 0.81 |
| Ethnicity (Asian) |  | 0.42 | 0.17 1.04 |  | 0.30 | 0.13 0.70 |
| Ethnicity (Native) |  | 0.74 | 0.29 1.91 |  | 1.65 | 0.85 3.20 |
| Sex (male=0, female=1) |  | 0.66 | 0.48 0.89 |  | 0.89 | 0.69 1.16 |
| **Goodness of Fit** |  |  |  |  |  |  |
| Deviance/df |  | 0.824 |  |  | 1.007 |  |
| Pearson Chi-Square/df |  | 1.042 |  |  | 1.008 |  |
| Loglikelihood |  | -761.542 |  |  | -994.497 |  |
| AIC |  | 1591.084 |  |  | 2056.993 |  |
| **Omnibus tests** |  |  |  |  |  |  |
| Chi-square |  | 267.437 |  |  | 143.257 |  |
| df |  | 33 |  |  | 33 |  |
| N |  | 1883 |  |  | 2009 |  |

Note: Participants reporting Native Hawaiian/Pacific Islander, Middle east/North Africa or Other as ethnicity are the reference group for the ethnicity variables. Personal Income < \$10.000 is the reference group for the "Personal Income" variables. Educational level < high school diploma is the reference group for the "Educational Level" variables. The models are computed with robust covariance matrix estimation.

**Table S17.** Logistic regression predicting vaping (not vaping=0, vaping=1) (with sex as covariate).

|  | *SAMPLE 1* |  |  | *SAMPLE 2* |  |  |
| --- | --- | --- | --- | --- | --- | --- |
|  | Incidence ratio | rate | 95% CI | Incidence ratio | rate | 95% CI |
| Caregiver strain |  | 1.06 | 0.90 1.26 |  | 1.10 | 0.93 1.29 |
| Work strain |  | 1.23 | 0.97 1.55 |  | 1.29 | 1.05 1.59 |
| Independence |  | 1.17 | 0.95 1.45 |  | 1.05 | 0.76 1.44 |
| Risk-taking |  | 1.20 | 0.98 1.48 |  | 1.00 | 0.80 1.25 |
| Emotional intelligence |  | 1.39 | 1.12 1.71 |  | 1.05 | 0.84 1.31 |
| Social support |  | 0.98 | 0.84 1.14 |  | 0.88 | 0.76 1.04 |
| Discrimination |  | 1.40 | 1.15 1.70 |  | 1.46 | 1.17 1.81 |
| Year of birth |  | 0.98 | 0.97 1.00 |  | 1.01 | 1.00 1.03 |
| Personal Income (\$10,000-\$19,999) | | 1.97 | 0.98 3.95 | | 1.20 | 0.60 2.38 |
| Personal Income (\$20,000-\$29,999) | | 1.70 | 0.85 3.40 | | 1.65 | 0.88 3.12 |
| Personal Income (\$30,000-\$39,999) | | 2.65 | 1.36 5.14 | | 1.04 | 0.52 2.09 |
| Personal Income (\$40,000-\$49,999) | | 2.11 | 1.01 4.39 | | 1.09 | 0.53 2.23 |
| Personal Income (\$50,000-\$59,999) | | 2.49 | 1.23 5.04 | | 1.48 | 0.72 3.05 |
| Personal Income (\$60,000-\$69,999) | | 2.99 | 1.42 6.29 | | 1.32 | 0.58 3.01 |
| Personal Income (\$70,000-\$79,999) | | 1.72 | 0.74 3.98 | | 0.90 | 0.35 2.33 |
| Personal Income (\$80,000-\$89,999) | | 0.72 | 0.16 3.22 | | 1.03 | 0.32 3.33 |
| Personal Income (\$90,000-\$99,999) | | 4.89 | 1.91 12.55 | | 1.02 | 0.33 3.14 |
| Personal Income (\$100,000-\$149,999) | | 2.26 | 0.98 5.21 | | 0.91 | 0.35 2.32 |
| Personal Income (More than \$150,000) | | 7.17 | 3.21 16.00 | | 0.46 | 0.06 3.77 |
| Educational Level (High school graduate) |  | 0.35 | 0.13 0.93 |  | 0.85 | 0.16 4.59 |
| Educational Level (Some college credit, no degree) |  | 0.29 | 0.11 0.76 |  | 0.81 | 0.16 4.16 |
| Educational Level (Trade/Technical/Vocational education) |  | 0.37 | 0.10 1.32 |  | 0.45 | 0.07 2.88 |
| Educational Level (Associate degree) |  | 0.28 | 0.10 0.78 |  | 1.17 | 0.23 6.04 |
| Educational Level (Bachelor's degree) |  | 0.17 | 0.07 0.44 |  | 0.66 | 0.13 3.35 |
| Educational Level (Master's degree) |  | 0.25 | 0.09 0.67 |  | 0.45 | 0.08 2.44 |
| Educational Level (professional degree) |  | 0.29 | 0.06 1.39 |  | 0.50 | 0.06 4.46 |
| Educational Level (Doctorate degree) |  | 0.21 | 0.06 0.68 |  | 0.49 | 0.06 4.39 |
| Ethnicity (White) |  | 1.85 | 1.02 3.36 |  | 1.05 | 0.53 2.05 |
| Ethnicity (Hispanic, Latinx, Spanish) |  | 1.23 | 0.64 2.37 |  | 1.36 | 0.63 2.92 |
| Ethnicity (Black, African American) |  | 1.44 | 0.67 3.08 |  | 0.73 | 0.35 1.53 |
| Ethnicity (Asian) |  | 1.18 | 0.58 2.39 |  | 0.73 | 0.29 1.87 |
| Ethnicity (Native) |  | 1.48 | 0.60 3.61 |  | 1.63 | 0.67 3.95 |
| Sex (male=0, female=1) |  | 0.45 | 0.31 0.64 |  | 0.76 | 0.52 1.11 |
| **Goodness of Fit** |  |  |  |  |  |  |
| Deviance/df |  | 0.643 |  |  | 0.572 |  |
| Pearson Chi-Square/df |  | 1.040 |  |  | 1.013 |  |
| Loglikelihood |  | -594.193 |  |  | -564.518 |  |
| AIC |  | 1256.386 |  |  | 1197.036 |  |
| **Omnibus tests** |  |  |  |  |  |  |
| Chi-square |  | 169.275 |  |  | 59.470 |  |
| df |  | 33 |  |  | 33 |  |
| N |  | 1882 |  |  | 2009 |  |

Note: Participants reporting Native Hawaiian/Pacific Islander, Middle east/North Africa or Other as ethnicity are the reference group for the ethnicity variables. Personal Income < \$10.000 is the reference group for the "Personal Income" variables. Educational level < high school diploma is the reference group for the "Educational Level" variables. The models are computed with robust covariance matrix estimation.

**Table S18.** Logistic regression predicting binge drinking (less than monthly=0, monthly, weekly, and daily=1) (with sex as covariate).

|  | *SAMPLE 1* |  |  | *SAMPLE 2* |  |  |
| --- | --- | --- | --- | --- | --- | --- |
|  | Incidence ratio | rate | 95% CI | Incidence ratio | rate | 95% CI |
| Caregiver strain |  | 0.92 | 0.81 1.06 |  | 0.91 | 0.80 1.04 |
| Work strain |  | 1.28 | 1.08 1.51 |  | 1.03 | 0.88 1.21 |
| Independence |  | 1.08 | 0.93 1.25 |  | 1.19 | 0.94 1.49 |
| Risk-taking |  | 1.53 | 1.31 1.78 |  | 1.25 | 1.07 1.45 |
| Emotional intelligence |  | 1.18 | 1.00 1.40 |  | 1.04 | 0.88 1.21 |
| Social support |  | 1.01 | 0.90 1.13 |  | 0.87 | 0.78 0.97 |
| Discrimination |  | 1.18 | 1.00 1.39 |  | 1.11 | 0.92 1.32 |
| Year of birth |  | 1.02 | 1.01 1.03 |  | 1.03 | 1.02 1.04 |
| Personal Income (\$10,000-\$19,999) | | 1.02 | 0.64 1.60 | | 1.26 | 0.78 2.03 |
| Personal Income (\$20,000-\$29,999) | | 1.14 | 0.73 1.81 | | 1.99 | 1.26 3.13 |
| Personal Income (\$30,000-\$39,999) | | 1.18 | 0.75 1.87 | | 1.34 | 0.82 2.20 |
| Personal Income (\$40,000-\$49,999) | | 1.29 | 0.79 2.09 | | 1.81 | 1.09 2.99 |
| Personal Income (\$50,000-\$59,999) | | 0.91 | 0.55 1.50 | | 1.39 | 0.81 2.38 |
| Personal Income (\$60,000-\$69,999) | | 1.19 | 0.70 2.03 | | 1.98 | 1.11 3.55 |
| Personal Income (\$70,000-\$79,999) | | 0.81 | 0.44 1.51 | | 1.44 | 0.78 2.65 |
| Personal Income (\$80,000-\$89,999) | | 1.15 | 0.52 2.55 | | 1.16 | 0.52 2.59 |
| Personal Income (\$90,000-\$99,999) | | 1.66 | 0.72 3.81 | | 2.31 | 1.14 4.67 |
| Personal Income (\$100,000-\$149,999) | | 1.13 | 0.62 2.08 | | 1.95 | 1.05 3.59 |
| Personal Income (More than \$150,000) | | 2.01 | 1.01 4.03 | | 2.44 | 1.01 5.92 |
| Educational Level (High school graduate) |  | 1.22 | 0.41 3.65 |  | 0.50 | 0.15 1.66 |
| Educational Level (Some college credit, no degree) |  | 1.68 | 0.58 4.82 |  | 0.90 | 0.29 2.81 |
| Educational Level (Trade/Technical/Vocational education) |  | 2.44 | 0.73 8.16 |  | 0.85 | 0.25 2.93 |
| Educational Level (Associate degree) |  | 1.37 | 0.45 4.18 |  | 0.65 | 0.20 2.11 |
| Educational Level (Bachelor's degree) |  | 1.54 | 0.54 4.41 |  | 0.63 | 0.20 1.96 |
| Educational Level (Master's degree) |  | 1.82 | 0.62 5.34 |  | 0.58 | 0.18 1.89 |
| Educational Level (professional degree) |  | 3.28 | 0.78 13.78 |  | 0.58 | 0.13 2.50 |
| Educational Level (Doctorate degree) |  | 1.77 | 0.52 5.95 |  | 0.65 | 0.15 2.74 |
| Ethnicity (White) |  | 1.89 | 1.08 3.31 |  | 0.93 | 0.54 1.60 |
| Ethnicity (Hispanic, Latinx, Spanish) |  | 1.63 | 0.96 2.77 |  | 0.86 | 0.50 1.51 |
| Ethnicity (Black, African American) |  | 0.74 | 0.36 1.53 |  | 1.05 | 0.57 1.93 |
| Ethnicity (Asian) |  | 0.59 | 0.30 1.13 |  | 0.54 | 0.27 1.09 |
| Ethnicity (Native) |  | 0.45 | 0.19 1.06 |  | 0.79 | 0.38 1.65 |
| Sex (male=0, female=1) |  | 0.55 | 0.42 0.71 |  | 0.77 | 0.59 1.01 |
| **Goodness of Fit** |  |  |  |  |  |  |
| Deviance/df |  | 1.000 |  |  | 1.002 |  |
| Pearson Chi-Square/df |  | 1.017 |  |  | 1.011 |  |
| Loglikelihood |  | -922.640 |  |  | -988.206 |  |
| AIC |  | 1913.280 |  |  | 2044.412 |  |
| **Omnibus tests** |  |  |  |  |  |  |
| Chi-square |  | 214.714 |  |  | 116.766 |  |
| df |  | 33 |  |  | 33 |  |
| N |  | 1880 |  |  | 2009 |  |

Note: Participants reporting Native Hawaiian/Pacific Islander, Middle east/North Africa or Other as ethnicity are the reference group for the ethnicity variables. Personal Income < \$10.000 is the reference group for the "Personal Income" variables. Educational level < high school diploma is the reference group for the "Educational Level" variables. The models are computed with robust covariance matrix estimation.

**Table S19.** Logistic regression predicting Overweight (BMI<25=0, BMI≥25 =1) (with sex as covariate).

|  | *SAMPLE 1* |  |  | *SAMPLE 2* |  |  |
| --- | --- | --- | --- | --- | --- | --- |
|  | Incidence ratio | rate | 95% CI | Incidence ratio | rate | 95% CI |
| Caregiver strain |  | 1.18 | 1.05 1.32 |  | 1.27 | 1.13 1.41 |
| Work strain |  | 1.04 | 0.91 1.19 |  | 1.17 | 1.02 1.33 |
| Independence |  | 0.99 | 0.88 1.12 |  | 1.05 | 0.87 1.26 |
| Risk-taking |  | 0.82 | 0.72 0.93 |  | 0.85 | 0.75 0.97 |
| Emotional intelligence |  | 0.99 | 0.86 1.14 |  | 1.03 | 0.90 1.18 |
| Social support |  | 1.00 | 0.91 1.10 |  | 1.07 | 0.97 1.18 |
| Discrimination |  | 1.23 | 1.06 1.42 |  | 1.26 | 1.08 1.48 |
| Year of birth |  | 0.98 | 0.97 0.99 |  | 0.97 | 0.96 0.98 |
| Personal Income (\$10,000-\$19,999) | | 1.12 | 0.79 1.58 | | 0.83 | 0.57 1.19 |
| Personal Income (\$20,000-\$29,999) | | 1.20 | 0.84 1.72 | | 0.84 | 0.58 1.21 |
| Personal Income (\$30,000-\$39,999) | | 1.13 | 0.78 1.63 | | 0.77 | 0.52 1.13 |
| Personal Income (\$40,000-\$49,999) | | 1.15 | 0.78 1.69 | | 0.77 | 0.52 1.15 |
| Personal Income (\$50,000-\$59,999) | | 1.46 | 0.97 2.19 | | 1.16 | 0.75 1.79 |
| Personal Income (\$60,000-\$69,999) | | 1.29 | 0.82 2.02 | | 1.28 | 0.76 2.16 |
| Personal Income (\$70,000-\$79,999) | | 1.27 | 0.75 2.14 | | 2.21 | 1.31 3.74 |
| Personal Income (\$80,000-\$89,999) | | 1.16 | 0.59 2.31 | | 0.99 | 0.54 1.79 |
| Personal Income (\$90,000-\$99,999) | | 1.91 | 0.84 4.32 | | 1.07 | 0.58 1.97 |
| Personal Income (\$100,000-\$149,999) | | 1.02 | 0.60 1.74 | | 1.01 | 0.60 1.70 |
| Personal Income (More than \$150,000) | | 1.27 | 0.65 2.48 | | 0.97 | 0.44 2.17 |
| Educational Level (High school graduate) |  | 0.86 | 0.39 1.90 |  | 0.24 | 0.07 0.84 |
| Educational Level (Some college credit, no degree) |  | 0.90 | 0.42 1.92 |  | 0.32 | 0.09 1.09 |
| Educational Level (Trade/Technical/Vocational education) |  | 0.88 | 0.34 2.28 |  | 0.21 | 0.06 0.76 |
| Educational Level (Associate degree) |  | 1.06 | 0.47 2.40 |  | 0.23 | 0.07 0.80 |
| Educational Level (Bachelor's degree) |  | 0.59 | 0.28 1.26 |  | 0.21 | 0.06 0.72 |
| Educational Level (Master's degree) |  | 0.78 | 0.35 1.73 |  | 0.21 | 0.06 0.74 |
| Educational Level (professional degree) |  | 0.58 | 0.18 1.82 |  | 0.13 | 0.03 0.52 |
| Educational Level (Doctorate degree) |  | 0.69 | 0.27 1.79 |  | 0.13 | 0.03 0.51 |
| Ethnicity (White) |  | 1.08 | 0.70 1.66 |  | 1.03 | 0.64 1.67 |
| Ethnicity (Hispanic, Latinx, Spanish) |  | 1.28 | 0.81 2.03 |  | 1.60 | 0.97 2.63 |
| Ethnicity (Black, African American) |  | 1.66 | 0.99 2.76 |  | 1.16 | 0.67 1.99 |
| Ethnicity (Asian) |  | 0.49 | 0.30 0.80 |  | 0.54 | 0.30 0.95 |
| Ethnicity (Native) |  | 2.10 | 1.02 4.34 |  | 1.17 | 0.61 2.23 |
| Sex (male=0, female=1) |  | 0.76 | 0.60 0.95 |  | 0.58 | 0.46 0.73 |
| **Goodness of Fit** |  |  |  |  |  |  |
| Deviance/df |  | 1.313 |  |  | 1.283 |  |
| Pearson Chi-Square/df |  | 1.020 |  |  | 1.020 |  |
| Loglikelihood |  | -1197.707 |  |  | -1256.644 |  |
| AIC |  | 2463.413 |  |  | 2581.288 |  |
| **Omnibus tests** |  |  |  |  |  |  |
| Chi-square |  | 137.355 |  |  | 163.390 |  |
| df |  | 33 |  |  | 33 |  |
| N |  | 1858 |  |  | 1993 |  |

Note: Participants reporting Native Hawaiian/Pacific Islander, Middle east/North Africa or Other as ethnicity are the reference group for the ethnicity variables. Personal Income < \$10.000 is the reference group for the "Personal Income" variables. Educational level < high school diploma is the reference group for the "Educational Level" variables. The models are computed with robust covariance matrix estimation.

**Table S20.** Negative binomial regression predicting number of days with poor physical health (during past 30 days) (combined samples).

|  | *SEX* |  |  | *GENDER* |  |  |
| --- | --- | --- | --- | --- | --- | --- |
|  | Incidence ratio | rate | 99.9% CI | Incidence ratio | rate | 99.9% CI |
| Caregiver strain |  | 1.15 | 1.06 1.25 |  | 1.152 | 1.063 1.248 |
| Work strain |  | 0.98 | 0.88 1.08 |  | 0.978 | 0.883 1.084 |
| Independence |  | 0.90 | 0.80 1.02 |  | 0.902 | 0.801 1.016 |
| Risk-taking |  | 0.98 | 0.89 1.09 |  | 0.979 | 0.884 1.083 |
| Emotional intelligence |  | 0.94 | 0.84 1.05 |  | 0.938 | 0.839 1.049 |
| Social support |  | 0.95 | 0.87 1.03 |  | 0.946 | 0.874 1.025 |
| Discrimination |  | 1.45 | 1.30 1.61 |  | 1.446 | 1.305 1.603 |
| Year of birth |  | 0.99 | 0.98 0.99 |  | 0.988 | 0.981 0.994 |
| Personal Income (\$10,000-\$19,999) | | 1.06 | 0.80 1.40 | | 1.063 | 0.804 1.406 |
| Personal Income (\$20,000-\$29,999) | | 0.99 | 0.75 1.30 | | 0.986 | 0.748 1.300 |
| Personal Income (\$30,000-\$39,999) | | 0.87 | 0.63 1.19 | | 0.868 | 0.632 1.191 |
| Personal Income (\$40,000-\$49,999) | | 0.85 | 0.62 1.17 | | 0.852 | 0.619 1.174 |
| Personal Income (\$50,000-\$59,999) | | 0.79 | 0.54 1.15 | | 0.794 | 0.545 1.156 |
| Personal Income (\$60,000-\$69,999) | | 0.76 | 0.52 1.12 | | 0.766 | 0.524 1.118 |
| Personal Income (\$70,000-\$79,999) | | 0.70 | 0.45 1.09 | | 0.701 | 0.447 1.098 |
| Personal Income (\$80,000-\$89,999) | | 0.54 | 0.33 0.89 | | 0.538 | 0.327 0.886 |
| Personal Income (\$90,000-\$99,999) | | 0.89 | 0.49 1.60 | | 0.922 | 0.516 1.647 |
| Personal Income (\$100,000-\$149,999) | | 0.72 | 0.46 1.13 | | 0.721 | 0.458 1.134 |
| Personal Income (More than \$150,000) | | 0.99 | 0.56 1.75 | | 1.055 | 0.598 1.859 |
| Educational Level (High school graduate) |  | 0.95 | 0.51 1.77 |  | 0.951 | 0.510 1.772 |
| Educational Level (Some college credit, no degree) |  | 0.96 | 0.53 1.75 |  | 0.970 | 0.535 1.759 |
| Educational Level (Trade/Technical/Vocational education) |  | 0.84 | 0.43 1.67 |  | 0.840 | 0.425 1.662 |
| Educational Level (Associate degree) |  | 0.88 | 0.48 1.63 |  | 0.877 | 0.474 1.622 |
| Educational Level (Bachelor's degree) |  | 0.78 | 0.43 1.41 |  | 0.779 | 0.431 1.408 |
| Educational Level (Master's degree) |  | 0.86 | 0.46 1.61 |  | 0.860 | 0.459 1.612 |
| Educational Level (professional degree) |  | 0.80 | 0.30 2.14 |  | 0.792 | 0.295 2.122 |
| Educational Level (Doctorate degree) |  | 0.70 | 0.33 1.48 |  | 0.695 | 0.328 1.471 |
| Ethnicity (White) |  | 0.98 | 0.70 1.35 |  | 0.950 | 0.685 1.318 |
| Ethnicity (Hispanic, Latinx, Spanish) |  | 0.97 | 0.66 1.42 |  | 0.953 | 0.652 1.393 |
| Ethnicity (Black, African American) |  | 0.89 | 0.62 1.27 |  | 0.873 | 0.610 1.248 |
| Ethnicity (Asian) |  | 0.83 | 0.55 1.24 |  | 0.808 | 0.538 1.214 |
| Ethnicity (Native) |  | 1.24 | 0.79 1.94 |  | 1.236 | 0.790 1.935 |
| Sex (male=0, female=1) |  | 0.92 | 0.77 1.11 |  |  |  |
| Woman |  |  |  |  | 0.926 | 0.773 1.109 |
| Non-binary/Gender fluid + Gender=other |  |  |  |  | 0.844 | 0.381 1.871 |
| Sample (Sample 1=1) |  | 0.82 | 0.65 1.02 |  | 0.816 | 0.652 1.022 |
| **Goodness of Fit** |  |  |  |  |  |  |
| Deviance/df |  | 1.107 |  |  | 1.107 |  |
| Pearson Chi-Square/df |  | 1.198 |  |  | 1.197 |  |
| Loglikelihood |  | -10001.663 |  |  | -10032.722 |  |
| AIC |  | 20075.326 |  |  | 20139.445 |  |
| **Omnibus tests** |  |  |  |  |  |  |
| Chi-square |  | 340.684 |  |  | 345.482 |  |
| df |  | 34 |  |  | 35 |  |
| N |  | 3879 |  |  | 3889 |  |

Note: Participants reporting Native Hawaiian/Pacific Islander, Middle east/North Africa or Other as ethnicity are the reference group for the ethnicity variables. Personal Income < \$10,000 is the reference group for the "Personal Income" variables. Educational level < high school diploma is the reference group for the "Educational Level" variables. The models are computed with robust covariance matrix estimation.

**Table S21.** Negative binomial regression predicting number of days with mental health (during past 30 days) (combined samples).

|  | *SEX* |  |  | *GENDER* |  |  |
| --- | --- | --- | --- | --- | --- | --- |
|  | Incidence ratio | rate | 99.9% CI | Incidence ratio | rate | 99.9% CI |
| Caregiver strain |  | 1.20 | 1.12 1.28 |  | 1.20 | 1.13 1.28 |
| Work strain |  | 1.07 | 0.98 1.17 |  | 1.07 | 0.98 1.17 |
| Independence |  | 0.93 | 0.84 1.02 |  | 0.93 | 0.84 1.02 |
| Risk-taking |  | 0.92 | 0.84 1.01 |  | 0.92 | 0.84 1.01 |
| Emotional intelligence |  | 0.91 | 0.83 1.01 |  | 0.91 | 0.83 1.01 |
| Social support |  | 0.83 | 0.78 0.89 |  | 0.84 | 0.78 0.90 |
| Discrimination |  | 1.40 | 1.28 1.54 |  | 1.40 | 1.28 1.53 |
| Year of birth |  | 1.02 | 1.01 1.02 |  | 1.02 | 1.01 1.02 |
| Personal Income (\$10,000-\$19,999) | | 0.99 | 0.79 1.23 | | 0.99 | 0.79 1.23 |
| Personal Income (\$20,000-\$29,999) | | 0.83 | 0.65 1.06 | | 0.84 | 0.66 1.07 |
| Personal Income (\$30,000-\$39,999) | | 0.81 | 0.63 1.04 | | 0.81 | 0.63 1.04 |
| Personal Income (\$40,000-\$49,999) | | 0.69 | 0.53 0.90 | | 0.69 | 0.53 0.90 |
| Personal Income (\$50,000-\$59,999) | | 0.66 | 0.49 0.90 | | 0.66 | 0.49 0.90 |
| Personal Income (\$60,000-\$69,999) | | 0.65 | 0.47 0.90 | | 0.65 | 0.47 0.90 |
| Personal Income (\$70,000-\$79,999) | | 0.61 | 0.41 0.90 | | 0.61 | 0.41 0.90 |
| Personal Income (\$80,000-\$89,999) | | 0.76 | 0.44 1.30 | | 0.75 | 0.44 1.28 |
| Personal Income (\$90,000-\$99,999) | | 0.78 | 0.46 1.34 | | 0.77 | 0.45 1.32 |
| Personal Income (\$100,000-\$149,999) | | 0.63 | 0.41 0.96 | | 0.63 | 0.41 0.95 |
| Personal Income (More than \$150,000) | | 0.76 | 0.44 1.30 | | 0.79 | 0.46 1.36 |
| Educational Level (High school graduate) |  | 1.14 | 0.65 2.01 |  | 1.14 | 0.65 2.00 |
| Educational Level (Some college credit, no degree) |  | 1.18 | 0.69 2.04 |  | 1.18 | 0.68 2.03 |
| Educational Level (Trade/Technical/Vocational education) |  | 1.32 | 0.69 2.53 |  | 1.32 | 0.69 2.53 |
| Educational Level (Associate degree) |  | 1.13 | 0.64 1.99 |  | 1.12 | 0.64 1.98 |
| Educational Level (Bachelor's degree) |  | 1.02 | 0.60 1.75 |  | 1.02 | 0.59 1.75 |
| Educational Level (Master's degree) |  | 1.17 | 0.66 2.07 |  | 1.17 | 0.66 2.07 |
| Educational Level (professional degree) |  | 1.05 | 0.46 2.43 |  | 1.04 | 0.45 2.40 |
| Educational Level (Doctorate degree) |  | 1.15 | 0.58 2.29 |  | 1.14 | 0.57 2.27 |
| Ethnicity (White) |  | 1.37 | 1.03 1.83 |  | 1.34 | 1.00 1.79 |
| Ethnicity (Hispanic, Latinx, Spanish) |  | 1.07 | 0.80 1.43 |  | 1.06 | 0.79 1.42 |
| Ethnicity (Black, African American) |  | 1.07 | 0.78 1.45 |  | 1.05 | 0.77 1.43 |
| Ethnicity (Asian) |  | 1.04 | 0.73 1.48 |  | 1.02 | 0.72 1.46 |
| Ethnicity (Native) |  | 1.47 | 0.93 2.30 |  | 1.46 | 0.93 2.29 |
| Sex (male=0, female=1) |  | 1.05 | 0.89 1.23 |  |  |  |
| Woman |  |  |  |  | 1.05 | 0.89 1.23 |
| Non-binary/Gender fluid + Gender=other |  |  |  |  | 1.23 | 0.78 1.93 |
| Sample (Sample 1=1) |  | 0.90 | 0.75 1.09 |  | 0.91 | 0.75 1.09 |
| **Goodness of Fit** |  |  |  |  |  |  |
| Deviance/df |  | 1.145 |  |  | 1.146 |  |
| Pearson Chi-Square/df |  | 1.059 |  |  | 1.058 |  |
| Loglikelihood |  | -11254.537 |  |  | -11285.196 |  |
| AIC |  | 22581.073 |  |  | 22644.392 |  |
| **Omnibus tests** |  |  |  |  |  |  |
| Chi-square |  | 567.157 |  |  | 568.564 |  |
| df |  | 34 |  |  | 35 |  |
| N |  | 3880 |  |  | 3890 |  |

**Table S22.**
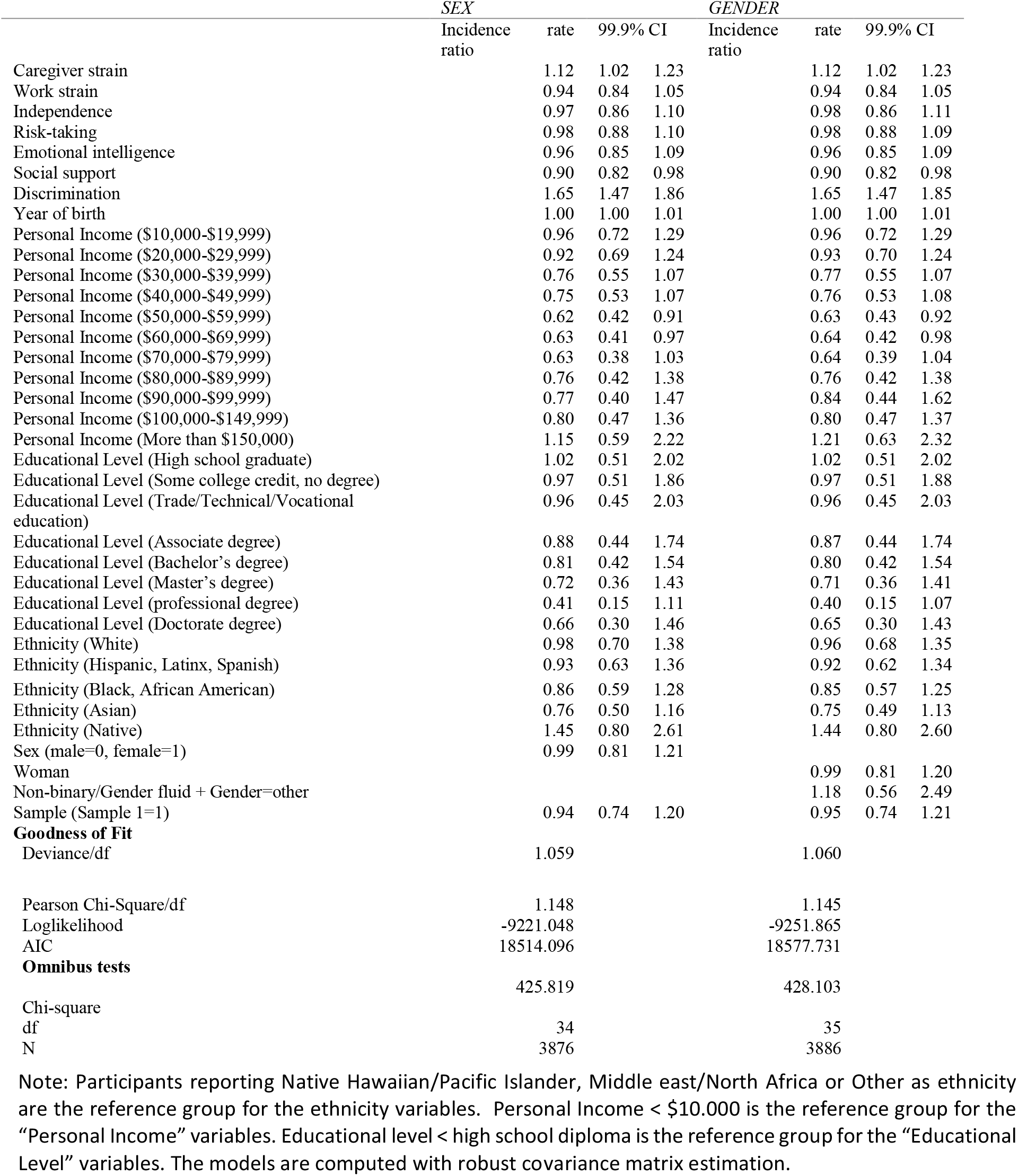
Negative binomial regression predicting number of days where poor mental or physical health prevented the respondent from doing usual activities (during past 30 days) (combined samples).

**Table S23.** Logistic regression predicting general health status (excellent, very good, good= 0, fair, poor= 1) (combined samples).

|  | *SEX* |  |  | *GENDER* |  |  |
| --- | --- | --- | --- | --- | --- | --- |
|  | Odds ratio | 99.9% CI |  | Odds ratio | 99.9% CI |  |
| Caregiver strain | 1.21 | 1.04 | 1.42 | 1.21 | 1.03 | 1.42 |
| Work strain | 0.93 | 0.76 | 1.15 | 0.93 | 0.76 | 1.14 |
| Independence | 0.83 | 0.66 | 1.03 | 0.83 | 0.66 | 1.03 |
| Risk-taking | 0.63 | 0.52 | 0.77 | 0.62 | 0.51 | 0.76 |
| Emotional intelligence | 0.89 | 0.72 | 1.10 | 0.89 | 0.72 | 1.11 |
| Social support | 0.81 | 0.70 | 0.94 | 0.81 | 0.70 | 0.94 |
| Discrimination | 1.48 | 1.19 | 1.85 | 1.49 | 1.20 | 1.86 |
| Year of birth | 0.98 | 0.97 | 0.99 | 0.98 | 0.97 | 0.99 |
| Personal Income (\$10,000-\$19,999) | 0.95 | 0.59 | 1.54 | 0.95 | 0.59 | 1.54 |
| Personal Income (\$20,000-\$29,999) | 0.81 | 0.49 | 1.35 | 0.81 | 0.49 | 1.35 |
| Personal Income (\$30,000-\$39,999) | 0.71 | 0.40 | 1.25 | 0.71 | 0.41 | 1.25 |
| Personal Income (\$40,000-\$49,999) | 0.67 | 0.37 | 1.23 | 0.67 | 0.37 | 1.23 |
| Personal Income (\$50,000-\$59,999) | 0.57 | 0.29 | 1.09 | 0.57 | 0.29 | 1.09 |
| Personal Income (\$60,000-\$69,999) | 0.79 | 0.38 | 1.62 | 0.79 | 0.39 | 1.63 |
| Personal Income (\$70,000-\$79,999) | 0.61 | 0.27 | 1.38 | 0.61 | 0.27 | 1.38 |
| Personal Income (\$80,000-\$89,999) | 0.35 | 0.10 | 1.29 | 0.35 | 0.10 | 1.29 |
| Personal Income (\$90,000-\$99,999) | 0.66 | 0.20 | 2.12 | 0.69 | 0.22 | 2.12 |
| Personal Income (\$100,000-\$149,999) | 0.37 | 0.14 | 0.97 | 0.37 | 0.14 | 0.98 |
| Personal Income (More than \$150,000) | 0.30 | 0.06 | 1.58 | 0.30 | 0.06 | 1.59 |
| Educational Level (High school graduate) | 0.59 | 0.19 | 1.84 | 0.59 | 0.19 | 1.85 |
| Educational Level (Some college credit, no degree) | 0.71 | 0.24 | 2.10 | 0.71 | 0.24 | 2.10 |
| Educational Level (Trade/Technical/Vocational education) | 0.65 | 0.18 | 2.31 | 0.65 | 0.18 | 2.31 |
| Educational Level (Associate degree) | 0.68 | 0.22 | 2.10 | 0.68 | 0.22 | 2.10 |
| Educational Level (Bachelor's degree) | 0.43 | 0.15 | 1.28 | 0.44 | 0.15 | 1.29 |
| Educational Level (Master's degree) | 0.40 | 0.13 | 1.28 | 0.40 | 0.12 | 1.28 |
| Educational Level (professional degree) | 0.27 | 0.04 | 1.87 | 0.27 | 0.04 | 1.87 |
| Educational Level (Doctorate degree) | 0.33 | 0.07 | 1.49 | 0.32 | 0.07 | 1.49 |
| Ethnicity (White) | 1.31 | 0.64 | 2.67 | 1.31 | 0.64 | 2.67 |
| Ethnicity (Hispanic, Latinx, Spanish) | 0.95 | 0.44 | 2.04 | 0.95 | 0.44 | 2.05 |
| Ethnicity (Black, African American) | 1.30 | 0.61 | 2.79 | 1.33 | 0.62 | 2.84 |
| Ethnicity (Asian) | 1.14 | 0.48 | 2.67 | 1.13 | 0.48 | 2.65 |
| Ethnicity (Native) | 0.69 | 0.26 | 1.86 | 0.69 | 0.26 | 1.86 |
| Sex (male=0, female=1) | 0.64 | 0.45 | 0.91 |  |  |  |
| Woman |  |  |  | 0.62 | 0.44 | 0.88 |
| Non-binary/Gender fluid + Gender=other |  |  |  | 0.84 | 0.19 | 3.72 |
| Sample (Sample 1=1) | 0.79 | 0.52 | 1.18 | 0.79 | 0.52 | 1.18 |
| **Goodness of Fit** |  |  |  |  |  |  |
| Deviance/df | 0.857 |  |  | 0.857 |  |  |
| Pearson Chi-Square/df | 1.006 |  |  | 1.010 |  |  |
| Loglikelihood | -1656.691 |  |  | -1653.960 |  |  |
| AIC | 3385.382 |  |  | 3377.919 |  |  |
| **Omnibus tests** |  |  |  | 351.378 |  |  |
|  | 358.997 |  |  | 351.378 |  |  |
| Chi-square |  |  |  |  |  |  |
| df |  |  |  | 34 |  |  |
| N | 3894 |  |  | 3904 |  |  |

Note: Participants reporting Native Hawaiian/Pacific Islander, Middle east/North Africa or Other as ethnicity are the reference group for the ethnicity variables. Personal Income < \$10,000 is the reference group for the "Personal Income" variables. Educational level < high school diploma is the reference group for the "Educational Level" variables. The models are computed with robust covariance matrix estimation.

**Table S24.** Logistic regression predicting smoking (not smoking=0, smoking=1) (combined samples).

|  | *SEX* |  |  | *GENDER* |  |  |
| --- | --- | --- | --- | --- | --- | --- |
|  | Odds ratio | 99.9% CI |  | Odds ratio | 99.9% CI |  |
| Caregiver strain | 1.29 | 1.12 | 1.50 | 1.29 | 1.12 | 1.50 |
| Work strain | 1.31 | 1.08 | 1.58 | 1.30 | 1.08 | 1.57 |
| Independence | 1.19 | 0.95 | 1.48 | 1.19 | 0.95 | 1.49 |
| Risk-taking | 1.20 | 1.00 | 1.44 | 1.21 | 1.01 | 1.45 |
| Emotional intelligence | 1.28 | 1.06 | 1.55 | 1.28 | 1.06 | 1.55 |
| Social support | 0.98 | 0.85 | 1.12 | 0.97 | 0.85 | 1.12 |
| Discrimination | 1.16 | 0.94 | 1.43 | 1.17 | 0.95 | 1.44 |
| Year of birth | 0.99 | 0.98 | 1.00 | 0.99 | 0.98 | 1.00 |
| Personal Income (\$10,000-\$19,999) | 1.21 | 0.71 | 2.05 | 1.22 | 0.72 | 2.08 |
| Personal Income (\$20,000-\$29,999) | 1.13 | 0.67 | 1.93 | 1.13 | 0.66 | 1.93 |
| Personal Income (\$30,000-\$39,999) | 1.08 | 0.61 | 1.92 | 1.10 | 0.62 | 1.94 |
| Personal Income (\$40,000-\$49,999) | 1.04 | 0.57 | 1.88 | 1.04 | 0.57 | 1.88 |
| Personal Income (\$50,000-\$59,999) | 1.01 | 0.54 | 1.88 | 1.01 | 0.54 | 1.89 |
| Personal Income (\$60,000-\$69,999) | 1.08 | 0.53 | 2.17 | 1.08 | 0.54 | 2.18 |
| Personal Income (\$70,000-\$79,999) | 1.09 | 0.52 | 2.28 | 1.10 | 0.53 | 2.29 |
| Personal Income (\$80,000-\$89,999) | 1.05 | 0.40 | 2.73 | 1.06 | 0.41 | 2.75 |
| Personal Income (\$90,000-\$99,999) | 1.00 | 0.37 | 2.71 | 0.98 | 0.36 | 2.65 |
| Personal Income (\$100,000-\$149,999) | 0.99 | 0.47 | 2.09 | 0.99 | 0.47 | 2.10 |
| Personal Income (More than \$150,000) | 2.53 | 1.07 | 6.01 | 2.46 | 1.03 | 5.84 |
| Educational Level (High school graduate) | 1.00 | 0.29 | 3.42 | 1.01 | 0.29 | 3.44 |
| Educational Level (Some college credit, no degree) | 0.87 | 0.26 | 2.89 | 0.88 | 0.26 | 2.92 |
| Educational Level (Trade/Technical/Vocational education) | 1.37 | 0.36 | 5.15 | 1.37 | 0.36 | 5.14 |
| Educational Level (Associate degree) | 0.82 | 0.24 | 2.83 | 0.82 | 0.24 | 2.82 |
| Educational Level (Bachelor's degree) | 0.40 | 0.12 | 1.33 | 0.40 | 0.12 | 1.34 |
| Educational Level (Master's degree) | 0.39 | 0.11 | 1.34 | 0.39 | 0.11 | 1.35 |
| Educational Level (professional degree) | 0.41 | 0.07 | 2.29 | 0.42 | 0.08 | 2.31 |
| Educational Level (Doctorate degree) | 0.52 | 0.12 | 2.20 | 0.53 | 0.12 | 2.24 |
| Ethnicity (White) | 0.89 | 0.44 | 1.83 | 0.90 | 0.44 | 1.85 |
| Ethnicity (Hispanic, Latinx, Spanish) | 0.62 | 0.29 | 1.31 | 0.62 | 0.30 | 1.31 |
| Ethnicity (Black, African American) | 0.51 | 0.22 | 1.18 | 0.51 | 0.22 | 1.19 |
| Ethnicity (Asian) | 0.31 | 0.11 | 0.84 | 0.31 | 0.11 | 0.83 |
| Ethnicity (Native) | 1.24 | 0.54 | 2.89 | 1.27 | 0.54 | 2.97 |
| Sex (male=0, female=1) | 0.77 | 0.56 | 1.07 |  |  |  |
| Woman |  |  |  | 0.78 | 0.56 | 1.07 |
| Non-binary/Gender fluid + Gender=other |  |  |  | 0.19 | 0.02 | 2.07 |
| Sample (Sample 1=1) | 1.44 | 0.94 | 2.20 | 1.45 | 0.95 | 2.22 |
| **Goodness of Fit** |  |  |  |  |  |  |
| Deviance/df | 0.936 |  |  | 0.934 |  |  |
| Pearson Chi-Square/df | 0.986 |  |  | 0.983 |  |  |
| Loglikelihood | -1804.942 |  |  | -1804.685 |  |  |
| AIC | 3679.885 |  |  | 3681.370 |  |  |
| **Omnibus tests** |  |  |  |  |  |  |
|  | 322.659 |  |  | 329.998 |  |  |
| Chi-square |  |  |  |  |  |  |
| df | 34 |  |  | 35 |  |  |
| N | 3892 |  |  | 3901 |  |  |

Note: Participants reporting Native Hawaiian/Pacific Islander, Middle east/North Africa or Other as ethnicity are the reference group for the ethnicity variables. Personal Income < \$10,000 is the reference group for the "Personal Income" variables. Educational level < high school diploma is the reference group for the "Educational Level" variables. The models are computed with robust covariance matrix estimation.

**Table S25.** Logistic regression predicting vaping (not vaping=0, vaping=1) (combined samples).

|  | *SEX* |  |  | *GENDER* |  |  |
| --- | --- | --- | --- | --- | --- | --- |
|  | Odds ratio | 99.9% CI |  | Odds ratio | 99.9% CI |  |
| Caregiver strain | 1.10 | 0.91 | 1.32 | 1.10 | 0.91 | 1.32 |
| Work strain | 1.27 | 0.98 | 1.64 | 1.26 | 0.98 | 1.63 |
| Independence | 1.15 | 0.86 | 1.52 | 1.15 | 0.87 | 1.53 |
| Risk-taking | 1.10 | 0.86 | 1.41 | 1.10 | 0.86 | 1.40 |
| Emotional intelligence | 1.25 | 0.97 | 1.60 | 1.25 | 0.97 | 1.60 |
| Social support | 0.93 | 0.78 | 1.11 | 0.93 | 0.78 | 1.12 |
| Discrimination | 1.44 | 1.13 | 1.82 | 1.44 | 1.14 | 1.83 |
| Year of birth | 1.00 | 0.99 | 1.02 | 1.00 | 0.99 | 1.02 |
| Personal Income (\$10,000-\$19,999) | 1.59 | 0.70 | 3.57 | 1.55 | 0.70 | 3.47 |
| Personal Income (\$20,000-\$29,999) | 1.80 | 0.83 | 3.90 | 1.75 | 0.81 | 3.76 |
| Personal Income (\$30,000-\$39,999) | 1.70 | 0.76 | 3.78 | 1.65 | 0.75 | 3.65 |
| Personal Income (\$40,000-\$49,999) | 1.50 | 0.63 | 3.56 | 1.46 | 0.62 | 3.45 |
| Personal Income (\$50,000-\$59,999) | 1.93 | 0.83 | 4.49 | 1.89 | 0.82 | 4.36 |
| Personal Income (\$60,000-\$69,999) | 2.00 | 0.80 | 4.99 | 1.94 | 0.78 | 4.82 |
| Personal Income (\$70,000-\$79,999) | 1.27 | 0.44 | 3.64 | 1.23 | 0.43 | 3.51 |
| Personal Income (\$80,000-\$89,999) | 0.89 | 0.19 | 4.08 | 0.86 | 0.19 | 3.94 |
| Personal Income (\$90,000-\$99,999) | 2.26 | 0.68 | 7.48 | 2.17 | 0.66 | 7.13 |
| Personal Income (\$100,000-\$149,999) | 1.53 | 0.55 | 4.25 | 1.49 | 0.54 | 4.11 |
| Personal Income (More than \$150,000) | 4.37 | 1.60 | 11.96 | 4.41 | 1.63 | 11.95 |
| Educational Level (High school graduate) | 0.48 | 0.12 | 1.93 | 0.48 | 0.12 | 1.93 |
| Educational Level (Some college credit, no degree) | 0.43 | 0.11 | 1.65 | 0.44 | 0.12 | 1.67 |
| Educational Level (Trade/Technical/Vocational education) | 0.34 | 0.06 | 1.84 | 0.35 | 0.06 | 1.86 |
| Educational Level (Associate degree) | 0.54 | 0.14 | 2.14 | 0.55 | 0.14 | 2.16 |
| Educational Level (Bachelor's degree) | 0.31 | 0.08 | 1.17 | 0.32 | 0.08 | 1.20 |
| Educational Level (Master's degree) | 0.35 | 0.09 | 1.40 | 0.36 | 0.09 | 1.43 |
| Educational Level (professional degree) | 0.35 | 0.05 | 2.53 | 0.35 | 0.05 | 2.53 |
| Educational Level (Doctorate degree) | 0.36 | 0.07 | 1.93 | 0.37 | 0.07 | 1.97 |
| Ethnicity (White) | 1.38 | 0.65 | 2.95 | 1.31 | 0.61 | 2.79 |
| Ethnicity (Hispanic, Latinx, Spanish) | 1.28 | 0.55 | 2.97 | 1.25 | 0.53 | 2.91 |
| Ethnicity (Black, African American) | 1.00 | 0.41 | 2.45 | 1.01 | 0.41 | 2.46 |
| Ethnicity (Asian) | 0.85 | 0.33 | 2.20 | 0.80 | 0.30 | 2.10 |
| Ethnicity (Native) | 1.55 | 0.55 | 4.33 | 1.58 | 0.56 | 4.41 |
| Sex (male=0, female=1) | 0.57 | 0.37 | 0.88 |  |  |  |
| Woman |  |  |  | 0.56 | 0.37 | 0.86 |
| Non-binary/Gender fluid + Gender=other |  |  |  | 0.42 | 0.05 | 3.62 |
| Sample (Sample 1=1) | 0.89 | 0.51 | 1.55 | 0.89 | 0.52 | 1.55 |
| **Goodness of Fit** |  |  |  |  |  |  |
| Deviance/df | 0.618 |  |  | 0.618 |  |  |
| Pearson Chi-Square/df | 1.027 |  |  | 1.033 |  |  |
| Loglikelihood | 2451.495 |  |  | -1194.642 |  |  |
| AIC | 2452.148 |  |  | 2461.285 |  |  |
| **Omnibus tests** |  |  |  |  |  |  |
| Chi-square | 174.129 |  |  | 176.985 |  |  |
| df | 34 |  |  | 35 |  |  |
| N | 3891 |  |  | 3900 |  |  |

Note: Participants reporting Native Hawaiian/Pacific Islander, Middle east/North Africa or Other as ethnicity are the reference group for the ethnicity variables. Personal Income < \$10,000 is the reference group for the "Personal Income" variables. Educational level < high school diploma is the reference group for the "Educational Level" variables. The models are computed with robust covariance matrix estimation.

**Table S26.** Logistic regression predicting binge drinking (less than monthly=0, monthly, weekly, and daily=1) (combined samples).

|  | *SEX* |  |  | *GENDER* |  |  |
| --- | --- | --- | --- | --- | --- | --- |
|  | Odds ratio | 99.9% CI |  | Odds ratio | 99.9% CI |  |
| Caregiver strain | 0.93 | 0.80 | 1.09 | 0.93 | 0.79 | 1.09 |
| Work strain | 1.15 | 0.95 | 1.39 | 1.15 | 0.95 | 1.39 |
| Independence | 1.12 | 0.92 | 1.36 | 1.12 | 0.91 | 1.36 |
| Risk-taking | 1.37 | 1.15 | 1.64 | 1.37 | 1.15 | 1.64 |
| Emotional intelligence | 1.12 | 0.93 | 1.35 | 1.12 | 0.93 | 1.35 |
| Social support | 0.94 | 0.82 | 1.07 | 0.93 | 0.82 | 1.07 |
| Discrimination | 1.16 | 0.95 | 1.42 | 1.16 | 0.96 | 1.42 |
| Year of birth | 1.02 | 1.01 | 1.04 | 1.02 | 1.01 | 1.04 |
| Personal Income (\$10,000-\$19,999) | 1.16 | 0.67 | 2.00 | 1.16 | 0.67 | 2.01 |
| Personal Income (\$20,000-\$29,999) | 1.60 | 0.94 | 2.70 | 1.59 | 0.94 | 2.68 |
| Personal Income (\$30,000-\$39,999) | 1.29 | 0.74 | 2.26 | 1.29 | 0.74 | 2.26 |
| Personal Income (\$40,000-\$49,999) | 1.58 | 0.89 | 2.80 | 1.58 | 0.89 | 2.81 |
| Personal Income (\$50,000-\$59,999) | 1.14 | 0.63 | 2.09 | 1.15 | 0.63 | 2.11 |
| Personal Income (\$60,000-\$69,999) | 1.54 | 0.80 | 2.95 | 1.53 | 0.80 | 2.95 |
| Personal Income (\$70,000-\$79,999) | 1.11 | 0.54 | 2.28 | 1.11 | 0.54 | 2.28 |
| Personal Income (\$80,000-\$89,999) | 1.19 | 0.47 | 3.02 | 1.19 | 0.47 | 3.03 |
| Personal Income (\$90,000-\$99,999) | 1.91 | 0.80 | 4.56 | 1.91 | 0.80 | 4.56 |
| Personal Income (\$100,000-\$149,999) | 1.50 | 0.74 | 3.04 | 1.50 | 0.74 | 3.05 |
| Personal Income (More than \$150,000) | 2.72 | 1.16 | 6.35 | 2.74 | 1.17 | 6.41 |
| Educational Level (High school graduate) | 0.81 | 0.22 | 2.98 | 0.80 | 0.22 | 2.97 |
| Educational Level (Some college credit, no degree) | 1.28 | 0.37 | 4.49 | 1.29 | 0.37 | 4.53 |
| Educational Level (Trade/Technical/Vocational education) | 1.39 | 0.34 | 5.62 | 1.38 | 0.34 | 5.59 |
| Educational Level (Associate degree) | 1.00 | 0.27 | 3.67 | 1.00 | 0.27 | 3.65 |
| Educational Level (Bachelor's degree) | 1.04 | 0.30 | 3.63 | 1.05 | 0.30 | 3.65 |
| Educational Level (Master's degree) | 1.12 | 0.31 | 4.03 | 1.12 | 0.31 | 4.05 |
| Educational Level (professional degree) | 1.37 | 0.26 | 7.05 | 1.37 | 0.26 | 7.04 |
| Educational Level (Doctorate degree) | 1.22 | 0.27 | 5.46 | 1.24 | 0.28 | 5.52 |
| Ethnicity (White) | 1.35 | 0.72 | 2.55 | 1.34 | 0.71 | 2.53 |
| Ethnicity (Hispanic, Latinx, Spanish) | 1.19 | 0.63 | 2.23 | 1.18 | 0.63 | 2.23 |
| Ethnicity (Black, African American) | 0.98 | 0.46 | 2.06 | 0.97 | 0.46 | 2.05 |
| Ethnicity (Asian) | 0.53 | 0.25 | 1.17 | 0.53 | 0.24 | 1.15 |
| Ethnicity (Native) | 0.62 | 0.24 | 1.59 | 0.64 | 0.25 | 1.64 |
| Sex (male=0, female=1) | 0.65 | 0.48 | 0.89 |  |  |  |
| Woman |  |  |  | 0.67 | 0.49 | 0.91 |
| Non-binary/Gender fluid + Gender=other |  |  |  | 0.34 | 0.07 | 1.74 |
| Sample (Sample 1=1) | 1.18 | 0.80 | 1.74 | 1.18 | 0.80 | 1.74 |
| **Goodness of Fit** |  |  |  |  |  |  |
| Deviance/df | 1.005 |  |  | 1.005 |  |  |
| Pearson Chi-Square/df | 1.015 |  |  | 1.014 |  |  |
| Loglikelihood | -1936.517 |  |  | -1936.050 |  |  |
| AIC | 3943.033 |  |  | 3944.100 |  |  |
| **Omnibus tests** |  |  |  |  |  |  |
|  | 282.699 |  |  | 283.632 |  |  |
| Chi-square |  |  |  |  |  |  |
| df | 34 |  |  | 35 |  |  |
| N | 3887 |  |  | 3887 |  |  |

Note: Participants reporting Native Hawaiian/Pacific Islander, Middle east/North Africa or Other as ethnicity are the reference group for the ethnicity variables. Personal Income < \$10,000 is the reference group for the "Personal Income" variables. Educational level < high school diploma is the reference group for the "Educational Level" variables. The models are computed with robust covariance matrix estimation.

**Table S27.** Logistic regression predicting Overweight (BMI<25=0, BMI≥25 =1) (combined samples).

|  | *SEX* |  |  | *GENDER* |  |  |
| --- | --- | --- | --- | --- | --- | --- |
|  | Odds ratio | 99.9% CI |  | Odds ratio | 99.9% CI |  |
| Caregiver strain | 1.21 | 1.07 | 1.38 | 1.22 | 1.07 | 1.38 |
| Work strain | 1.10 | 0.95 | 1.29 | 1.10 | 0.94 | 1.29 |
| Independence | 1.01 | 0.85 | 1.20 | 1.01 | 0.85 | 1.19 |
| Risk-taking | 0.84 | 0.72 | 0.97 | 0.84 | 0.72 | 0.97 |
| Emotional intelligence | 1.00 | 0.85 | 1.18 | 1.00 | 0.85 | 1.18 |
| Social support | 1.04 | 0.93 | 1.17 | 1.04 | 0.93 | 1.16 |
| Discrimination | 1.24 | 1.03 | 1.48 | 1.23 | 1.03 | 1.47 |
| Year of birth | 0.97 | 0.97 | 0.98 | 0.97 | 0.97 | 0.98 |
| Personal Income (\$10,000-\$19,999) | 0.96 | 0.63 | 1.46 | 0.97 | 0.64 | 1.47 |
| Personal Income (\$20,000-\$29,999) | 1.01 | 0.66 | 1.53 | 1.02 | 0.67 | 1.55 |
| Personal Income (\$30,000-\$39,999) | 0.92 | 0.59 | 1.43 | 0.94 | 0.61 | 1.47 |
| Personal Income (\$40,000-\$49,999) | 0.93 | 0.59 | 1.48 | 0.95 | 0.60 | 1.50 |
| Personal Income (\$50,000-\$59,999) | 1.30 | 0.79 | 2.13 | 1.32 | 0.81 | 2.17 |
| Personal Income (\$60,000-\$69,999) | 1.28 | 0.72 | 2.25 | 1.30 | 0.74 | 2.30 |
| Personal Income (\$70,000-\$79,999) | 1.71 | 0.94 | 3.12 | 1.74 | 0.96 | 3.18 |
| Personal Income (\$80,000-\$89,999) | 1.08 | 0.51 | 2.29 | 1.10 | 0.52 | 2.33 |
| Personal Income (\$90,000-\$99,999) | 1.38 | 0.62 | 3.09 | 1.39 | 0.63 | 3.07 |
| Personal Income (\$100,000-\$149,999) | 1.03 | 0.55 | 1.90 | 1.05 | 0.57 | 1.94 |
| Personal Income (More than \$150,000) | 1.09 | 0.47 | 2.54 | 1.08 | 0.47 | 2.51 |
| Educational Level (High school graduate) | 0.59 | 0.20 | 1.72 | 0.59 | 0.20 | 1.72 |
| Educational Level (Some college credit, no degree) | 0.68 | 0.24 | 1.92 | 0.68 | 0.24 | 1.92 |
| Educational Level (Trade/Technical/Vocational education) | 0.53 | 0.16 | 1.73 | 0.53 | 0.16 | 1.72 |
| Educational Level (Associate degree) | 0.60 | 0.21 | 1.77 | 0.60 | 0.20 | 1.75 |
| Educational Level (Bachelor's degree) | 0.45 | 0.16 | 1.28 | 0.45 | 0.16 | 1.27 |
| Educational Level (Master's degree) | 0.52 | 0.18 | 1.52 | 0.51 | 0.17 | 1.50 |
| Educational Level (professional degree) | 0.33 | 0.08 | 1.30 | 0.33 | 0.08 | 1.30 |
| Educational Level (Doctorate degree) | 0.39 | 0.11 | 1.38 | 0.39 | 0.11 | 1.37 |
| Ethnicity (White) | 1.05 | 0.61 | 1.81 | 1.07 | 0.62 | 1.85 |
| Ethnicity (Hispanic, Latinx, Spanish) | 1.42 | 0.80 | 2.50 | 1.44 | 0.82 | 2.53 |
| Ethnicity (Black, African American) | 1.38 | 0.74 | 2.58 | 1.39 | 0.75 | 2.59 |
| Ethnicity (Asian) | 0.50 | 0.27 | 0.94 | 0.51 | 0.27 | 0.95 |
| Ethnicity (Native) | 1.55 | 0.69 | 3.44 | 1.57 | 0.70 | 3.50 |
| Sex (male=0, female=1) | 0.67 | 0.51 | 0.87 |  |  |  |
| Woman |  |  |  | 0.68 | 0.52 | 0.88 |
| Non-binary/Gender fluid + Gender=other |  |  |  | 0.76 | 0.24 | 2.42 |
| Sample (Sample 1=1) | 0.90 | 0.65 | 1.24 | 0.90 | 0.66 | 1.25 |
| **Goodness of Fit** |  |  |  |  |  |  |
| Deviance/df | 1.296 |  |  | 1.296 |  |  |
| Pearson Chi-Square/df | 1.010 |  |  | 1.010 |  |  |
| Loglikelihood | -2472.355 |  |  | -2478.706 |  |  |
| AIC | 5014.711 |  |  | 5029.412 |  |  |
| **Omnibus tests** |  |  |  |  |  |  |
|  | 267.790 |  |  | 268.953 |  |  |
| Chi-square |  |  |  |  |  |  |
| df | 34 |  |  | 35 |  |  |
| N | 3851 |  |  | 3860 |  |  |

Note: Participants reporting Native Hawaiian/Pacific Islander, Middle east/North Africa or Other as ethnicity are the reference group for the ethnicity variables. Personal Income < \$10,000 is the reference group for the “Personal Income” variables. Educational level < high school diploma is the reference group for the “Educational Level” variables. The models are computed with robust covariance matrix estimation.

**Table S28.**
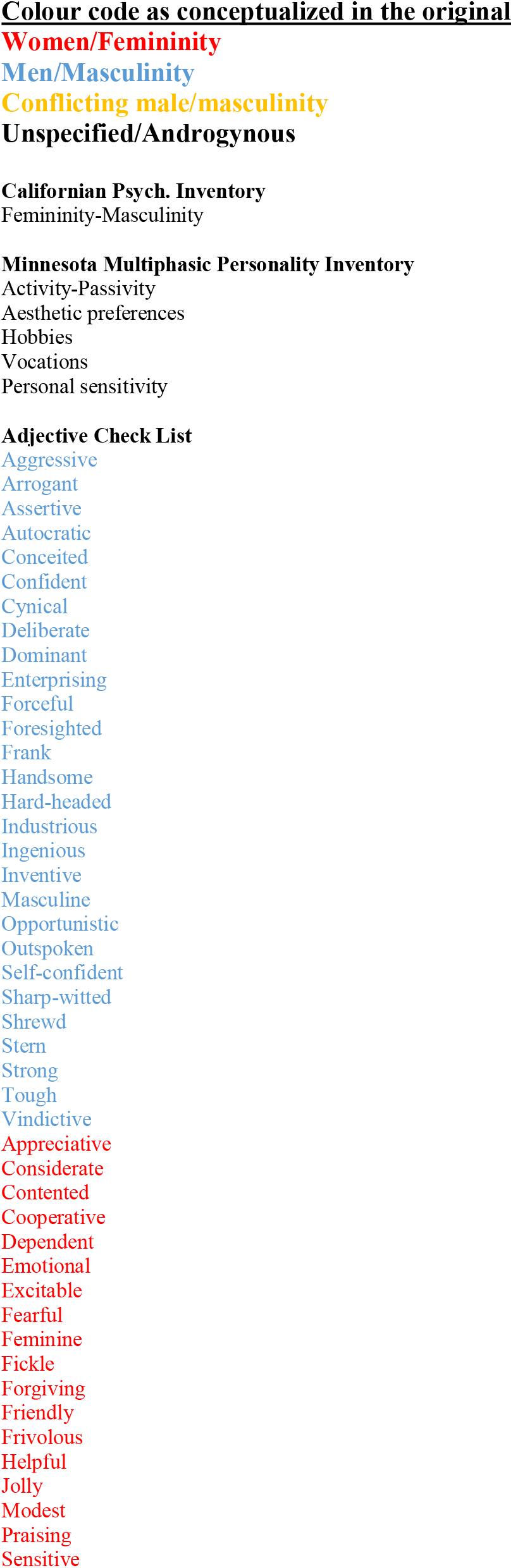
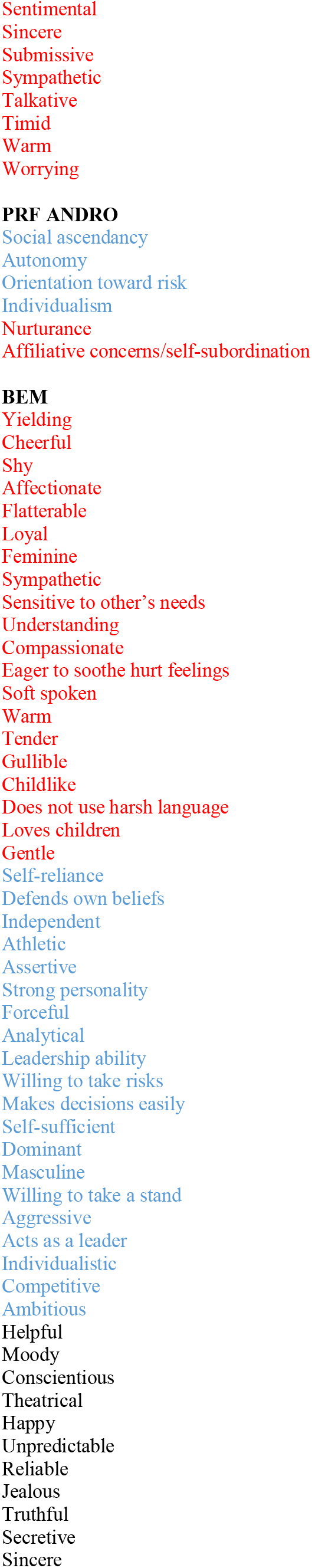
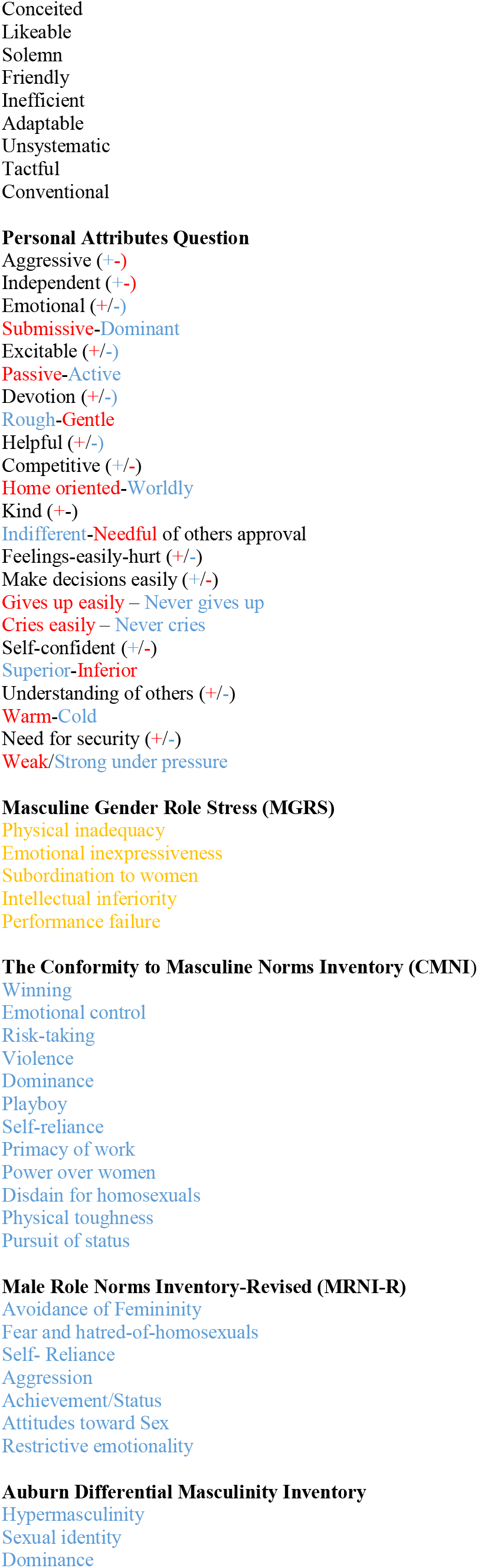
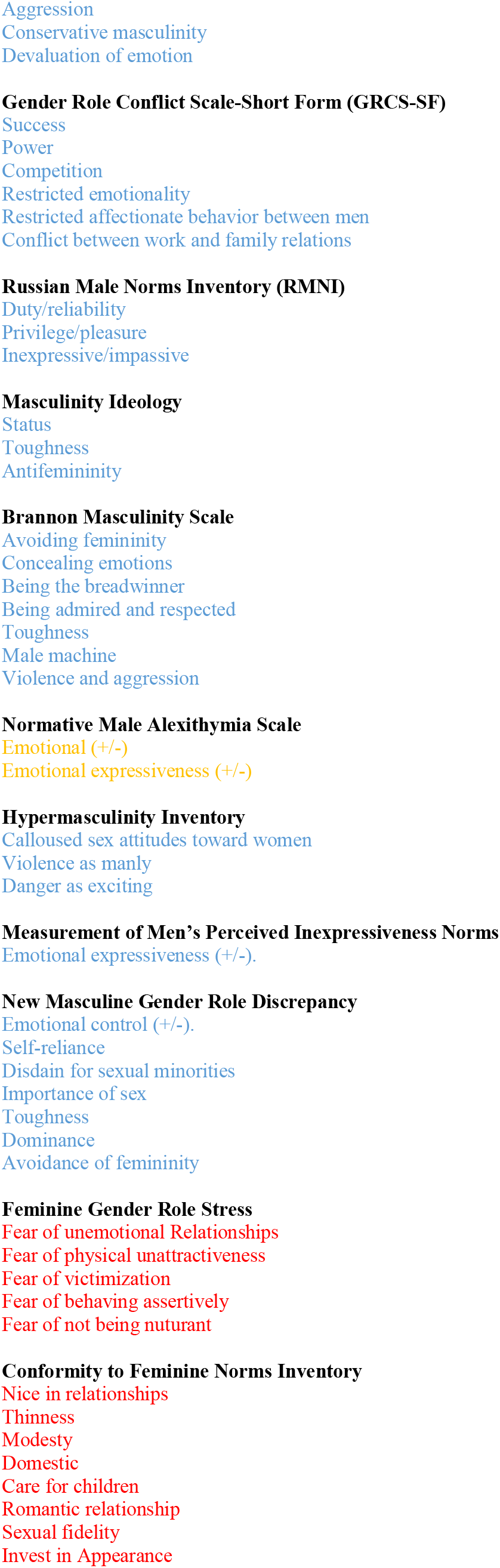
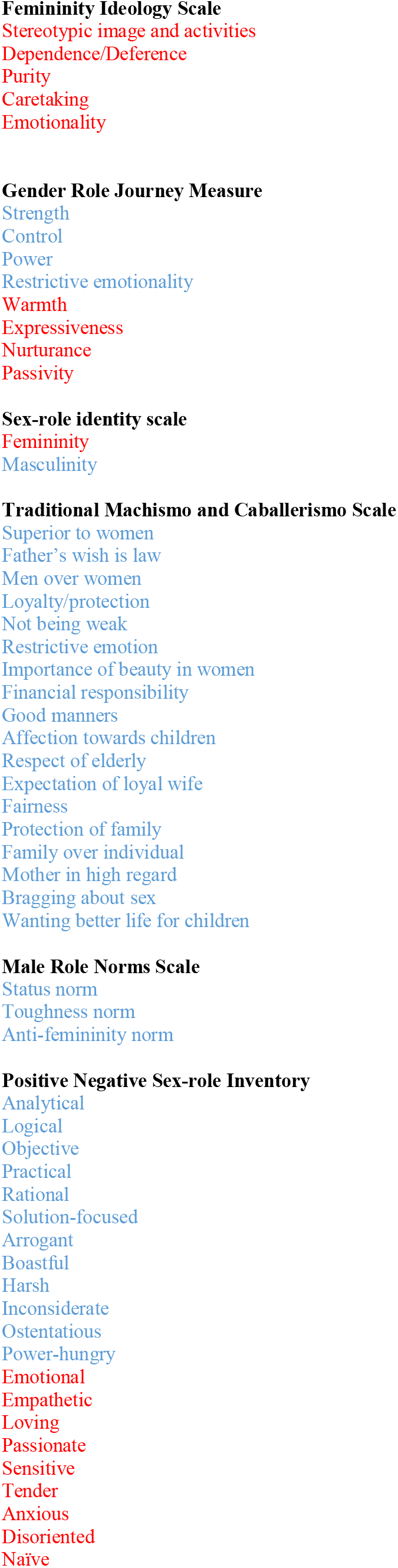
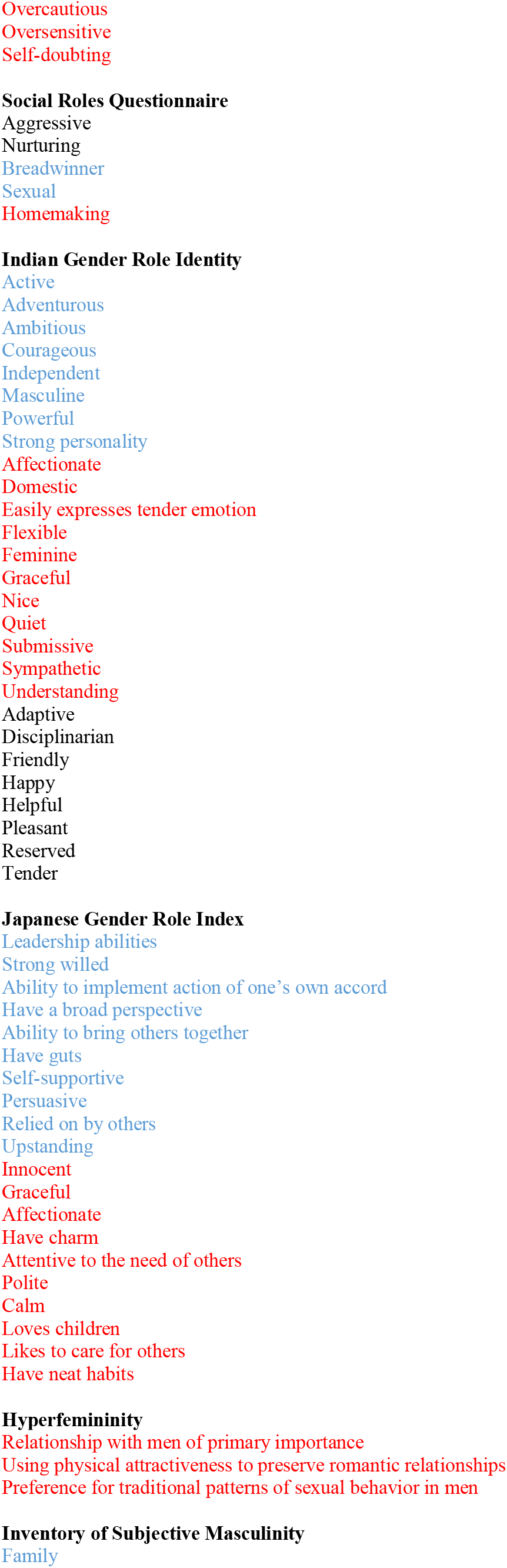
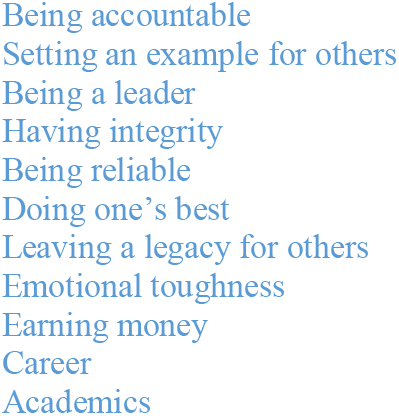
Gender characteristics

**Table S29.** Rank of gender characteristics based on occurrences (>2).

|  |  |
| --- | --- |
| Expressiveness and emotionality (37) | Emotional, Emotional, Emotional (+/-), Emotional control, Emotional Control, Emotional expressiveness (+/-), Emotional expressiveness (+/-), Emotional inexpressiveness, Emotionality, Expressiveness, Excitable, Excitable, Restricted emotionality, Restrictive emotion, Restrictive Emotionality, Restrictive emotionality, Inexpressive/Impassive, Concealing emotions, Feelings-easily-hurt (+/-), Fear of Unemotional Relationships, Personal sensitivity, Sensitive, Sentimental, Cries easily – Never cries, Secretive, Moody, Devaluation of emotion, Affection towards children, Affectionate, Restricted affectionate behavior between men, Devotion, Compassionate, Emotional, Sensitive, Affectionate, Easily expresses tender emotion, Affectionate |
| Empathy, caring and nurturing (37) | Warm, Warm, Warm, Warmth, Friendly, Friendly, Helpful, Helpful (+-) Helpful, Likeable, Kind, Forgiving, Appreciative, Praising, Care for children, Caretaking, Affection towards children, Loves children, Fear of not Being Nurturant, Eager to soothe hurt feelings, Sensitive to other's needs, Nurturance, Home oriented, Domestic, Nurturance, Affiliative concerns, Understanding, Understanding of others, Tender, Empathetic, Tender, Nurturing, Understanding, Sympathetic, Helpful, Tender, Attentive to the needs of others, Loves children, Likes to care for others |
| Self-reliance and independence (18) | Self-reliance, Self-reliance, Self-reliance, Self-Reliance, Self-sufficient, Indifferent-Needful of others approval (+/-), Independent, Independent, Individualistic, Dependence/Deference, Dependent, Autonomy, Individualism, Self-confident, Self-confident, Confident, Independent, Ability to implement action of one's own accord, Self-supportive |
| Striving for status, competition, (17) | Achievement/Status, Pursuit of status, Status, Being admired and respected, Competition, Competitive, Competitive, Winning, Ambitious, Gives up easily/Never gives up, Power, Power, superior, Social Ascendancy, Status norm, Power-hungry, Ambitious |
| Dominance (9) | Dominance, Dominance, Dominance, Dominant, Dominant, Forceful, Forceful, Autocratic, Control |
| Toughness (9) | Tough, Toughness, Toughness, Toughness, Weak/Strong under pressure, Physical inadequacy, Physical toughness, Not being weak, Toughness norm, Emotional toughness, |
| Decision-making (11) | Willing to take a stand, Make decisions easily (+/-), Makes decisions easily, Acts as a leader, Foresighted, Defends own beliefs, Hard-headed, Leadership ability, Solution-focused, Leadership abilities, Being a leader |
| Aggressiveness (7) | Aggression, Aggressive, Aggressive, Aggressive, Aggression, Aggression, Aggressive |
| Assertive (6) | Assertive, Assertive, Fear of Behaving Assertively, Outspoken, Frank, Boastful, Ostentatious |
| Truthful, sincere, reliable (5) | Truthful, Sincere, Sincere, Reliable, Conscientious |
| Modesty (6) | Modesty, Shy, Modest, Gentle, Timid, Quiet |
| Risk (5) | Willing to take risks, Danger as exciting, Risk-taking, Enterprising, Orientation Toward Risk |
| Adaptable, gullible (5) | Adaptable, Gullible, Fickle, Yielding, Adaptive |
| Worry, fearful (3) | Worrying, Fearful, Need for security (+/-), Overcautious |
| Active-Passive (4) | Passive-Active, Passivity, Activity-Passivity, Active |
| Violence (3) | Violence, Violence, Violence as manly |
| Avoidance of femininity (3) | Avoidance of Femininity, Avoidance of Femininity, Avoiding femininity, Anti-femininity. |

**Table S30.** Search-terms for meta-analyses of existing scales measuring each gender variable.

| Gender Variable | Search terms |
| --- | --- |
| Caregiving | caregiv* [pubmed]; (caregiv* OR care*) [psychinfo] |
| Time Use | ("time use" OR "time poverty" OR "time deficit" OR "leisure time" OR "unpaid work" OR "domestic work" OR "leisure time physical activity" OR "physical activity" OR "paid work" OR "sleep" OR "sleep time" OR "sleep patterns") |
| Occupation | ("Occupational Segregation" OR "Occupational Preferences" OR "Occupational Patterns" OR "demand-control" OR "emotional labor" OR occupation OR profession OR "Job strain" OR "job hazard*" OR "occupational stratification") |
| Competitive/striving | ("competitive attitude" OR competit* OR success OR power OR winning OR striving) |
| Self-reliant/independent | ("self-efficacy" OR "self-determination" OR agen* or assertive or adventurous OR "self-reliance" OR "self-reliant") |
| Sensation-seeking/risk-taking | ("risk-taking" OR "sensation-seeking" OR "risky behavior") |
| Communal | (collective* OR "communal orientation" OR communal* OR agreeable OR interdependen* OR nurtur* OR "tender-minded*" OR connected) |
| Empathetic | (empath* OR "emotional intelligence") |
| Expressive | (emotion* OR express* OR secretive OR extrover* OR introver* OR "restrictive emotionality" OR "restrictive emotional behavior" OR "affectionate behavior" OR affection OR inexpression OR impassive OR passivity OR "emotional control") |
| Social Support | "social support" |
| Gender Discrimination | ("gender discrimination" OR discriminat*) |

**Table S31.** Health-related items and response options.

| Item name | Item | Response categories |
| --- | --- | --- |
| General health | In general, would you say your health is... | 1= Excellent, 2= Very good, 3=Good, 4=fair, 5=poor (recoded to dummy variable: excellent, very good, good=0, fair, poor=1) |
| Physical health | Now thinking about your physical health, which includes physical illness and injury, how many days during the past 30 days were your physical health not good? | Number of days (1-30) |
| Mental health | Now thinking about your mental health, which includes stress, depression, and problems with emotions, how many days during the past 30 days were your mental health not good? | Number of days (1-30) |
| Activity limitations | During the past 30 days, approximately how many days did poor physical or mental health keep you from doing your usual activities, such as self-care, work, or recreation? | Number of days (1-30) |
| Smoking | How many cigarettes do you smoke per day, not including e-cigarettes? | Number of cigarettes (recoded to dummy variable: not smoking=0, smoking=1) |
| Vaping | How many times do you vape or use an e-cigarette product per day? | Number of times vaping (recoded to dummy variable: not vaping=0, vaping=1) |
| Binge drinking | Men: In the past 3 months, how often have you had 5 or more drinks on one occasion?<br>Women: In the past 3 months, how often have you had 4 or more drinks on one occasion? | 1=Never, 2=Less than monthly, 3=Monthly, 4=Weekly, 5=Daily or almost daily (recoded to unisex dummy variable: binge drinking less than monthly=0, binge drinking monthly, weekly or daily/almost daily=1) |
| BMI | [BMI is calculated based on self-reported height and weight] | BMI recoded to Dummy: BMI $\leq 25$ = 0, BMI > 25 = 1 |

**Table S32.**
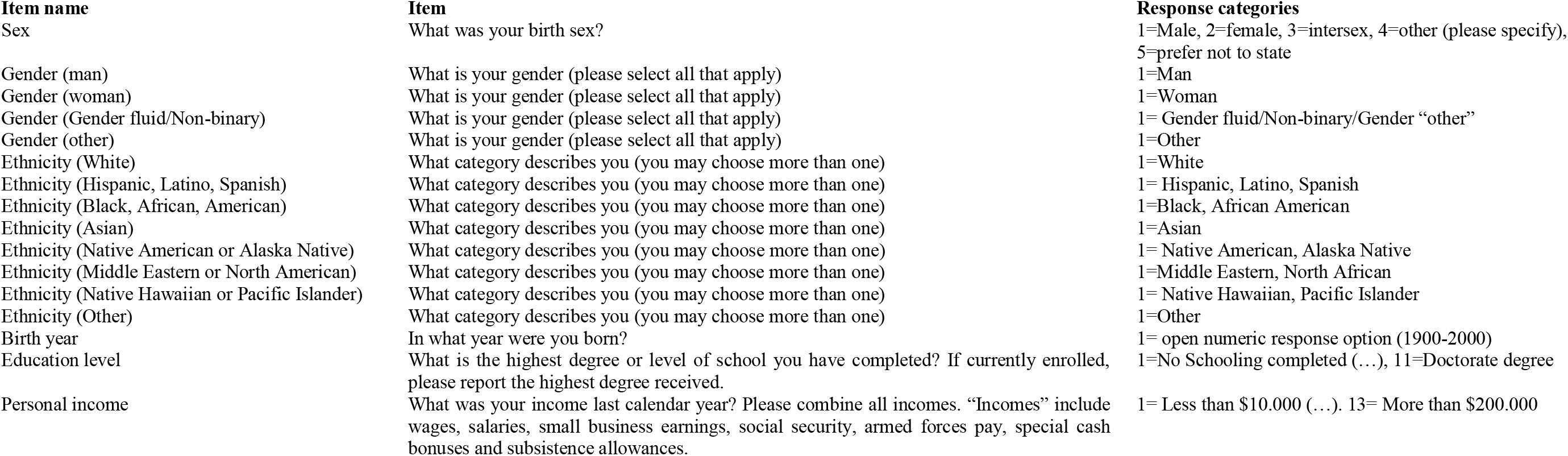
Demographic items and response options.

| Item name | Item | Response categories |
| --- | --- | --- |
| Sex | What was your birth sex? | 1=Male, 2=female, 3=intersex, 4=other (please specify), 5=prefer not to state |
| Gender (man) | What is your gender (please select all that apply) | 1=Man |
| Gender (woman) | What is your gender (please select all that apply) | 1=Woman |
| Gender (Gender fluid/Non-binary) | What is your gender (please select all that apply) | 1= Gender fluid/Non-binary/Gender “other” |
| Gender (other) | What is your gender (please select all that apply) | 1=Other |
| Ethnicity (White) | What category describes you (you may choose more than one) | 1=White |
| Ethnicity (Hispanic, Latino, Spanish) | What category describes you (you may choose more than one) | 1= Hispanic, Latino, Spanish |
| Ethnicity (Black, African, American) | What category describes you (you may choose more than one) | 1=Black, African American |
| Ethnicity (Asian) | What category describes you (you may choose more than one) | 1=Asian |
| Ethnicity (Native American or Alaska Native) | What category describes you (you may choose more than one) | 1= Native American, Alaska Native |
| Ethnicity (Middle Eastern or North African) | What category describes you (you may choose more than one) | 1=Middle Eastern, North African |
| Ethnicity (Native Hawaiian or Pacific Islander) | What category describes you (you may choose more than one) | 1= Native Hawaiian, Pacific Islander |
| Ethnicity (Other) | What category describes you (you may choose more than one) | 1=Other |
| Birth year | In what year were you born? | 1= open numeric response option (1900-2000) |
| Education level | What is the highest degree or level of school you have completed? If currently enrolled, please report the highest degree received. | 1=No Schooling completed (...), 11=Doctorate degree |
| Personal income | What was your income last calendar year? Please combine all incomes. “Incomes” include wages, salaries, small business earnings, social security, armed forces pay, special cash bonuses and subsistence allowances. | 1= Less than \$10.000 (...). 13= More than \$200.000 |

**Table S33.** Recoding of ten variables to allow for the largest possible sample in the EFA.

|  |
| --- |
| **Caregiving items**<br>The group of current caregivers in sample 1 consists of 605 respondents. To utilize the largest possible sample in the EFA, we recoded the following three items so that people not currently taking care of someone in need were ascribed the value 1 (same procedure was used in samples 2 and 3) |
| **careemot.** In the past year, how often did you feel emotionally exhausted because of your caretaking responsibilities? 1= **Never**, 2= Once in a while, 3= Sometimes, 4= Most of the time, 5=Always |
| **carephys.** In the past year, how often did you feel physically exhausted because of your caretaking responsibilities? 1= **Never**, 2= Once in a while, 3= Sometimes, 4= Most of the time, 5=Always |
| **carefut.** In the past year, how often have your caretaking responsibilities caused you to worry about the future?<br>1= **Never**, 2= Once in a while, 3= Sometimes, 4= Most of the time, 5=Always |
| **Work items**<br>The group of current employees in sample 1 consists of 1462 respondents. To utilize the largest possible sample in the EFA, we recoded the following six items so that people not currently employed were ascribed the value 1 (same procedure was used in sample 2) |
| **workspeed.** How often does your job require working fast? 1= **Never**, 2= Once in a while, 3= Sometimes, 4= Most of the time, 5=Always |
| **workrep.** How often does your job involve repetitive tasks? 1= **Never**, 2= Once in a while, 3= Sometimes, 4= Most of the time, 5=Always |
| **workemot.** How often do you feel emotionally exhausted from your work activities? 1= **Never**, 2= Once in a while, 3= Sometimes, 4= Most of the time, 5=Always |
| **workphys.** How often do you feel physically exhausted from your work activities? 1= **Never**, 2= Once in a while, 3= Sometimes, 4= Most of the time, 5=Always |
| **workrisk.** How often does your job involve risk of harm or injury? 1= **Never**, 2= Once in a while, 3= Sometimes, 4= Most of the time, 5=Always |
| **workhaz.** How often does your job involve hazards, such as smoke, heat, noise, or chemicals? 1= **Never**, 2= Once in a while, 3= Sometimes, 4= Most of the time, 5=Always |
| **Discrimination**<br>258 respondents have never worked and were not asked the question about perceived discrimination when getting hired (discrhire) in sample 1. To utilize the largest possible sample in the EFA, we recoded this item so that respondents that have never worked were ascribed the value 1 (all respondents were asked this question in sample 2, so no recoding was necessary). |
| **discrhire.** Because of your gender, how often have you felt discriminated against, when getting hired? 1= **Never**, 2= Once in a while, 3= Sometimes, 4= Most of the time, 5=Always |

**Table S34.** Exploratory Factor Analysis (Full factor model).

|  | 1 | 2 | 3 | 4 | 5 | 6 | 7 |
| --- | --- | --- | --- | --- | --- | --- | --- |
| *discrgen* | **0.853** | -0.043 | -0.060 | -0.077 | 0.093 | -0.003 | 0.093 |
| *discrpub* | **0.840** | 0.002 | -0.056 | -0.063 | 0.055 | 0.012 | 0.035 |
| *discred* | **0.811** | -0.031 | -0.049 | 0.033 | -0.054 | 0.040 | -0.007 |
| *discrmed* | **0.753** | 0.012 | 0.033 | -0.010 | -0.050 | 0.047 | -0.008 |
| *discrhire* | **0.728** | 0.082 | 0.021 | 0.009 | -0.039 | 0.066 | 0.070 |
| *discrfam* | **0.624** | -0.019 | 0.047 | 0.002 | -0.003 | -0.128 | -0.042 |
| *workspeed* | -0.041 | **0.831** | -0.006 | -0.046 | 0.084 | 0.053 | 0.068 |
| *workphys* | 0.017 | **0.816** | 0.020 | -0.014 | -0.003 | -0.086 | -0.066 |
| *workemot* | 0.052 | **0.800** | 0.019 | -0.086 | 0.018 | -0.058 | -0.019 |
| *workrep* | -0.031 | **0.799** | 0.003 | -0.065 | 0.040 | 0.014 | 0.039 |
| *timework* | -0.120 | **0.582** | -0.155 | -0.024 | 0.016 | 0.071 | 0.122 |
| *workrisk* | 0.126 | **0.493** | 0.039 | 0.182 | -0.141 | -0.014 | -0.101 |
| *workhaz* | 0.116 | **0.427** | 0.026 | 0.167 | -0.118 | -0.042 | -0.102 |
| *timeleisure* | 0.042 | -0.358 | -0.343 | 0.015 | -0.034 | -0.127 | -0.128 |
| *timehouse* | 0.069 | -0.230 | 0.130 | 0.056 | 0.060 | 0.019 | 0.068 |
| *timecom* | -0.003 | 0.162 | -0.061 | 0.150 | 0.058 | -0.069 | 0.014 |
| *carephys* | -0.010 | -0.001 | **0.938** | 0.007 | -0.020 | 0.008 | -0.025 |
| *careemot* | -0.003 | -0.006 | **0.933** | 0.001 | 0.015 | -0.027 | -0.015 |
| *carefut* | -0.018 | -0.006 | **0.903** | 0.003 | 0.000 | -0.021 | -0.011 |
| *timecare* | -0.006 | -0.126 | **0.703** | -0.080 | 0.055 | 0.069 | 0.068 |
| *timesleep* | 0.017 | -0.042 | -0.175 | -0.047 | -0.001 | 0.053 | -0.005 |
| *riskgen* | 0.029 | -0.047 | 0.000 | **0.754** | -0.050 | 0.113 | 0.123 |
| *riskfinan* | -0.031 | 0.057 | 0.025 | **0.670** | -0.149 | 0.109 | 0.028 |
| *riskrecrea* | 0.023 | 0.010 | -0.003 | **0.593** | -0.028 | 0.069 | 0.051 |
| *compgen* | -0.125 | -0.031 | 0.006 | **0.528** | 0.077 | -0.142 | 0.088 |
| *compdom* | -0.021 | -0.001 | 0.026 | **0.489** | 0.054 | -0.226 | -0.014 |
| *timeexc* | 0.019 | -0.072 | -0.120 | 0.243 | 0.044 | -0.008 | 0.013 |
| *commfriend* | 0.060 | 0.004 | -0.011 | -0.006 | **0.667** | -0.091 | 0.093 |
| *commemp* | 0.003 | 0.012 | -0.003 | 0.024 | **0.587** | -0.086 | 0.201 |
| *exprfriend* | -0.010 | 0.030 | -0.034 | 0.051 | **0.522** | -0.034 | -0.295 |
| *commdecis* | -0.032 | -0.035 | 0.124 | -0.170 | **0.466** | 0.037 | 0.049 |
| *exprother* | -0.049 | 0.008 | 0.001 | 0.101 | **0.434** | 0.199 | -0.110 |
| *exprown* | -0.042 | -0.006 | -0.016 | 0.022 | 0.367 | 0.252 | 0.137 |
| *socsuplone* | -0.064 | 0.002 | -0.057 | -0.060 | -0.143 | **0.709** | 0.064 |
| *socsuplove* | 0.098 | -0.030 | 0.058 | -0.073 | 0.188 | **0.619** | -0.063 |
| *socsupchores* | 0.112 | 0.010 | 0.087 | -0.004 | -0.028 | **0.510** | -0.191 |
| *qualfam* | -0.089 | -0.043 | -0.037 | 0.070 | 0.101 | 0.386 | -0.043 |
| *commother* | 0.019 | 0.016 | 0.004 | -0.048 | 0.240 | -0.386 | -0.198 |
| *socsupadv* | 0.059 | -0.011 | -0.033 | 0.000 | 0.235 | 0.296 | -0.233 |
| *qualfamarg* | -0.167 | 0.007 | -0.100 | -0.226 | 0.001 | 0.263 | 0.142 |
| *indepprob* | 0.051 | -0.001 | 0.039 | 0.193 | 0.194 | -0.006 | **0.706** |
| *indepgen* | 0.052 | 0.034 | -0.013 | 0.157 | 0.203 | 0.030 | **0.679** |
| *indephelp* | 0.002 | -0.013 | -0.011 | -0.049 | -0.133 | -0.044 | **0.568** |
| *Exprhelp (reverse scored)* | 0.048 | -0.038 | 0.053 | -0.196 | -0.132 | -0.207 | **0.489** |

Note: N=1999. KMO: 0.826; Bartlett's Test of Sphericity: $\chi^2 = 36185.003$ , $df = 946$ , $p < 0.001$ , Extraction method: Principal Axis Factoring. Rotation Method: Promax w. Kaiser Normalization. Rotation converged in 8 iterations. Total Variance Explained: 50.8%. Item loadings from the Pattern Matrix. The total variance explained by each factor was: Factor 1= 12.5%, Factor 2= 9.1%, Factor 3= 8.4%, Factor 4= 7.4%, Factor 5= 5.4%, Factor 6= 4.4%, Factor 7= 3.7%.

**Table S35.**
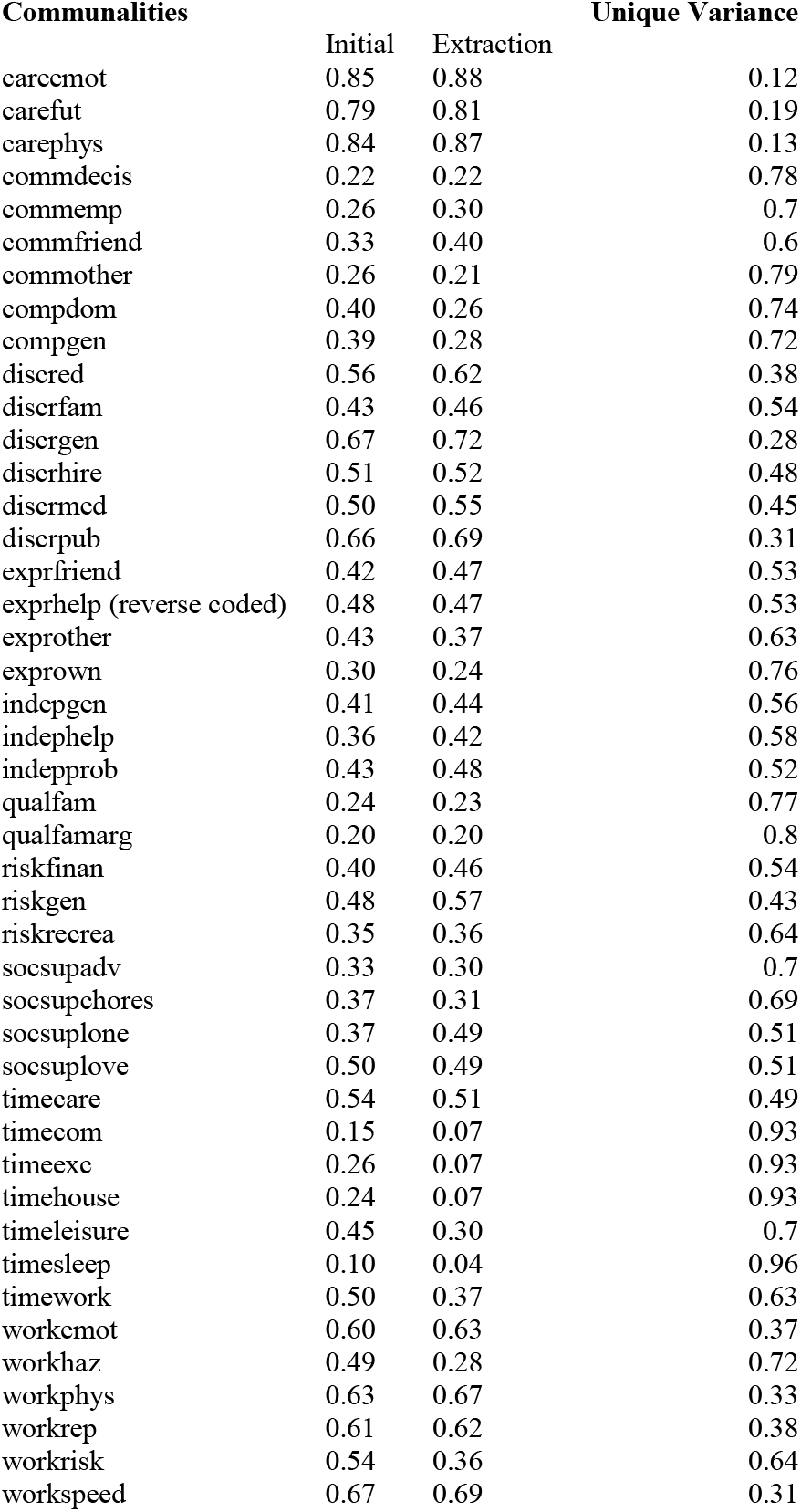
Communalities and unique variances for exploratory factor analysis presented in Table S34.

**Table S36.**
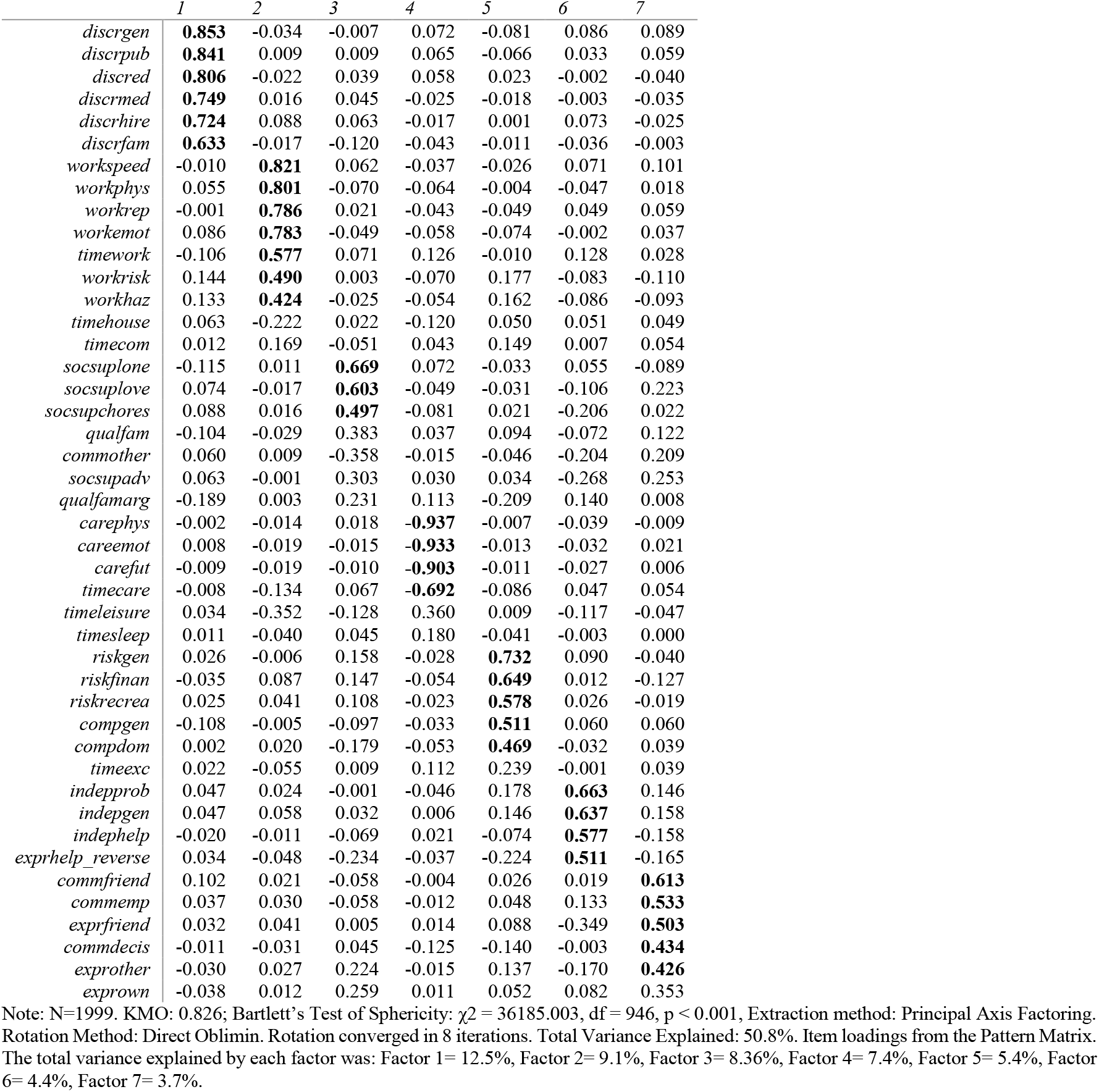
Exploratory Factor Analysis (Full factor model), Oblimin rotation.

**Table S37.**
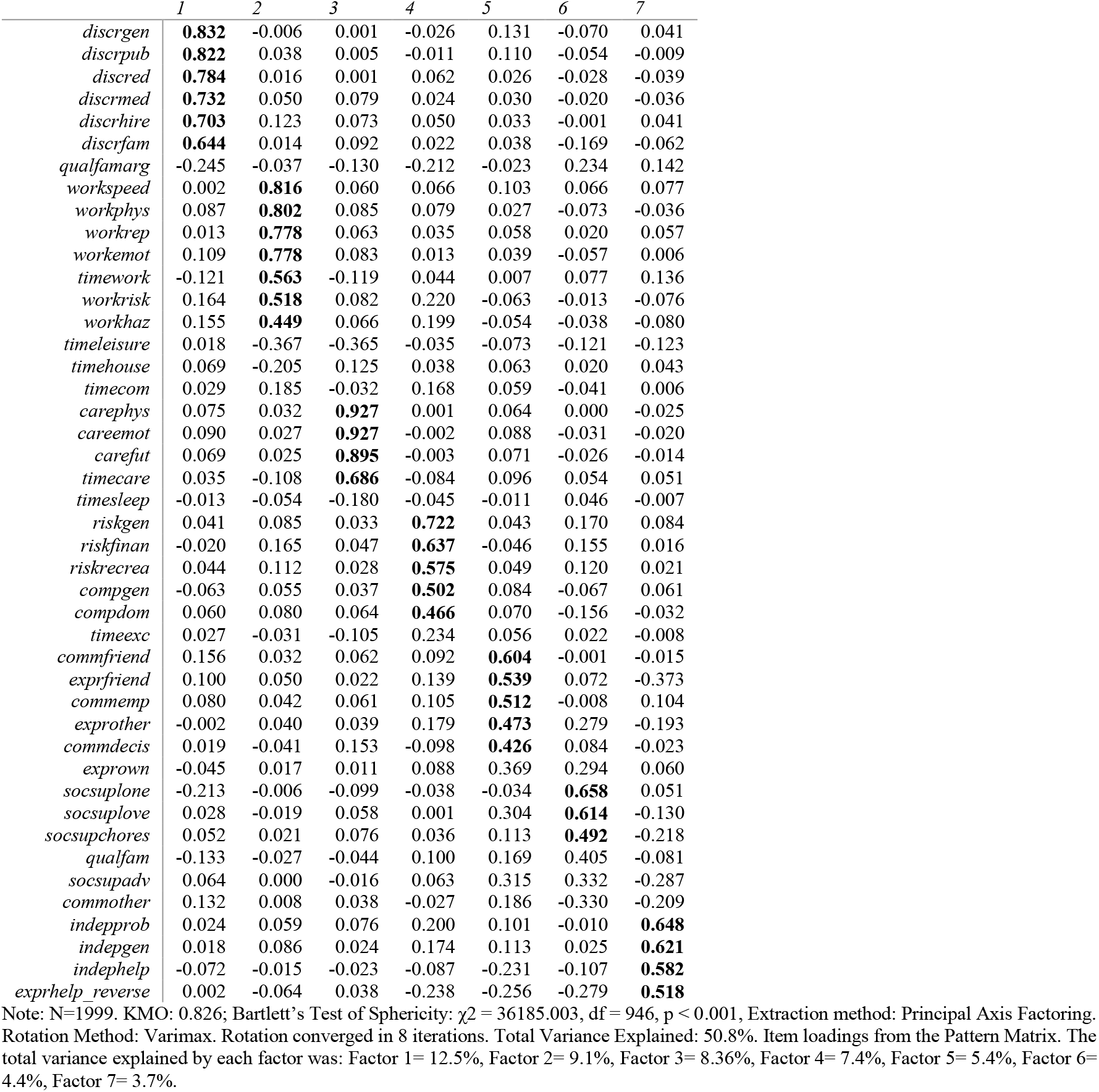
Exploratory Factor Analysis (Full factor model), Varimax rotation.

**Table S38.**
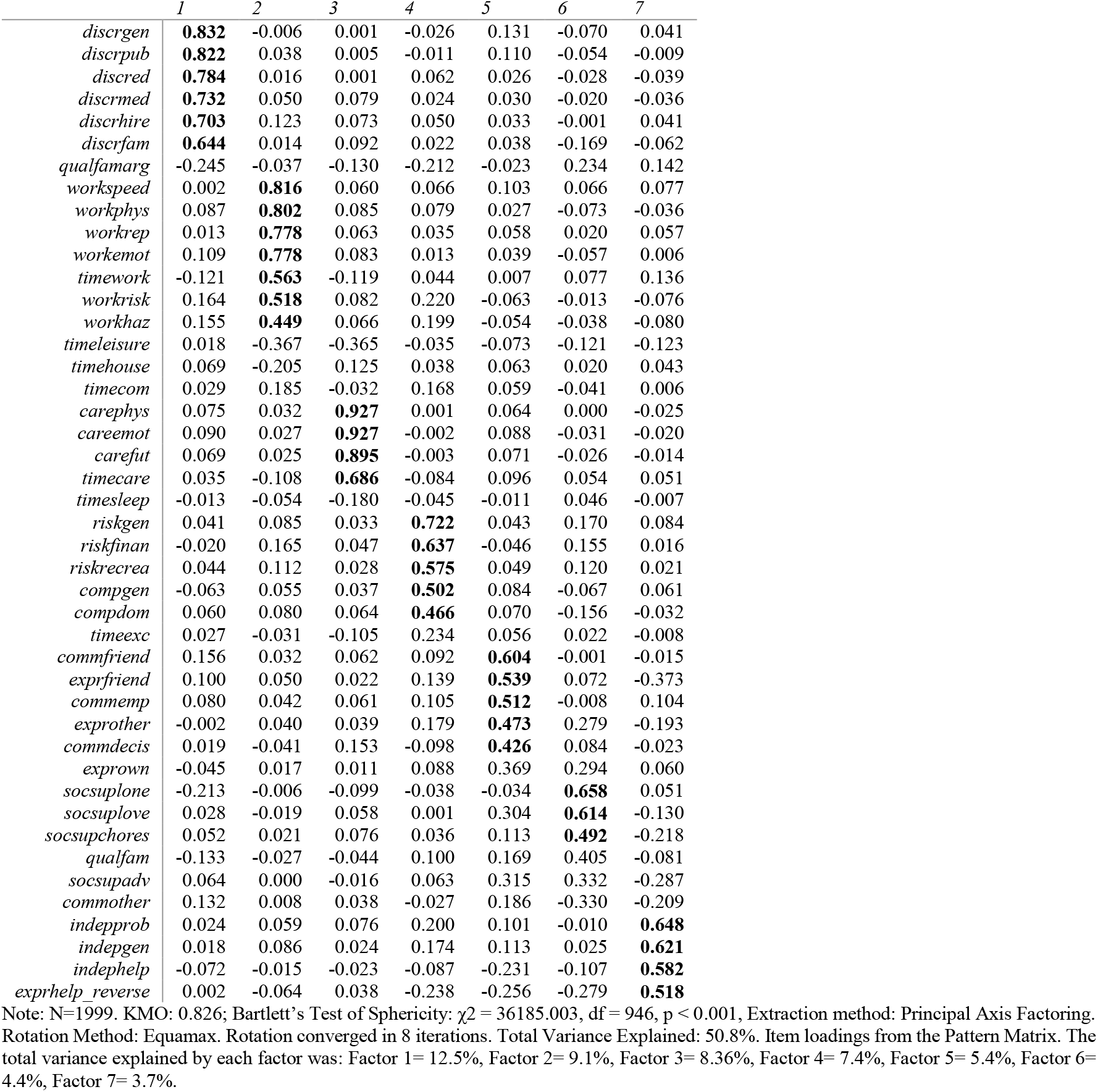
Exploratory Factor Analysis (Full factor model), Equamax rotation.

**Table S39.**
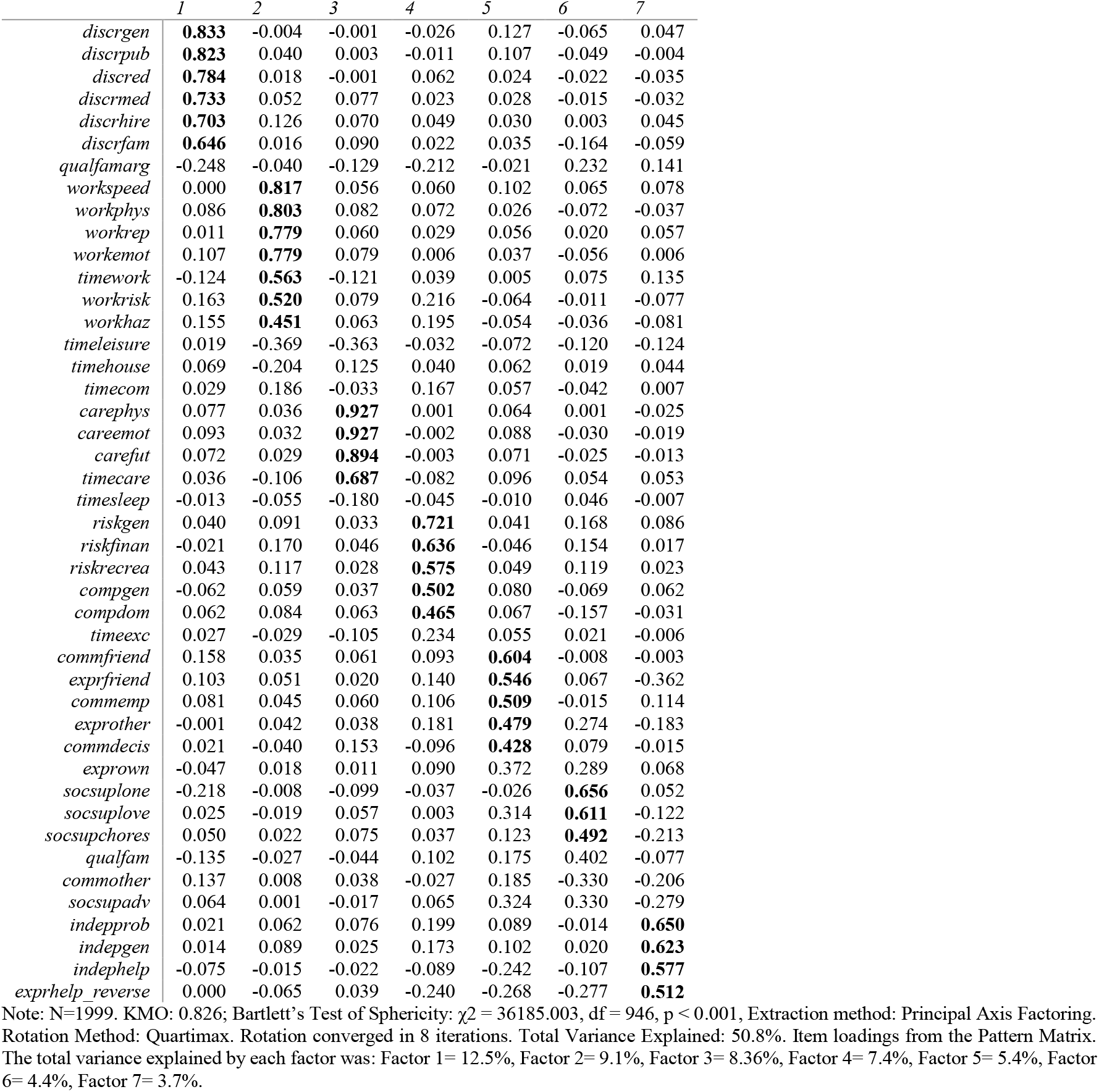
Exploratory Factor Analysis (Full factor model), Quartimax rotation.

**Table S40.**
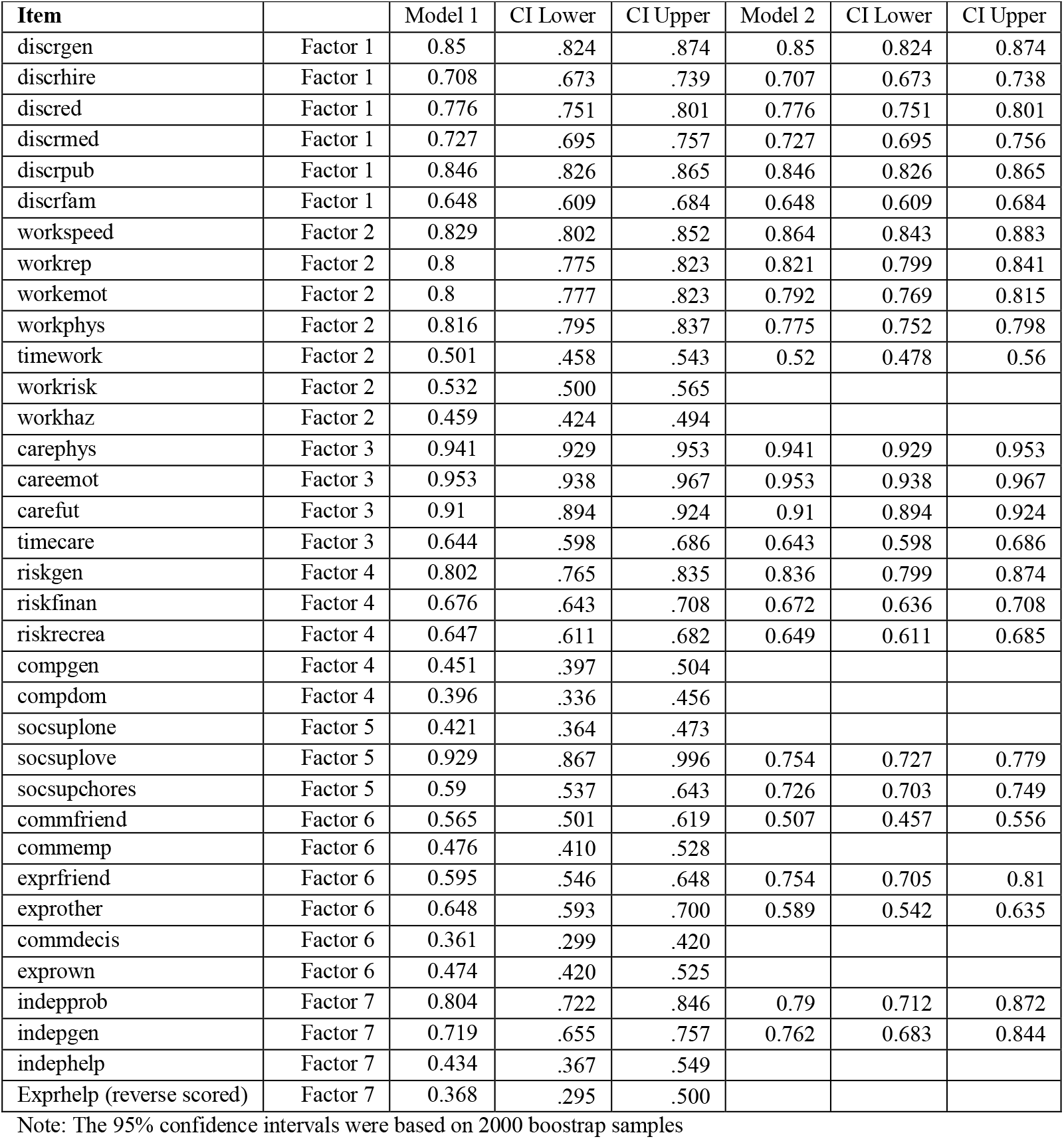
Factor loadings for CFA Models 1 and 2 in sample 1.

**Table S41.**
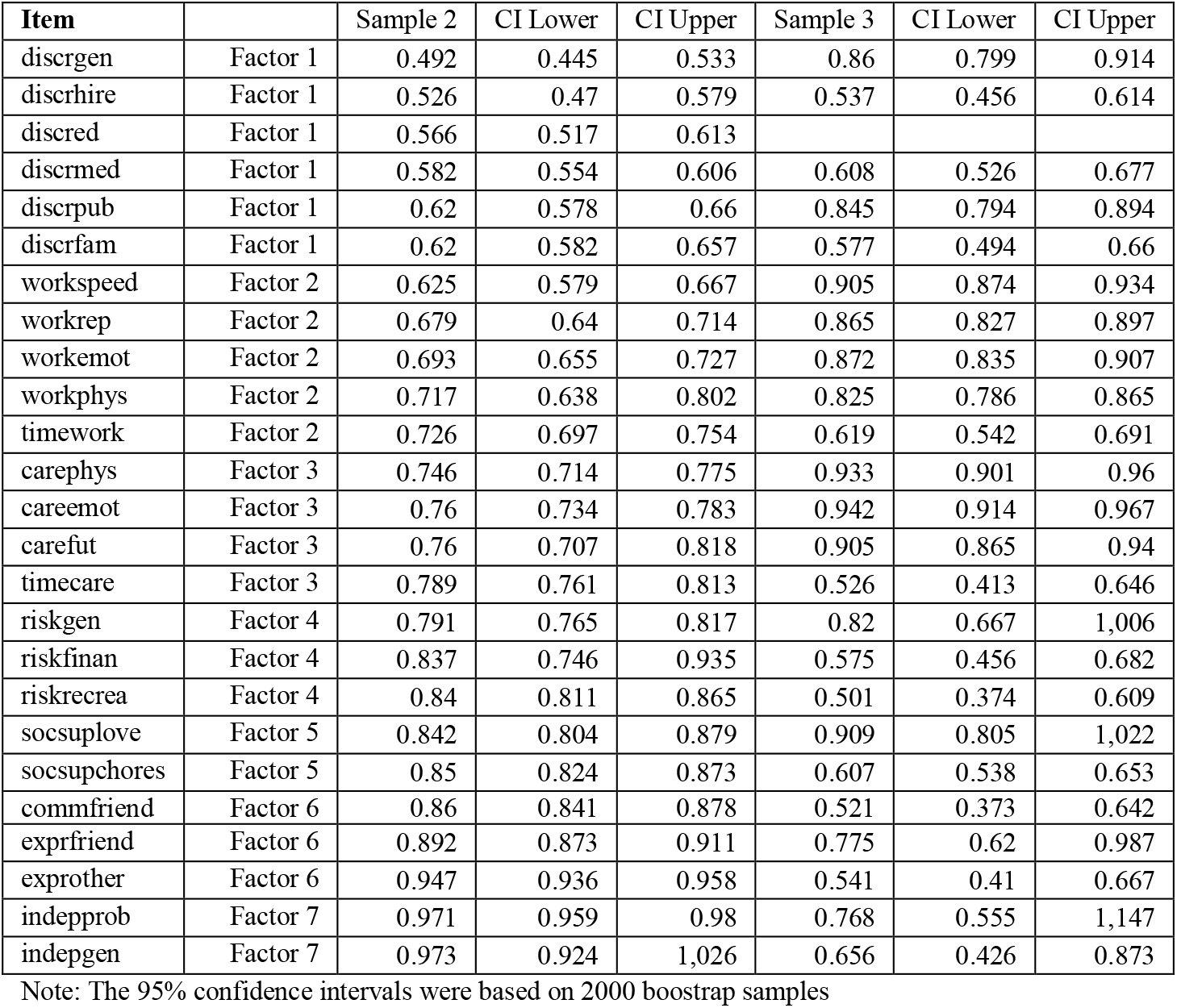
Factor loadings for CFA Samples 2 and 3 (Configural invariance).

**Table S42.**
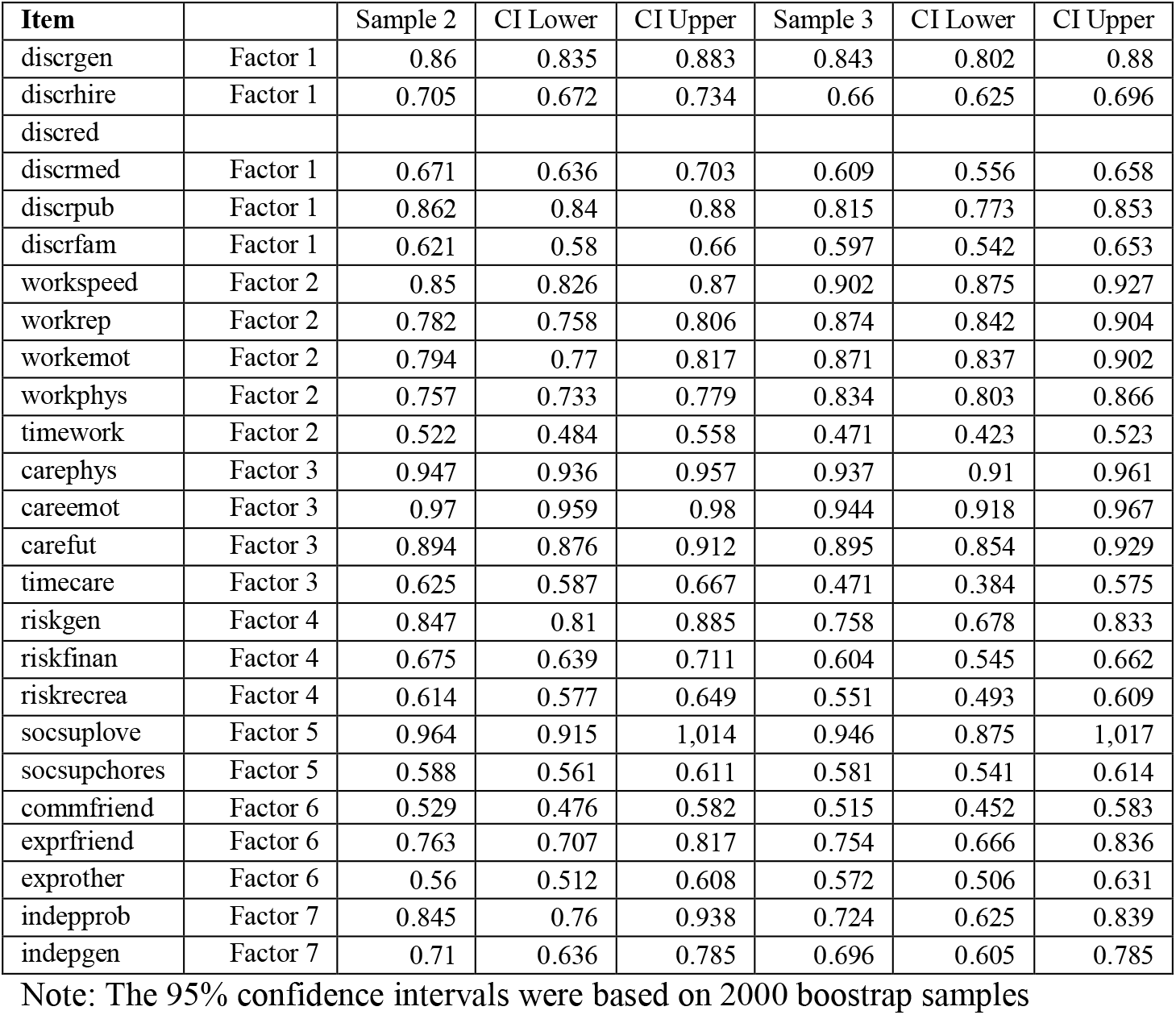
Factor loadings for CFA Samples 2 and 3 (Metric invariance, 24 items).

**Table S43.** Factor loadings for final CFA in samples 2 and 3 (Scalar invariance, 24 items).

| Item |  | Sample 2 | CI Lower | CI Upper | Sample 3 | CI Lower | CI Upper |
| --- | --- | --- | --- | --- | --- | --- | --- |
| discregen | Factor 1 | 0.859 | 0.835 | 0.883 | 0.837 | 0.792 | 0.876 |
| discrehire | Factor 1 | 0.701 | 0.667 | 0.73 | 0.64 | 0.607 | 0.675 |
| discred |  |  |  |  |  |  |  |
| discremed | Factor 1 | 0.672 | 0.636 | 0.703 | 0.614 | 0.561 | 0.664 |
| discrepub | Factor 1 | 0.863 | 0.842 | 0.882 | 0.821 | 0.777 | 0.858 |
| discrefam | Factor 1 | 0.62 | 0.579 | 0.66 | 0.6 | 0.548 | 0.654 |
| workspeed | Factor 2 | 0.851 | 0.828 | 0.871 | 0.909 | 0.883 | 0.934 |
| workrep | Factor 2 | 0.787 | 0.763 | 0.81 | 0.875 | 0.842 | 0.905 |
| workemot | Factor 2 | 0.79 | 0.764 | 0.814 | 0.875 | 0.841 | 0.906 |
| workphys | Factor 2 | 0.754 | 0.729 | 0.777 | 0.842 | 0.81 | 0.873 |
| timework | Factor 2 | 0.526 | 0.489 | 0.562 | 0.487 | 0.437 | 0.54 |
| carephys | Factor 3 | 0.947 | 0.936 | 0.957 | 0.937 | 0.91 | 0.961 |
| careemot | Factor 3 | 0.97 | 0.959 | 0.98 | 0.944 | 0.918 | 0.967 |
| carefut | Factor 3 | 0.894 | 0.876 | 0.912 | 0.895 | 0.855 | 0.929 |
| timecare | Factor 3 | 0.625 | 0.587 | 0.667 | 0.471 | 0.383 | 0.575 |
| riskgen | Factor 4 | 0.849 | 0.811 | 0.886 | 0.758 | 0.678 | 0.835 |
| riskfinan | Factor 4 | 0.673 | 0.637 | 0.708 | 0.608 | 0.548 | 0.663 |
| riskrecrea | Factor 4 | 0.613 | 0.577 | 0.648 | 0.559 | 0.5 | 0.619 |
| socsuplove | Factor 5 | 0.963 | 0.915 | 1.013 | 0.946 | 0.877 | 1.015 |
| socsupchores | Factor 5 | 0.588 | 0.562 | 0.611 | 0.584 | 0.545 | 0.617 |
| commfriend | Factor 6 | 0.533 | 0.481 | 0.584 | 0.535 | 0.471 | 0.604 |
| exprfriend | Factor 6 | 0.765 | 0.709 | 0.818 | 0.768 | 0.688 | 0.846 |
| exprother | Factor 6 | 0.555 | 0.509 | 0.602 | 0.587 | 0.522 | 0.644 |
| indepprob | Factor 7 | 0.866 | 0.776 | 0.969 | 0.731 | 0.62 | 0.861 |
| indeppgen | Factor 7 | 0.691 | 0.606 | 0.772 | 0.673 | 0.58 | 0.763 |

Note: The 95% confidence intervals were based on 2000 bootstrap samples

**Table S44.**
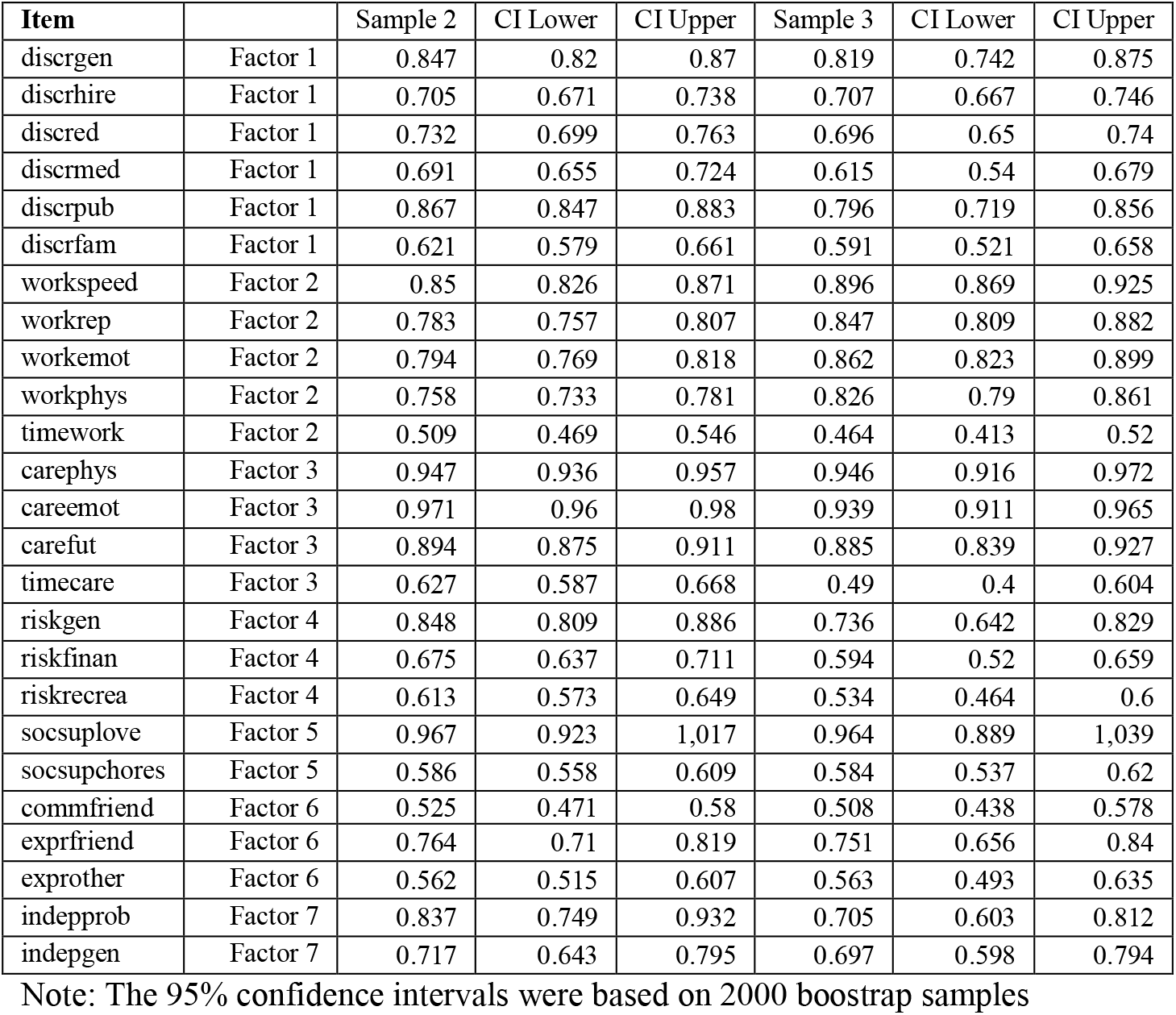
Factor loadings for final CFA in samples 2 and 3 (Metric invariance, 25 items).

**Table S45.** Factor loadings for final CFA samples 2 and 3 (Scalar invariance, 25 items).

| Item |  | Sample 2 | CI Lower | CI Upper | Sample 3 | CI Lower | CI Upper |
| --- | --- | --- | --- | --- | --- | --- | --- |
| discregen | Factor 1 | 0.846 | 0.819 | 0.869 | 0.815 | 0.731 | 0.876 |
| discrehire | Factor 1 | 0.707 | 0.674 | 0.74 | 0.674 | 0.634 | 0.717 |
| discred | Factor 1 | 0.732 | 0.7 | 0.764 | 0.683 | 0.638 | 0.73 |
| discremed | Factor 1 | 0.691 | 0.655 | 0.724 | 0.618 | 0.543 | 0.681 |
| discrepub | Factor 1 | 0.866 | 0.847 | 0.883 | 0.797 | 0.715 | 0.859 |
| discrefam | Factor 1 | 0.621 | 0.579 | 0.662 | 0.595 | 0.527 | 0.661 |
| workspeed | Factor 2 | 0.85 | 0.827 | 0.871 | 0.899 | 0.87 | 0.929 |
| workrep | Factor 2 | 0.785 | 0.759 | 0.808 | 0.841 | 0.804 | 0.876 |
| workemot | Factor 2 | 0.792 | 0.767 | 0.816 | 0.862 | 0.821 | 0.9 |
| workphys | Factor 2 | 0.758 | 0.733 | 0.781 | 0.83 | 0.793 | 0.866 |
| timework | Factor 2 | 0.508 | 0.468 | 0.545 | 0.468 | 0.416 | 0.523 |
| carephys | Factor 3 | 0.947 | 0.936 | 0.957 | 0.945 | 0.915 | 0.972 |
| careemot | Factor 3 | 0.971 | 0.96 | 0.98 | 0.939 | 0.911 | 0.966 |
| carefut | Factor 3 | 0.894 | 0.875 | 0.911 | 0.885 | 0.839 | 0.927 |
| timecare | Factor 3 | 0.627 | 0.587 | 0.668 | 0.49 | 0.4 | 0.605 |
| riskgen | Factor 4 | 0.849 | 0.81 | 0.888 | 0.73 | 0.632 | 0.825 |
| riskfinan | Factor 4 | 0.673 | 0.635 | 0.709 | 0.594 | 0.52 | 0.659 |
| riskrecrea | Factor 4 | 0.613 | 0.574 | 0.65 | 0.539 | 0.469 | 0.604 |
| socsuplove | Factor 5 | 0.966 | 0.921 | 1.016 | 0.964 | 0.891 | 1.038 |
| socsupchores | Factor 5 | 0.586 | 0.559 | 0.61 | 0.588 | 0.543 | 0.626 |
| commfriend | Factor 6 | 0.527 | 0.474 | 0.581 | 0.525 | 0.454 | 0.596 |
| exprfriend | Factor 6 | 0.765 | 0.711 | 0.819 | 0.76 | 0.669 | 0.845 |
| exprother | Factor 6 | 0.56 | 0.513 | 0.605 | 0.579 | 0.51 | 0.647 |
| indepprob | Factor 7 | 0.848 | 0.762 | 0.946 | 0.705 | 0.597 | 0.819 |
| indeppgen | Factor 7 | 0.707 | 0.631 | 0.787 | 0.685 | 0.585 | 0.784 |

Note: The 95% confidence intervals were based on 2000 bootstrap samples

